# One size fits all: A systematic review of the sample types used for the diagnostics of respiratory viruses in children

**DOI:** 10.64898/2026.06.02.26354258

**Authors:** Orchid M. Allicock, Arushi Dogra, Jacqueline H. Cho, Keyner Rojas, Hannah O. Hasson, Britney Omene, Melissa C. Funaro, Claire S. Laxton, Inci Yildirim

## Abstract

Nasopharyngeal (NP) swabs remain the dominant gold standard for respiratory infection diagnostics. While there has been increased use of alternative sample types since the COVID-19 pandemic, guidance on their use for detecting respiratory viruses is not yet definitive, especially for children. In this systematic review and meta-analysis, we aimed to compare the diagnostic accuracy and tolerability of multiple respiratory specimen types for detecting respiratory viruses in pediatric populations.

Searches were conducted on July 17, 2025 in MEDLINE, Embase, Web of Science, and Scopus, with screening and data extraction performed in Covidence. English-language primary research articles published since 2000 comparing respiratory virus detection rates in children, using nucleic acid amplification tests between paired respiratory specimens, were included. Risk of bias was assessed using Quality Assessment of Diagnostic Accuracy Studies criteria. We calculated pooled sensitivities and specificities of index specimens: nasopharyngeal aspirates (NPA), mid-turbinate swabs (MT), anterior nasal swabs (ANS), oropharyngeal swabs (OP), and bronchoalveolar lavage fluid (BAL), as compared to the reference, NP swabs, using random-effects modeling, firstly without discrimination by virus. Index specimens were then grouped by sample collection site as nasal, oral, and lower respiratory tract (LRT) specimens for virus-specific analyses. Overall performance and statistical validity were evaluated by hierarchical summary receiver operating characteristic (HSROC) analysis. Data regarding sampling tolerability was also assessed.

We screened 2,448 studies and identified 36 publications (total N participants = 10,687) that reported diagnostic test accuracy using paired index-reference data in children. Of these, 18 (total N participants = 4,310) used NP specimens as the reference and were included in the diagnostic test accuracy analysis. Virus-agnostic pooled sensitivity estimates indicated that MT (0.92%) performed most similarly to NP, though sensitivities of ANS (0.79%) and OP (0.70%) were also moderately high for detection of any respiratory virus. BAL sensitivity was the lowest (0.37%). All sample types demonstrated high specificity (0.98%-0.99%). Group estimates and HSROC statistics found that nasal specimens, when grouped, had the highest sensitivity and accuracy for all examined viruses, including for influenza (92%) and RSV (90%). By comparison, oral and LRT specimens performed less well, with more variability, though both showed moderately high sensitivities for RSV (78%, 76%, respectively) and influenza (82%, 80%, respectively), and LRT samples showed high sensitivity for HMPV (82%). Analysis of sample tolerability found that NP swabs consistently ranked as the least comfortable and least preferred, while nasal swabs and saliva both performed well. Datasets for LRT and oral specimens were sparser than for nasal, and this contributed to greater variability, underscoring the need for further diagnostic accuracy studies on alternatives to NP sampling.

These data support the viability of nasal and oral alternatives to NP swabs and affirm their application in pediatric care, particularly in outpatient settings. Such alternatives could greatly improve sampling tolerability and increase global access, including in resource-limited settings, to accurate diagnostic methods for respiratory infections.

## Background

Respiratory tract infections are one of the main causes of infant mortality globally, and account for 14% of deaths in those under 5 years of age [1, 2]. Infants have increased susceptibility to infection, particularly to respiratory viruses with high transmission rates [3, 4]. Therefore, accurate and timely diagnosis of respiratory infections is essential for both effective patient management and public health surveillance. Molecular diagnostic methods, particularly nucleic acid amplification tests (NAATs) such as reverse transcription quantitative PCR (RT-qPCR), have become the gold standard for detecting respiratory viruses owing to their high sensitivity and specificity and their ability to simultaneously detect multiple viruses using multiplex methods [5].

The performance of these diagnostic assays is influenced by several factors, including the type and quality of the clinical specimen collected. Common specimen types include nasopharyngeal swabs (NPS), oropharyngeal swabs (OP), nasopharyngeal aspirates (NPA), induced sputum (IS), and bronchoalveolar lavage (BAL) fluid. While nasopharyngeal specimens, including NPA and NPS, are accepted as the gold standard, each specimen type varies in its ability to capture viral material, which can affect the sensitivity of detection across respiratory viruses [6–8].

In an American Society for Microbiology-sponsored Practical Guidance for Clinical Microbiology document, Charlton et al (2019) report NPA are highly sensitive for the detection of influenza A/B (FLU), respiratory syncytial virus (RSV), rhinovirus/enterovirus (RV/EV), adenovirus (ADV), human metapneumovirus (hMPV), parainfluenza viruses (PIV), and coronaviruses (HCoV) [8]. However, the optimal specimen type may vary depending on the specific respiratory virus. For example, Charlton et al. (2019) also report that sensitivity with OP was high for FLU but diminished for the other viruses tested, particularly RV/EV, hMPV, and PIV. Additionally, data on the sensitivity of nasal specimens were variable and thus not included in the study.

As summarized in **Table 1**, current WHO and CDC guidance generally recommend upper respiratory tract specimens, particularly NPS or nasal specimens such as mid-turbinate (MT) swabs, as primary specimens for respiratory virus detection, with lower tract samples reserved for severe disease. Pediatric recommendations are well-defined for some pathogens, such as RSV, offering age- and severity-level-specific advice, with comprehensive meta-analyses previously conducted [9]. By contrast, for other viruses, such as PIV, no single gold-standard specimen has been formally established, and recommendations typically follow general respiratory virus collection guidance.

**Table 1.**
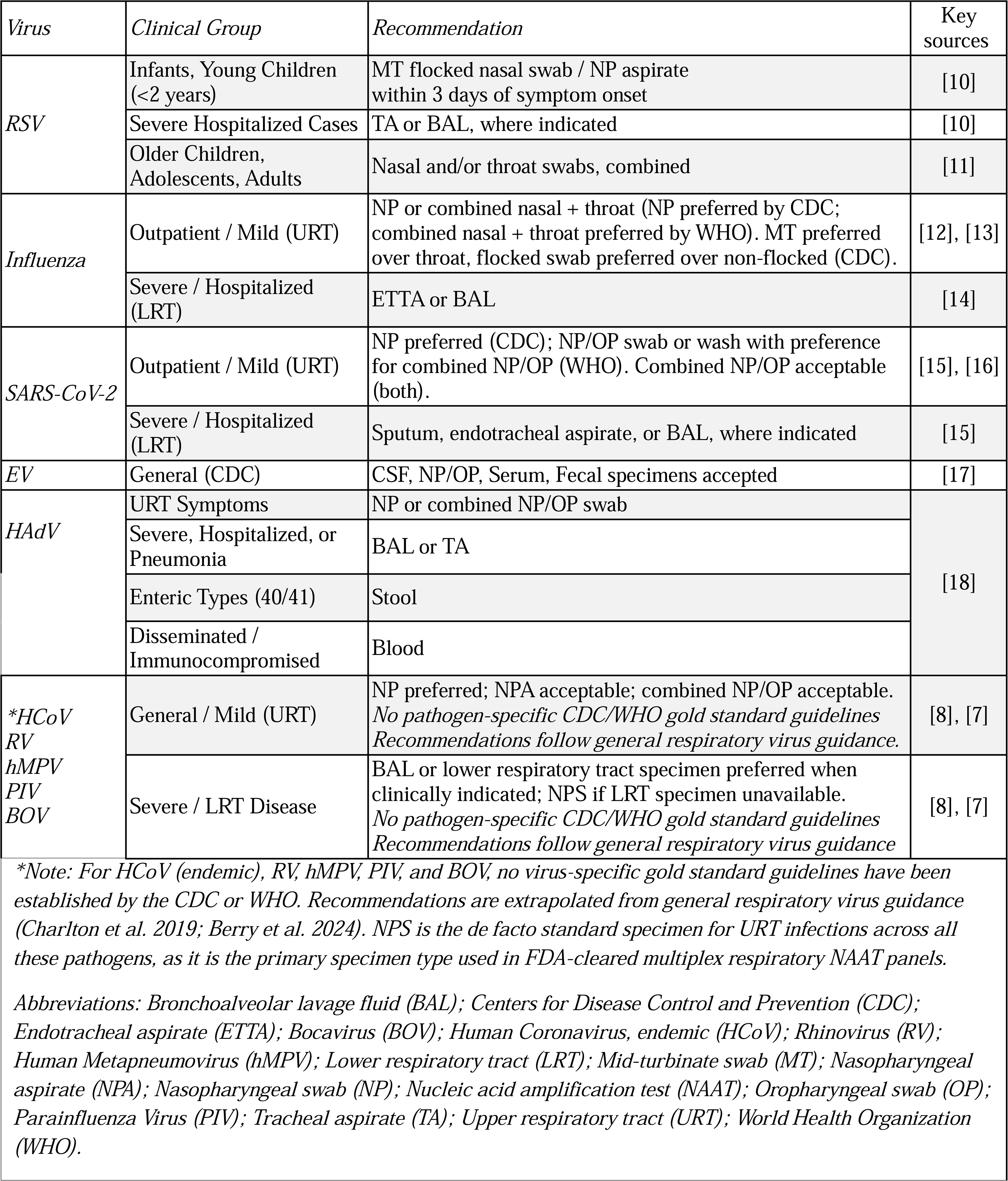
Gold Standard Specimen Guidance by Disease.

| *Virus* | *Clinical Group* | *Recommendation* | *Key sources* |
| --- | --- | --- | --- |
| RSV | Infants, Young Children (<2 years) | MT flocked nasal swab / NP aspirate within 3 days of symptom onset | [10] |
|  | Severe Hospitalized Cases | TA or BAL, where indicated | [10] |
|  | Older Children, Adolescents, Adults | Nasal and/or throat swabs, combined | [11] |
| Influenza | Outpatient / Mild (URT) | NP or combined nasal + throat (NP preferred by CDC; combined nasal + throat preferred by WHO). MT preferred over throat, flocked swab preferred over non-flocked (CDC). | [12], [13] |
|  | Severe / Hospitalized (LRT) | ETTA or BAL | [14] |
| SARS-CoV-2 | Outpatient / Mild (URT) | NP preferred (CDC); NP/OP swab or wash with preference for combined NP/OP (WHO). Combined NP/OP acceptable (both). | [15], [16] |
|  | Severe / Hospitalized (LRT) | Sputum, endotracheal aspirate, or BAL, where indicated | [15] |
| EV | General (CDC) | CSF, NP/OP, Serum, Fecal specimens accepted | [17] |
| HAdV | URT Symptoms | NP or combined NP/OP swab | [18] |
|  | Severe, Hospitalized, or Pneumonia | BAL or TA |  |
|  | Enteric Types (40/41) | Stool |  |
|  | Disseminated / Immunocompromised | Blood |  |
| *HCoV<br>RV<br>hMPV<br>PIV<br>BOV | General / Mild (URT) | NP preferred; NPA acceptable; combined NP/OP acceptable. *No pathogen-specific CDC/WHO gold standard guidelines Recommendations follow general respiratory virus guidance.* | [8], [7] |
|  | Severe / LRT Disease | BAL or lower respiratory tract specimen preferred when clinically indicated; NPS if LRT specimen unavailable. *No pathogen-specific CDC/WHO gold standard guidelines Recommendations follow general respiratory virus guidance* | [8], [7] |

*\*Note: For HCoV (endemic), RV, hMPV, PIV, and BOV, no virus-specific gold standard guidelines have been established by the CDC or WHO. Recommendations are extrapolated from general respiratory virus guidance (Charlton et al. 2019; Berry et al. 2024). NPS is the de facto standard specimen for URT infections across all these pathogens, as it is the primary specimen type used in FDA-cleared multiplex respiratory NAAT panels.*
*Abbreviations: Bronchoalveolar lavage fluid (BAL); Centers for Disease Control and Prevention (CDC); Endotracheal aspirate (ETTA); Bocavirus (BOV); Human Coronavirus, endemic (HCoV); Rhinovirus (RV); Human Metapneumovirus (hMPV); Lower respiratory tract (LRT); Mid-turbinate swab (MT); Nasopharyngeal aspirate (NPA); Nasopharyngeal swab (NP); Nucleic acid amplification test (NAAT); Oropharyngeal swab (OP); Parainfluenza Virus (PIV); Tracheal aspirate (TA); Upper respiratory tract (URT); World Health Organization (WHO).*

Despite these insights, comprehensive comparative data specifically concerning the pediatric population, across a broader range of respiratory viruses and specimen types, remain limited. Understanding how specimen type influences diagnostic sensitivity is crucial for optimizing testing strategies, reducing false negatives, and ensuring appropriate clinical and public health responses. This meta-analysis aims to address these gaps by systematically evaluating how the differences in clinical specimen types and sampling sites affect the sensitivity, specificity and diagnostic accuracy of diagnostic tests for respiratory viruses in children.

## Methods

### Protocol registration and study criteria

The study protocol was registered with PROSPERO (CRD420251164948) on 9 October 2025. The reporting of this study was guided by the standards of the Preferred Reporting Items for Systematic Reviews and Meta-analyses (PRISMA) [19]. We developed the eligibility criteria based on the Population-Intervention-Comparator-Outcomes-Time Frame-Setting (PICOTS) framework (**S1 Table**) [20]. Briefly, primary studies were eligible if they reported diagnostic test performance or compared respiratory virus detection rates across different specimens. We excluded secondary research (e.g., reviews and meta-analyses), animal studies, adult studies, and *in vitro* studies.

### Information sources and search strategy

On July 17, 2025, a comprehensive search of multiple databases was performed using MEDLINE, Embase, Web of Science, and Scopus. The search was limited to English-language articles and articles published from 2000 to the date of the search. Included studies were limited to those published from 2000 onwards, as this reflects the period at which NAATs began being used in the clinic for respiratory pathogens [21]. All search strategies are in **Data 1** in **S2 Dataset**. Search results were pooled in EndNote [www.endnote.com] and de-duplicated using the Yale Reference Deduplicator [https://library.medicine.yale.edu/reference-deduplicator]. This set was uploaded to Covidence (Veritas Health Innovation, Melbourne, Australia) for screening, which identified additional duplicates.

### Manuscript screening process and data extraction

To streamline the systematic review process, we used the Covidence software platform (Veritas Health Innovation, Melbourne, Australia). Two independent authors (OMA, AD, KR, HH, BO, JC, CSL) independently assessed eligibility via abstract screening and completed full-text review and manual data extraction in Covidence. Any disagreements were resolved by OMA or AD. To calculate the sensitivity and specificity of each specimen, relevant data, including the total number of samples tested, the total true positives/negatives (positive/negative in both reference and index), and the total false positives/negatives (positive/negative in index but not reference), were recorded. The reference and index specimen(s) designations were retained according to the original study’s designations. The data extraction protocol, including the list if data items extracted, is available in **Data 2, S2 Dataset**.

### Risk of Bias and Applicability

Study quality assessment was performed simultaneously with full text data extraction by two independent reviewers using Quality Assessment of Diagnostic Accuracy Studies (QUADAS-2) criteria, with exact methodology detailed in **Data 3** in **S2 Dataset** [22]. Disagreements were resolved by consensus. The results of the risk assessment were compiled using the Robvis package (v0.3.0.9) [23].

### Tolerability analysis

For those studies that measured sample collection discomfort on a Likert-type scale, discomfort was normalized into a percentage of the scale maximum and then separated by respondent (child, caregiver, or sample taker). Each value was weighted by N/sum of all respondents for that sample type (**Data 4** in **S2 Dataset**).

### Data analysis

For included studies in which true/false positive and negative numbers had been reported, a meta-analysis of diagnostic test accuracy (DTA) was performed. Data from 2-by-2 tables were used to measure diagnostic tests’ sensitivity and specificity, calculate 95% confidence intervals (CIs), and present the results graphically in forest plots, bar plots, and Hierarchical Summary Receiver Operating Characteristic (HSROC) curves. Data with null sensitivity or null specificity values were not included in the respective sensitivity or specificity analyses. The meta and mada packages in R were used; metaprop objects were used to determine specificity and sensitivity values and to formulate the forest plots, and bivariate reitsma objects were used to fit random-effects HSROC models, plotting sensitivity against specificity for each comparison [24–26].

Pooled random-effects sensitivity and specificity estimate in forest plots were calculated using logit transformation, then back-transformed. The data was fitted to a generalized linear mixed model, and confidence interval values were used with the Hartung–Knapp adjustment to account for low study count. The pooled estimates were weighted by inverse-variance to account for studies with large variances. Summary bar plots were created using ggplot2 and plotted the point estimates and confidence intervals of the metaprop objects. Statistical significance was assessed using p-values, where p-values <0.05 were considered statistically significant.

The accuracy (Λ), threshold (Θ), and shape (β) parameters of each HSROC curve is reported. Area Under the Curve (AUC) and Diagnostic Odds Ratio (DOR) were derived by the reitsma object and the correlation coefficient (r) and Q* index, where sensitivity equals specificity, were calculated using model intercepts. All analyses were performed in R version 4.5.2.

## Results

### Study selection process and search results

Our search strategy identified 2,448 unique references, of which 200 studies met the criteria for full-text screening (**Fig 1**). Following the full text review, 164 studies were excluded. Exclusion was primarily related to the methodology of specimen comparison; most excluded studies either did not include more than one respiratory specimen, did not pair specimens (or did not collect them within 72 hours of each other), or did not test paired specimens using NAAT. Two studies in which paired specimens were collected more than 24 hours apart were retained but flagged as high risk of bias in the flow and timing domain of the QUADAS-2 assessment. The search identified 36 publications, including 10,687 participants, that met the inclusion criteria and were included in the final analysis. All extracted data items are summarized in **Table 2**.

**Fig 1.**
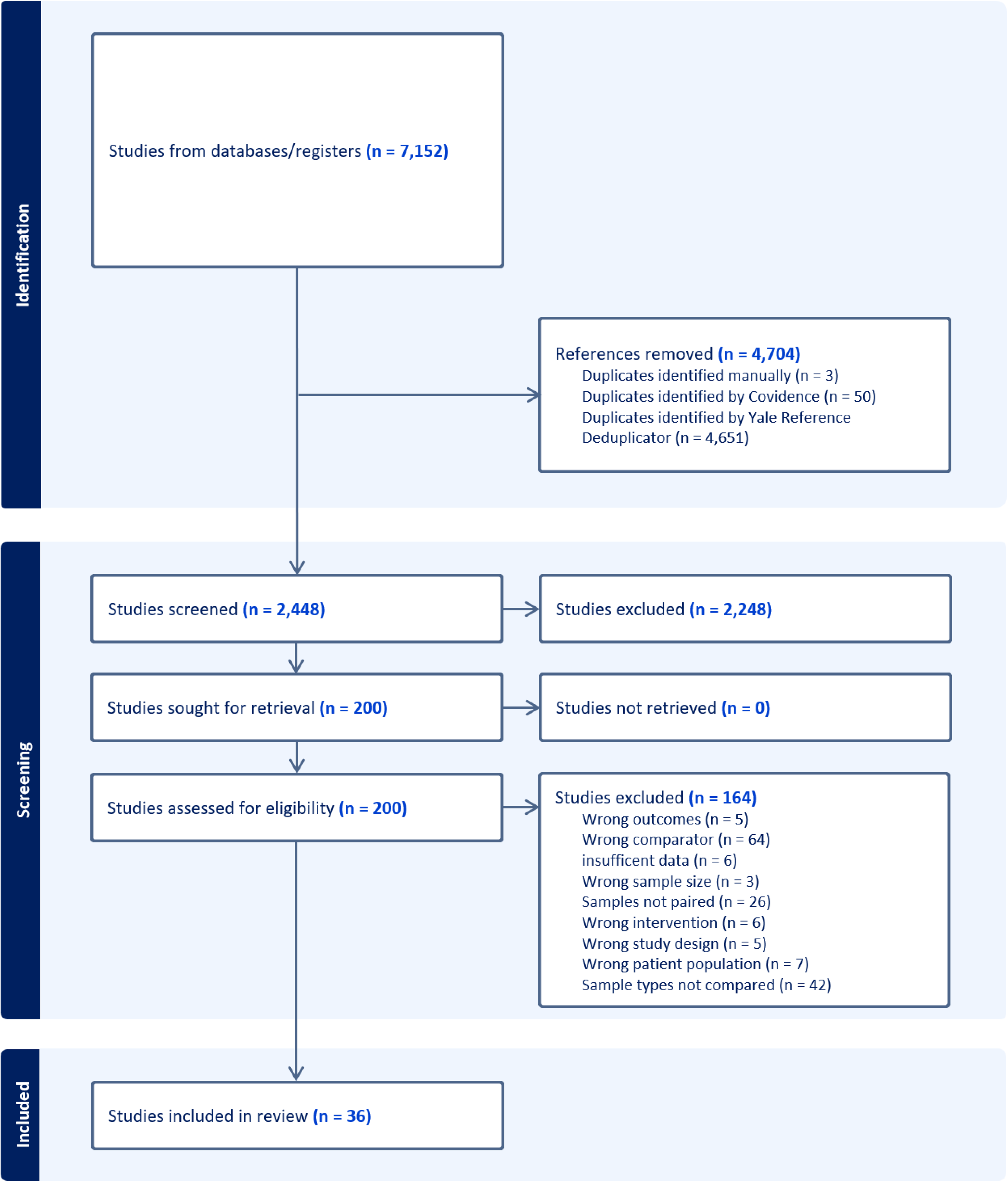
PRISMA flow chart showing the selection of publications included in the study. A total of 2448 studies were screened, and 36 studies were included. The majority of excluded studies did not compare two appropriate respiratory specimens or did not collect and present paired data.

**Table 2.**
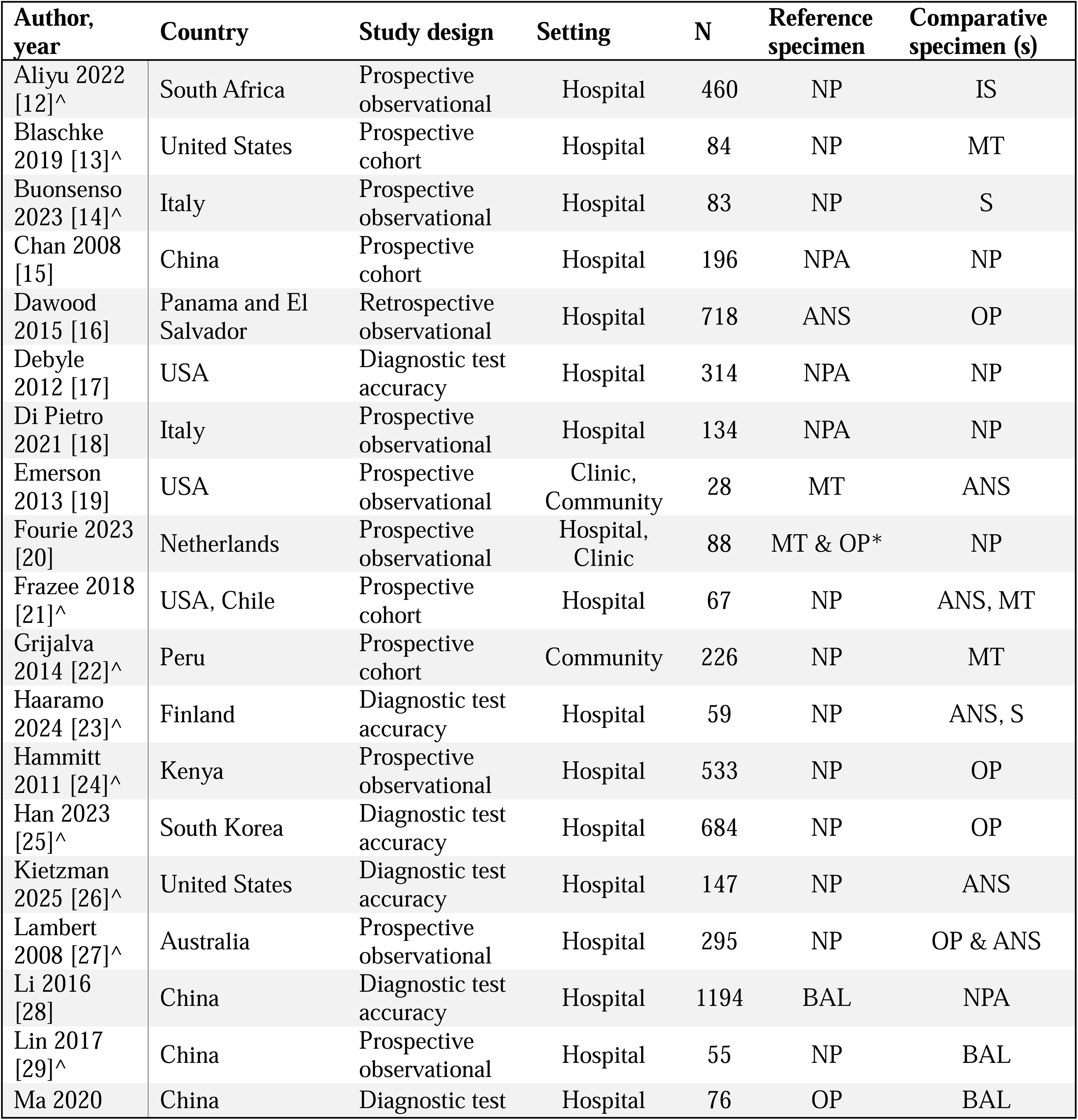
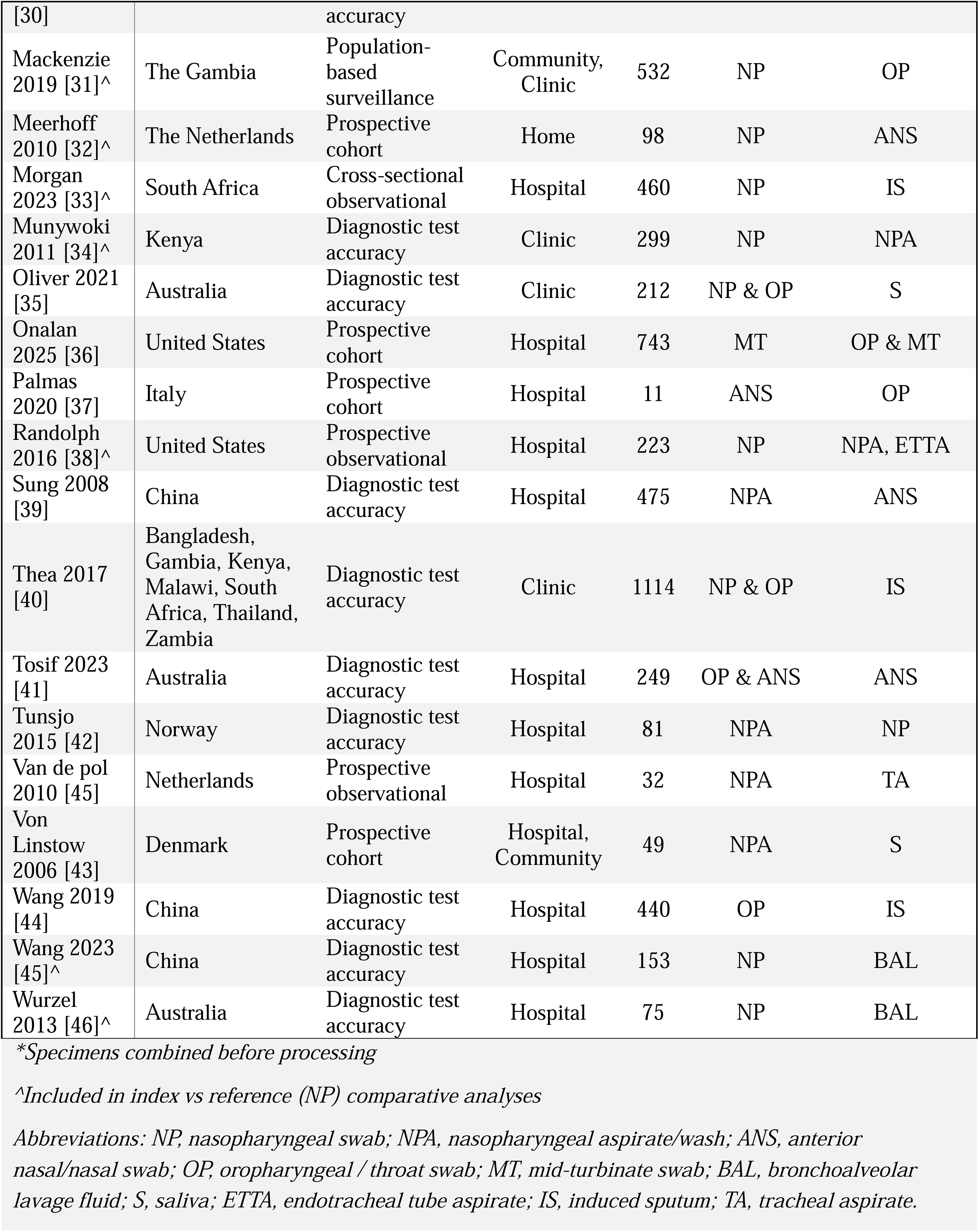
Location, size, and specimen types of included studies.

### Included studies

The systematic review included 36 publications, published between 2006 and 2025. As outline in **Table 3**, studies were most frequently conducted in the Western Pacific Region (32.4%), followed by the Americas and Europe (24.3%), and Africa (16.2%). China and the United States were the most represented countries (each at 15.9%), followed by Australia (9.1%).

**Table 3.**
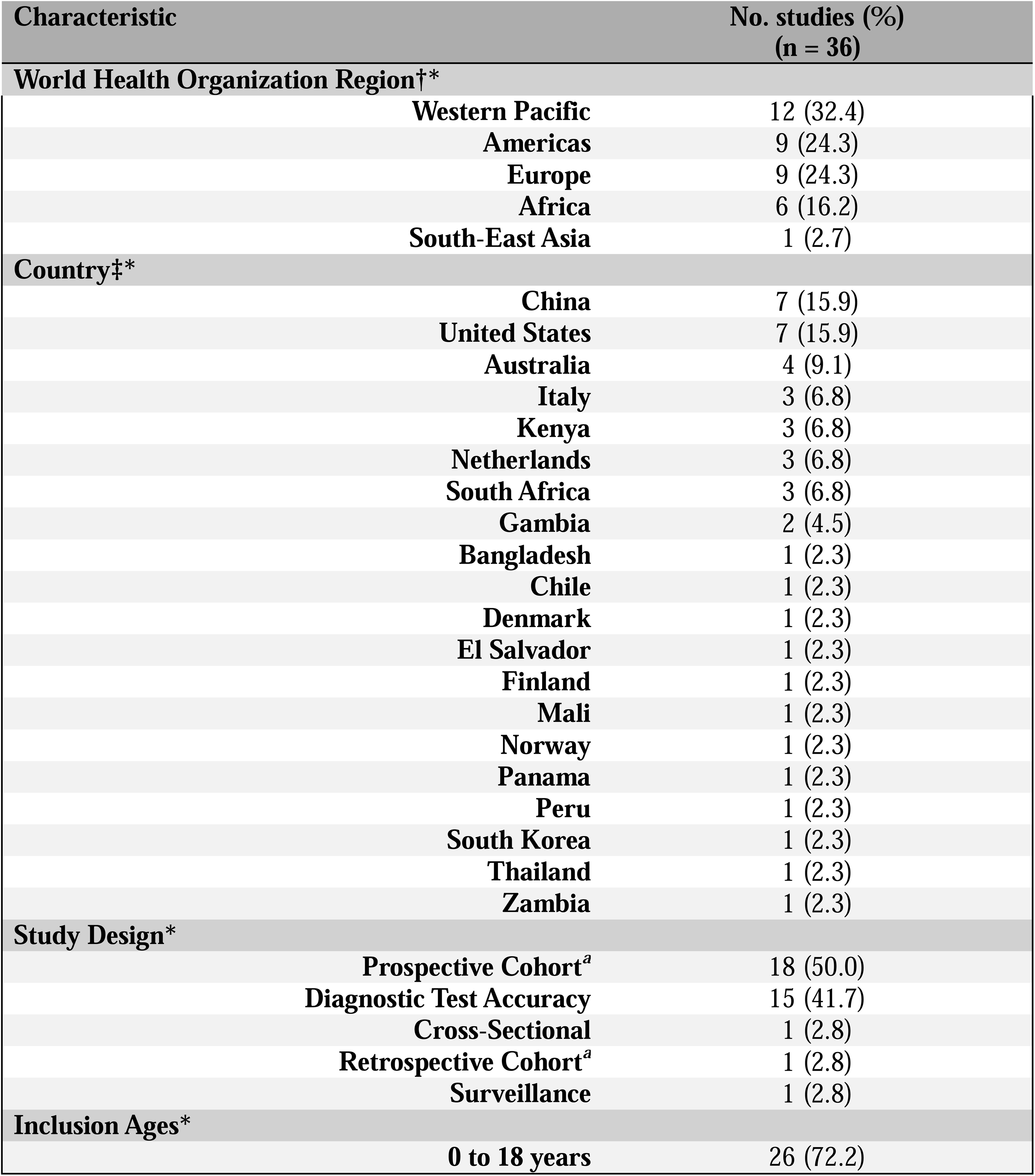
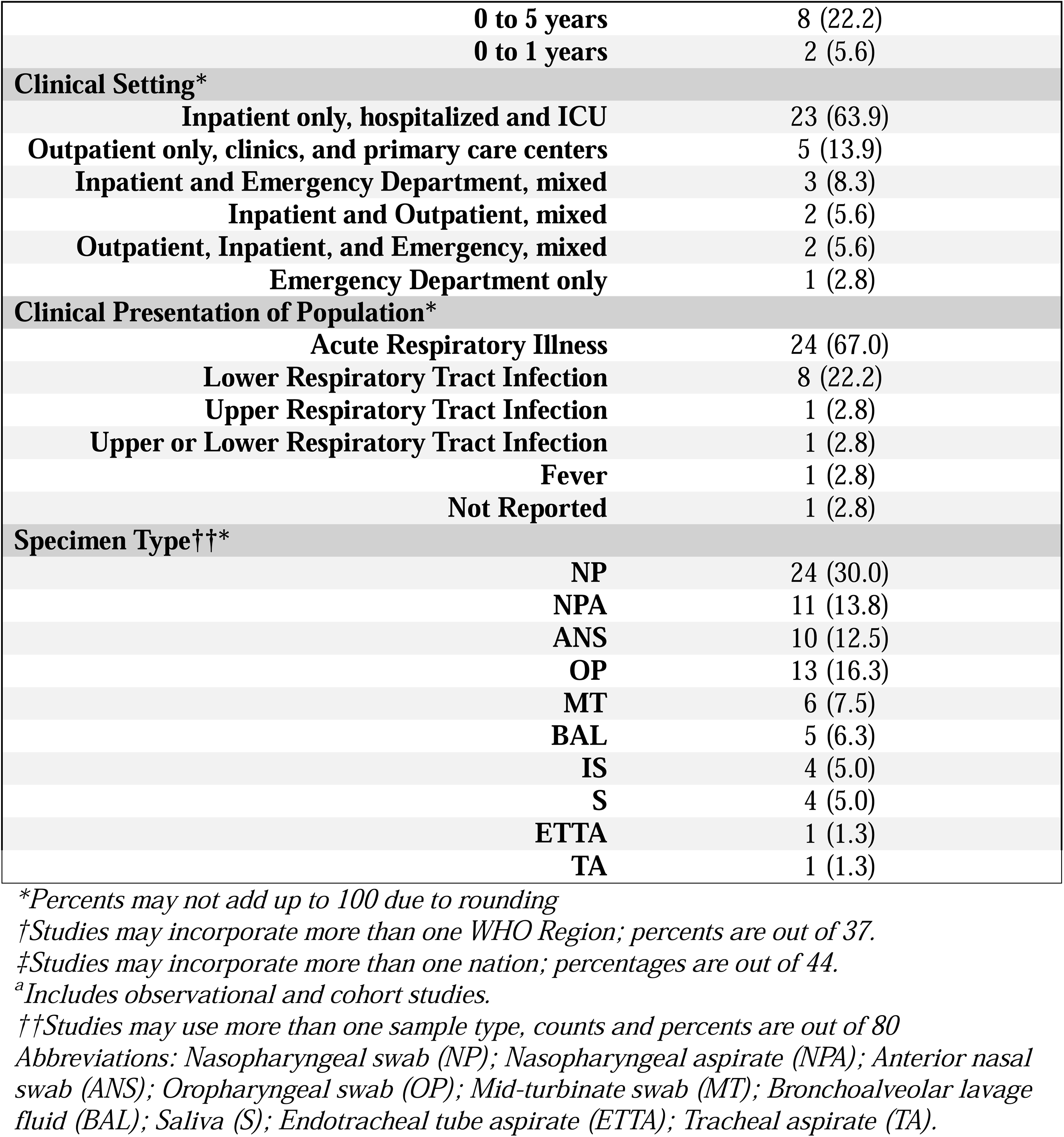
Study characteristics.

| Characteristic | No. studies (%)<br>(n = 36) |
| --- | --- |
| **World Health Organization Region†\*** |  |
| Western Pacific | 12 (32.4) |
| Americas | 9 (24.3) |
| Europe | 9 (24.3) |
| Africa | 6 (16.2) |
| South-East Asia | 1 (2.7) |
| **Country‡\*** |  |
| China | 7 (15.9) |
| United States | 7 (15.9) |
| Australia | 4 (9.1) |
| Italy | 3 (6.8) |
| Kenya | 3 (6.8) |
| Netherlands | 3 (6.8) |
| South Africa | 3 (6.8) |
| Gambia | 2 (4.5) |
| Bangladesh | 1 (2.3) |
| Chile | 1 (2.3) |
| Denmark | 1 (2.3) |
| El Salvador | 1 (2.3) |
| Finland | 1 (2.3) |
| Mali | 1 (2.3) |
| Norway | 1 (2.3) |
| Panama | 1 (2.3) |
| Peru | 1 (2.3) |
| South Korea | 1 (2.3) |
| Thailand | 1 (2.3) |
| Zambia | 1 (2.3) |
| **Study Design\*** |  |
| Prospective Cohort<sup>a</sup> | 18 (50.0) |
| Diagnostic Test Accuracy | 15 (41.7) |
| Cross-Sectional | 1 (2.8) |
| Retrospective Cohort<sup>a</sup> | 1 (2.8) |
| Surveillance | 1 (2.8) |
| **Inclusion Ages\*** |  |
| 0 to 18 years | 26 (72.2) |

|  |  |  |
| --- | --- | --- |
|  | **0 to 5 years** | 8 (22.2) |
|  | **0 to 1 years** | 2 (5.6) |
| **Clinical Setting\*** |  |  |
|  | **Inpatient only, hospitalized and ICU** | 23 (63.9) |
|  | **Outpatient only, clinics, and primary care centers** | 5 (13.9) |
|  | **Inpatient and Emergency Department, mixed** | 3 (8.3) |
|  | **Inpatient and Outpatient, mixed** | 2 (5.6) |
|  | **Outpatient, Inpatient, and Emergency, mixed** | 2 (5.6) |
|  | **Emergency Department only** | 1 (2.8) |
| **Clinical Presentation of Population\*** |  |  |
|  | **Acute Respiratory Illness** | 24 (67.0) |
|  | **Lower Respiratory Tract Infection** | 8 (22.2) |
|  | **Upper Respiratory Tract Infection** | 1 (2.8) |
|  | **Upper or Lower Respiratory Tract Infection** | 1 (2.8) |
|  | **Fever** | 1 (2.8) |
|  | **Not Reported** | 1 (2.8) |
| **Specimen Type††\*** |  |  |
|  | **NP** | 24 (30.0) |
|  | **NPA** | 11 (13.8) |
|  | **ANS** | 10 (12.5) |
|  | **OP** | 13 (16.3) |
|  | **MT** | 6 (7.5) |
|  | **BAL** | 5 (6.3) |
|  | **IS** | 4 (5.0) |
|  | **S** | 4 (5.0) |
|  | **ETTA** | 1 (1.3) |
|  | **TA** | 1 (1.3) |

\*Percents may not add up to 100 due to rounding
†Studies may incorporate more than one WHO Region; percents are out of 37.
‡Studies may incorporate more than one nation; percentages are out of 44.
<sup>a</sup>Includes observational and cohort studies.
††Studies may use more than one sample type, counts and percents are out of 80
Abbreviations: Nasopharyngeal swab (NP); Nasopharyngeal aspirate (NPA); Anterior nasal swab (ANS); Oropharyngeal swab (OP); Mid-turbinate swab (MT); Bronchoalveolar lavage fluid (BAL); Saliva (S); Endotracheal tube aspirate (ETTA); Tracheal aspirate (TA).

Half of the studies were prospective cohort designs (50.0%), and most of the remainder were diagnostic test accuracy studies (41.7%). Most studies enrolled pediatric populations spanning infancy through adolescence, with a few restricted to younger age ranges (0-5 years: 22.2%; 0-1 year: 5.6%). Study settings were predominantly inpatient or hospital-based (63.9%), with fewer outpatient-only studies (13.9%), and the remainder were conducted in mixed inpatient/outpatient and/or emergency department settings.

Most studies enrolled children with acute respiratory illness (67.0%), while 22.2% focused on lower respiratory tract infection. Across all included comparisons, including both reference and index tests, NPS were the most frequently used (30.0%), followed by OP or throat (TS) swabs (16.3%), and finally by ANS (12.5%) and MT swabs (7.5%). Lower respiratory specimens, including IS, endotracheal tube aspirates (ETTA), tracheal aspirates (TA), and BAL, as well as saliva (S) specimens, were less commonly represented.

### Risk of Bias and Applicability

Overall, 3 of 36 studies (8.3%) were judged to have a high risk of bias in at least one QUADAS-2 domain. Quality assessment results are summarized in **Fig. 2** and reported in full in **Data 4, S2 Dataset**. In the patient selection domain, most studies were judged to be at low risk of bias (32 studies, 88.9%); however, 3 studies (8.3%) had no information due to insufficient reporting of sampling methods or setting, and 1 study (2.8%) was rated as high risk. In the index test domain, 34 studies (94.4%) were assessed as having low risk of bias, with 2 (5.6%) rated as having no information; no studies were rated as high risk. The reference standard domain demonstrated the lowest overall risk, with 35 studies (97.2%) at low risk and 1 study (2.8%) rated as no information; no studies were rated as high risk. In the flow and timing domain, most studies demonstrated appropriate test timing and use of reference standards (30 studies, 83.3%), but 3 studies (8.3%) were rated as high risk due to incomplete inclusion of patients in the analysis, and 3 studies (8.3%) had no information. Applicability concerns were minimal across domains, with most studies closely aligned with the review question and target population.

**Fig 2.**
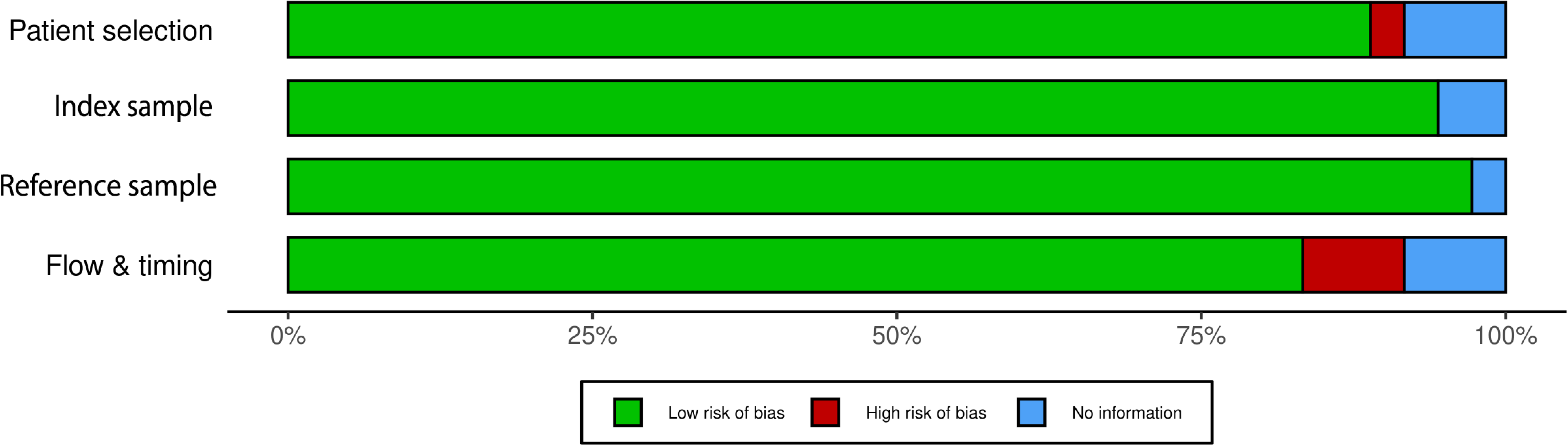
Risk of bias and applicability concerns for included studies assessed using the QUADAS-2 tool. Each bar represents the proportion of studies (n=36) rated as low risk of bias (green), high risk of bias (red), or insufficient information (blue) across the four QUADAS-2 domains: patient selection, index test, reference standard, and flow and timing. Most studies were rated as low risk across all domains. High risk of bias was most prevalent in the flow and timing domain, while lack of reporting contributed to “no information” ratings, particularly in the patient selection and index domains.

### Meta-analysis results

Across the 36 papers included per the selection criteria, the viruses covered and their respective study counts were: RSV, 26 studies; PIV, 23 studies; ADV, 22 studies; FLU, 21 studies; hMPV, 20 studies; RV/EV and Parechoviruses (PeV), 19 studies; HCoV, 16 studies; Bocavirus (BOV), 9 studies; SARS-CoV-2, 8 studies. The members of the *Picornaviridae* family, RV, EV, and PeV, were grouped during data analysis, as they were often pooled in diagnostic panels that did not distinguish them [27–30]. Forest plots summarizing all diagnostic data for some of the most common viruses in these studies, RSV, ADV, FLU, PIV, hMPV, and RV/EV/PeV are shown in **S3 Fig**, with pooled estimates included for subgrouping according to reference specimen.

To establish valid comparability for the central analysis, the 18 studies, including 4,310 participants, utilizing NPS as the reference test were identified and isolated, as indicated in **Table 2**. This subset included the 3 (16.67%) studies with a high risk of bias in at least one domain; one study (5.56%) had a high risk of bias in patient selection and 3 (16.67%) had a high risk of bias in flow and timing. Each study examined multiple viruses, and some covered multiple index tests [31–33]. After removing data with null sensitivity or null specificity from each analysis, these studies produced 104 sets of diagnostic sensitivity data and 105 sets of diagnostic specificity data when stratified by viruses. The index tests and respective data set counts were: anterior nasal swabs (ANS), 24 data sets; BAL, 18 data sets; S, 17 data sets; NPA, 12 sensitivity/13 specificity data sets; OP, 11 data sets; MT, 8 data sets; ETTA, 7 data sets; IS, 2 data sets; and mixed ANS and OP swab (ANS + OP), 5 data sets.

Initial analysis of pooled estimates comparing the diagnostic sensitivity and specificity of the most common specimens to NP, including data for all viruses, suggests the highest pooled sensitivity in NPA (0.85) and MT (0.92), with ANS (0.79) and OP (0.70) close behind. Although the pooled BAL sensitivity estimate is significantly lower (0.37), it is accompanied by an extremely large 95% confidence interval. Specificity values are high (≥ 0.98) across the board (**Fig 3**).

**Fig 3.**
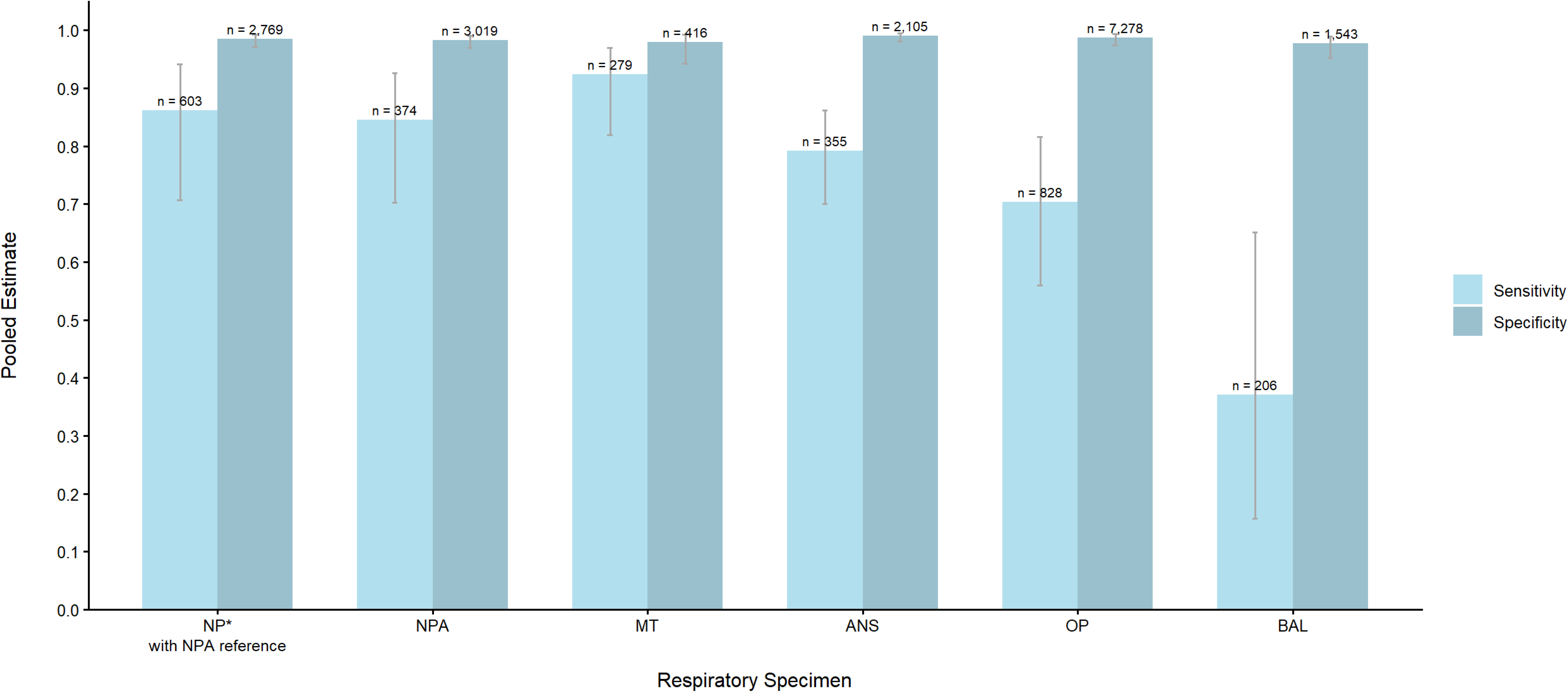
Sensitivity and Specificity of Viral Detection by Index Test with NP Reference. Pooled sensitivity and specificity estimates of the diagnostic validity of index specimens (NPA, MT, ANS, OP, and BAL) from studies utilizing an NP reference are shown across all respiratory viruses. The first bar is an exception, depicting the pooled sensitivity and specificity of an NP index against an NPA reference. Pooled estimates are weighted by inverse-variance. Error bars depict 95% confidence intervals. Sample sizes (n) above each bar represent the total denominator for the respective statistics. For sensitivity estimates, n is the sum of True Positives and False Negatives. For specificity estimates, n is the sum of True Negatives and False Positives. All data are available in detail in the corresponding forest plots in S3 Fig. of the supporting information. Abbreviations: Nasopharyngeal swab (NP); Nasopharyngeal aspirate (NPA); Mid-turbinate swab (MT); Anterior nasal swab (ANS); Oropharyngeal swab (OP); Bronchoalveolar lavage fluid (BAL)

In order to contextualize NP as a reference specimen, the four studies and 14 corresponding data sets examining NP as an index specimen against an NPA reference were also pooled, and the data is depicted in the first bar of **Fig 3**. Compared with an NPA standard, NP demonstrated sensitivity (r = 0.86) and specificity (r = 0.99) very similar to those of an NPA index vs an NP reference, with 95% confidence intervals overlapping when all viruses were included. The full forest plots for each comparison are summarized in Fig 3 and can be found in the **S4 Fig**.

Individual specimen types were then grouped into four clinically meaningful categories, with NP/NPA treated as the reference standard. Nasal specimens pooled ANS, MT, and rhinorrhea (RA) swabs. Oral specimens pooled OP, TS, and saliva collections. Lower respiratory tract (LRT) specimens pooled BAL, IS, ETTA, and TA. Sensitivity estimates were then analyzed within each pooled category and stratified by viruses. Exact pooled estimates are shown in **S5 Fig**. As shown in **Fig 4**, across viruses, nasal specimens generally yielded the highest pooled sensitivities relative to the NP reference, whereas oral specimens were typically lower and more variable. LRT specimens showed virus-dependent performance with notably wide confidence intervals, reflecting small numbers of studies and/or small sample sizes. This was particularly apparent for LRT comparisons among ADV, EV/RV/PeV, hMPV, and HCoV.

**Fig 4.**
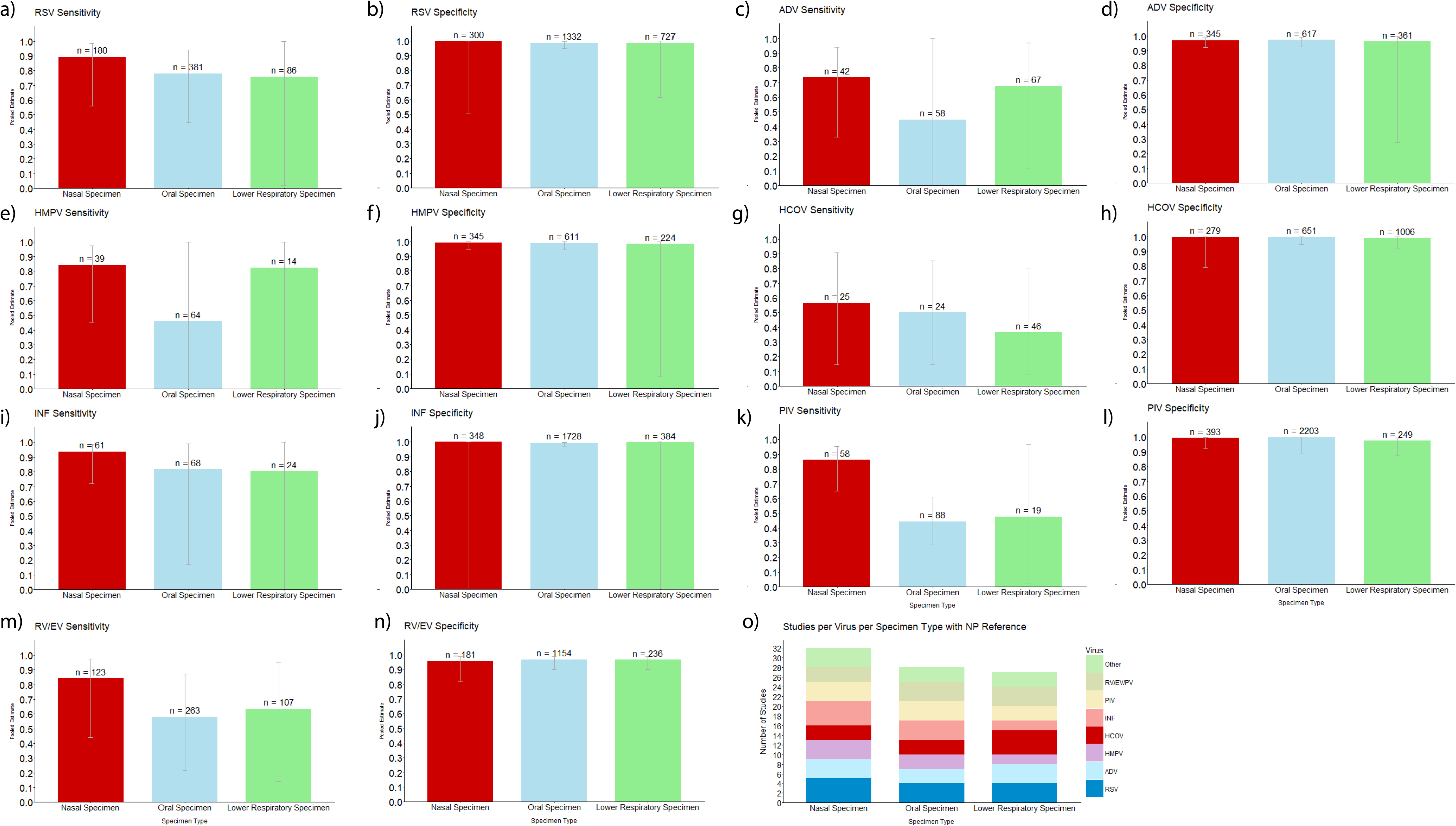
Sensitivity and Specificity of Nasal, Oral, and Lower Respiratory Specimens in the Detection of Several Viruses Against NP Reference. Pooled sensitivity estimates of the diagnostic validity of respiratory specimens (categorized as nasal, oral, or lower respiratory specimens) against an NP reference specimen are shown for (A) RSV, (C) ADV, (E) hMPV, (G) HCoV, (I) INF, (K) PIV, and (M) RV/EV. Respective specificity estimates are also shown for (B) RSV, (D) ADV, (F) hMPV, (H) HCoV, (J) INF, (L) PIV, and (N) RV/EV. Nasal specimens include ANS, MT, and RA. Oral specimens include OP and saliva. Lower respiratory specimens include BAL, ETTA, TA, and IS. Pooled estimates are weighted by inverse-variance. Error bars depict 95% confidence intervals, and sample sizes (n) above each bar represent the total denominator for the respective statistic, i.e., True Positives + False Negatives for sensitivity estimates and True Negatives + False Positives for specificity estimates. Finally, (H) depicts a bar plot of the number of data sets extracted and used in this analysis by virus and specimen type. Abbreviations: Respiratory syncytial viruses (RSV); Adenovirus (ADV); Human metapneumovirus (hMPV); Human coronaviruses (HCoV); Influenza (INF); Parainfluenza viruses (PIV); Rhinovirus (RV); Enteroviruses (EV); Nasopharyngeal swab (NP); Mid-turbinate swab (MT); Anterior nasal swab (ANS); Rhinorrhea (RA); Oropharyngeal swab (OP); Bronchoalveolar lavage fluid (BAL); Endotracheal tube aspirate (ETTA); Tracheal aspirate (TA); Induced sputum (IS)

For FLU testing, sensitivity was high and broadly similar across specimen groups, with nasal specimens most sensitive at 0.92, followed by oral at 0.82, and LRT specimens at 0.80. Most RSV data came from oral sampling, yet the pooled estimate still favored nasal collection overall (0.90), with oral (0.78) and LRT (0.76) both showing sensitivity. With PIV, nasal specimens (0.86) substantially outperformed oral (0.44) and LRT collection (0.49), with LRT showing the lowest sensitivity. In contrast, nasal (0.74) and LRT (0.69) specimens were higher than oral (0.46) for ADV testing, indicating that ADV detection may be preserved in both upper nasal and lower airway sampling. For hMPV, nasal specimens had the highest sensitivity (0.85), closely followed by LRT (0.82), whereas oral specimens were notably lower (0.46). For HCoV, nasal specimens showed the highest sensitivity (0.57), compared with oral (0.50) and LRT (0.37), though all three groups had wide confidence intervals and no strong separation by specimen type. Within the grouped RV, EV, and PeV category, nasal specimens had the highest pooled sensitivity (0.85), followed by LRT (0.64) and oral (0.61) specimens. Notably, the EV/PeV study did not use NP as a reference, so it does not contribute to the NP-referenced bar plot and is only relevant for select forest plot comparisons (**S3 Fig**).

Specificity estimates were uniformly high across all three specimen pools and all viruses, as shown in **Fig 4**. Pooled specificity was 0.99 for both nasal and oral specimens and 0.98 for LRT specimens when compared against NP as a reference. Unlike sensitivity, there was no meaningful separation between specimen types in terms of specificity, and confidence intervals were generally narrow, reflecting consistent and reliable specificity across nasal, oral, and LRT sampling relative to NP.

HSROC curves were generated in order to collate pooled sensitivity and specificity diagnostic accuracy data for respiratory specimen groupings against an NP reference across all included respiratory viruses (**Fig 5**). According to the AUC values of these models, all three specimen groupings demonstrated strong overall discrimination. Nasal specimens performed the best against NP specimens, with an AUC of 0.978, but oral and LRT specimen AUCs suggest comparable discrimination (oral AUC = 0.966, LRT AUC = 0.922). The NPA specimen had an overall AUC of 0.971 against the NP reference. NPA, nasal, and oral specimens had correspondingly high DORs, all exceeding 100. However, the LRT specimen had a DOR of 34.482; while still high, this is significantly less than the other comparisons. The same can be seen in the calculated measures of accuracy, including the Λ value and the Q* index, which represents the point on the curve where sensitivity equals specificity. The LRT Λ value (3.603) and Q* index (0.854) are both distinctively lower than the other comparison categories. These statistics are listed for each HSROC curve in **Table 4**.

**Fig 5.**
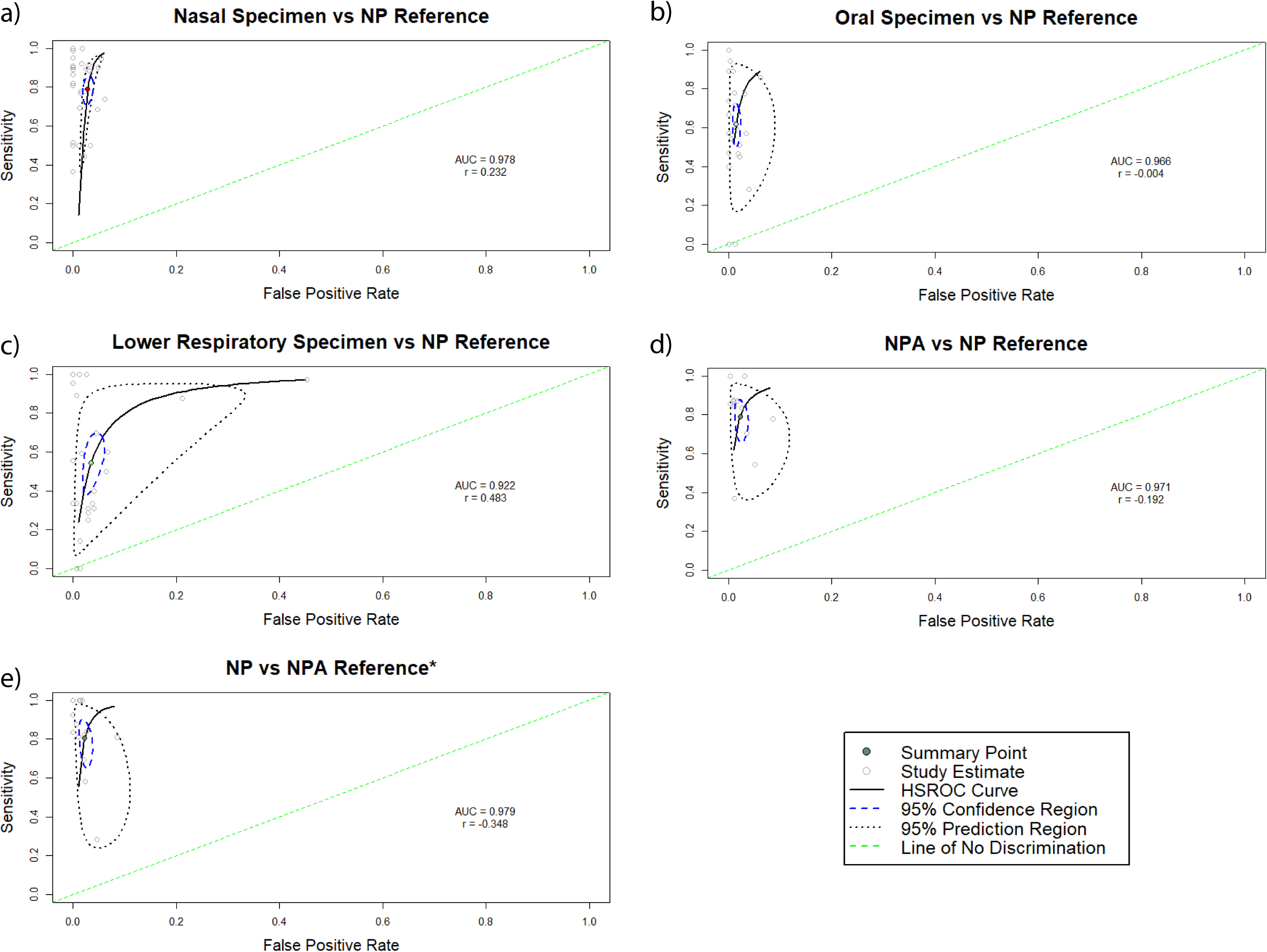
Viral Diagnostic Accuracy of Respiratory Specimen Groupings Against NP Reference. HSROC curves plotting sensitivity vs (1 - specificity), or false positive rate, of respiratory specimens against an NP reference are shown, across all respiratory viruses included in the analysis, i.e. RSV, ADV, hMPV, HCoV, INF, PIV, and RV/EV. Respiratory specimens are categorized as (A) nasal specimens, including ANS, MT, and RA, (B) oral specimens, including OP and saliva, and (C) lower respiratory specimens, including BAL, ETTA, TA, and IS. For comparison between NP and NPA, HSROC curves are included of both (D) NPA against an NP reference and (E) NP against an NPA reference. Pooled point estimates are colored points. A 95% confidence region is indicated by a blue dashed circle and a 95% prediction region by a larger black dotted circle. Gray points represent the distribution of point estimates from all data sets corresponding to the respective curve. AUC and correlation coefficient (r) values are listed for each plot. Abbreviations: Respiratory syncytial viruses (RSV); Adenovirus (ADV); Human metapneumovirus (hMPV); Human coronaviruses (HCoV); Influenza (INF); Parainfluenza viruses (PIV); Rhinovirus (RV); Enteroviruses (EV); Nasopharyngeal swab (NP); Mid-turbinate swab (MT); Anterior nasal swab (ANS); Rhinorrhea (RA); Oropharyngeal swab (OP); Bronchoalveolar lavage fluid (BAL); Endotracheal tube aspirate (ETTA); Tracheal aspirate (TA); Induced sputum (IS); Area under the curve (AUC)

**Table 4.**
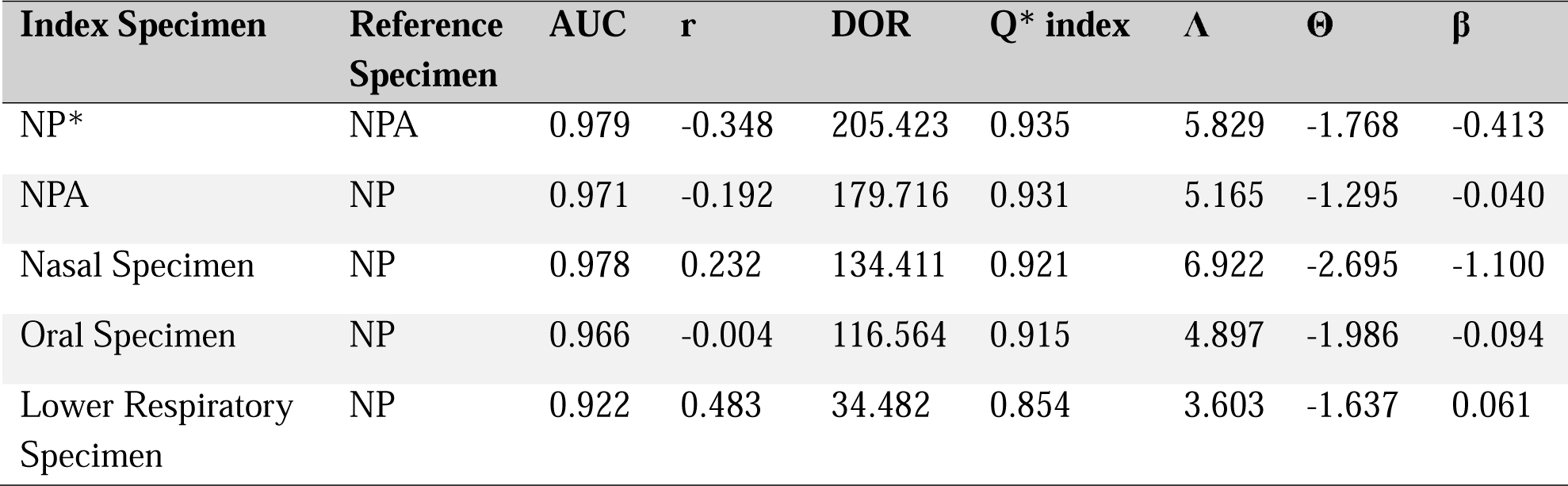
Measures of accuracy, heterogeneity, and shape across the Hierarchical Summary Receiver Operating Characteristic (HSROC) analyses. Reported statistics include the Area Under the Curve (AUC), correlation coefficient (r), Diagnostic Odds Ratio (DOR), Q* index, Accuracy (Λ), threshold (Θ), and shape (β). Q* index is the value on the curve at which specificity and sensitivity are equal. All analyses are conducted against an NP reference except for the first entry (*), which compares data from a nasopharyngeal swab (NP) index against a nasopharyngeal aspirate reference.

Furthermore, differences can be seen in curve shape and variability. The LRT comparison has a conspicuously broader spectrum of study estimates than the nasal and oral groupings, with a much larger 95% prediction region. Meanwhile, the nasal specimen curve is visibly skewed. These observations are reflected in the curves’ r and β values. While several of the comparison groups have weakly negative correlation coefficients, the correlation coefficients for nasal specimen (r = 0.232) and LRT specimen (r = 0.483) are notably positive, deviating from expectation. Additionally, the majority of the curves have β values close to 0, indicating symmetry. The exception is the nasal specimen plot (β = -1.100).

### Specimen collection comfort and preferences

Of the 36 studies included in the meta-analysis, 7 studies, including 912 participants, examined participants’ experiences with different sampling methods, as summarized in **Data 5** in **S2 Dataset**. Of these, 4 studies reported aggregated preference data, while 3 studies reported Likert-scale data, comparing NP against NS or saliva, and NS+TS against RA (nasal secretion) swabs (**Table 5**). In general, the gold-standard nasopharyngeal swabs were consistently ranked poorly for tolerability. Where participants graded discomfort on a scale, NP discomfort was rated as high [27, 34, 35], and where preference was indicated, NP was the least preferred sample type, except when compared to nasal wash [30, 32, 36, 37]. NS were typically well tolerated and ranked highly for preference. The preference for NS was reduced when combined with a TS [30, 32, 34], but throat swabbing alone was not included in any head-to-head analyses, and data on combined sampling was very limited. Saliva was the best-tolerated and most preferred sample type, outperforming NS, NS+TS and NP swabs in the two studies where it was included [27, 32]. Additionally, the collection of nasal secretions from RA using a swab was better tolerated than combined NS+TS [34] highlighting the potential of novel, minimally invasive collection methods.

**Table 5.** Ranking of the perceived discomfort of respiratory sample collection from studies that provided head-to-head ordinal data. Data are an aggregated, n-weighted average of ratings from patients, caregivers, and healthcare professionals.

|  | Aggregated n-weighted discomfort (0 = best) | N, total respondents |
| --- | --- | --- |
| NS | 3.3 | 247 |
| S | 7.2 | 96 |
| RS | 7.1 | 176 |
| NP | 16.7 | 154 |
| NS+OP | 39.3 | 176 |

Abbreviations: Nasal swab (NS); Saliva (S); Rhinorrhea swab (RS); Nasopharyngeal swab (NP); Oropharyngeal swab (OP).

## Discussion

The key focus of this meta-analysis was to establish per-virus sensitivity and specificity measures for various sample types, specifically in pediatric populations, to better inform future diagnostic and surveillance efforts. We found that, compared with nasopharyngeal specimens, nasal specimens (MT and ANS) had the highest sensitivity across all viruses. When grouped by anatomical site, where the samples were grouped (nasal, oral, or LRT) and stratified by virus, nasal samples again generally provided the highest sensitivity, with oral and LRT sensitivities generally lower and varying more by virus.

Specificity was overall high across all sample types compared with NP. When compared with existing recommendations (**Table 1**), which typically favor nasal, NP, or combined NP/OP swabs, our data generally supports the use of these samples in pediatric settings, though we did not have sufficient data to include combined sample types in our analysis. All data on combined specimen samples from the studies in our meta-analysis can be found in the forest plots in **S3 Fig**, within the plots for their respective viruses.

Because many viruses and specimen types were represented by only a few head-to-head studies, related specimen types were pooled into broader nasal, oral, and LRT categories to increase statistical power [9]. Pooling increased the number of studies contributing to each comparison, limiting the influence of heterogeneity between studies and narrowing variability relative to estimating the accuracy for each individual specimen type separately [38]. This approach has been used previously in a similar meta-analysis [39].

HSROC analysis of this grouped sample sensitivity and specificity data yielded high AUCs for all three categories (nasal, oral, and LRT specimens), suggesting good performance relative to NP as the reference. Of these, LRT specimens seem to have the lowest AUC and only produced moderately good DOR and accuracy values compared to the high DOR and accuracy of the other specimen types. In combination with the higher variability, large prediction region, and atypically positive correlation coefficient of the LRT data, this is likely a result of low study count per LRT subgroup and low sample populations within LRT studies, indicating a need for further research on the diagnostic power of lower respiratory specimens in comparison to the gold standard, NPS.

While grouping by sampling type improved our statistical power, it also risks flattening the specific niches of particular pathogens. However, in real clinical practice, the distinction between specific niches, for example, an anterior nares vs mid turbinate swab, is not likely to be meaningful in very young patients. Where the flattening effect of these groupings is likely to be most impactful is in the group of oral/throat specimens, including OP or throat swabs alongside saliva, which are known, for example, to host markedly different bacterial communities [40]. Similarly, the differences between, for example, induced sputum and bronchoalveolar lavage are stark. Differences in LRT sampling may reflect both true sampling depth and the clinically driven context of collection, so pooling LRT specimens likely contributes to the particularly wide confidence intervals observed for several viruses.

However, when considering the diagnostic accuracy of LRT samples, comparing them with URT samples implies a somewhat fictional sense of choice. All but one study (which included IS) using LRT samples were conducted in hospitalized patients. It is unlikely that LRT specimen collection, which may be necessary in a pediatric intensive care unit, would be a clinician’s first choice in a community setting. This is reflected in the existing WHO and CDC recommendations (**Table 1**). However, we believe a head-to-head examination of diagnostic accuracy remains useful for providing an overall picture of what clinicians can expect from each sample type.

An additional consideration when selecting an ideal sample type, beyond diagnostic accuracy, is practical utility and tolerability. Across the 7 studies that assessed tolerability, saliva and nasal swabs were consistently well tolerated. These analyses were limited by the sparsity of head-to-head data, with some sample types being better represented than others. The level of invasiveness of sampling and the ability to self-collect may be of particular importance in certain clinical settings, for example, in children who cannot easily comply with being swabbed. Taken together, these data suggest that nasal swabs are generally well tolerated and perform well diagnostically compared with NP specimens, though more data are needed to support other well-tolerated sample types, such as saliva.

Our study has some limitations that are worth noting. Firstly, the number of studies included, 36, was relatively small. Further, the number of studies for which we could conduct in-depth comparative analyses was lower still (18 of the 36 included), as not all studies had standardized reference and index samples for such analyses. This limited the effective sample size for several virus-specimen comparisons and contributed to wide confidence intervals, particularly for LRT estimates.

To ensure methodological consistency, we recalculated all sensitivity and specificity measures extracted from each dataset, using the study’s stated reference as the gold standard. This assumes that the reference is inherently more sensitive than the index, treating any index positive as a false positive if the paired reference was negative, even if the original study used a consensus method, in which any sample that was positive was considered a true positive. This method, though necessary for comparing across studies, introduces a bias that favors the reference, which, for our analysis, was either NPS or NPA. The impact and inherent limitation of this can be seen in comparisons between NP and NPA, using one as a reference and then the other, where both have a sensitivity of approximately 85% relative to each other. However, the fact that all specimen types showed very high specificity indicates that the actual number of “false positives” as determined by our methodology, which may have been “true positives” using a consensus method, were low.

Finally, some of the datasets were extremely small. For example, Lin 2017 and Wurzel 2013 had only 3 and 4 positive reference HCOV results or “true positives” respectively, which necessarily reduces power, inflates variance, and exaggerates low sensitivity values (Lin et al., 2017; Wurzel et al., 2013). While our pooled random-effects analyses weighted the studies by the inverse of variance, correcting some of this effect, they do not consider n values directly and limited representativeness remains a concern. This contributed to wide confidence intervals in our analysis, reducing the statistical certainty of our conclusions.

Accurate and timely diagnosis of respiratory viral infections is essential for both guiding clinical intervention, including reducing the overprescription of antibiotics, and managing infection transmission, particularly in children [41]. Here, we examined the full available literature to determine the optimal respiratory specimen types for viral detection in pediatric settings. Our findings broadly support the existing recommendations, with nasal swabs performing well compared to nasopharyngeal specimens for the detection of a broad range of viruses and demonstrating good tolerability. Oral and LRT specimens also performed moderately well for specific viruses, though with much greater variability in general. This work, therefore, also highlights the need for further diagnostic accuracy and tolerability studies to draw more conclusive insights, especially regarding the performance of LRT specimens, for which data are very limited.

## Supporting information

PRISMA checklist

PRISMA abstract checklist

## Data Availability

All data produced in the present work are contained in the manuscript and supporting files.

## Abbreviations

NP: Nasopharyngeal swab
NPA: Nasopharyngeal aspirate/wash
OP: Oropharyngeal/throat swab
MT: Mi-turbinate swab
BAL: Bronchoalveolar lavage fluid
S: Saliva (drool)
ANS: Anterior nasal/nasal swab
ETTA: Endotracheal tube aspirate
IS: Induced sputum
TA: Tracheal aspirate
RA: Rhinorrhea
SARS-CoV-2: Severe acute respiratory syndrome coronavirus 2
HCoV: Human Coronaviruses (229E, OC43, NL63, HKU1)
RSV: Respiratory syncytial viruses
FLU: Influenza (includes influenza A and B)
PIV: Parainfluenza viruses 1-4
ADV: Adenovirus
RV: Rhinovirus
EV: Enteroviruses (respiratory subtypes)
PeV: Parechoviruses
hMPV: Human Metapneumovirus
BOV: Bocavirus

## Contributions

Conceptualization: OMA, AD, CSL, IY; Data Curation: OMA, AD, JHC, KR, HOH, BO, MCF, CSL; Formal Analysis: OMA, AD, JHC, KR, CSL; Investigation: OMA, AD, JHC, KR, CSL; Visualization: AD, JHC, KR; Project Administration: OMA, AD, CSL; Supervision: OMA, CSL, IY; Writing – Original Draft Preparation: OMA, AD, CSL, JHC, KR, IY; Writing – Review & Editing: all authors.

## Supporting Information Legends

***S1 Table. PICOTS eligibility criteria for studies included in this review.***

***S2 Dataset. Detailed supporting information regarding search strategy, data extraction, QUADAS-2 Risk of Bias and Applicability Assessment, and comfort and tolerability analysis.***

***S3 Fig. Forest plots of sensitivity and specificity across specimen types for each virus.** All data sets on each respective viTrus across included studies were used to visualize the relative and pooled (A) sensitivities and (B) specificities of RSV detection; (C) sensitivities and (D) specificities of ADV detection; (E) sensitivities and (F) specificities of hMPV detection; (G) sensitivities and (H) specificities of INF detection; (I) sensitivities and (J) specificities of PIV detection; and (K) sensitivities and (L) specificities of RV/EV/PeV detection. Data is subgrouped by reference test, and pooled estimates are weighted by inverse-variance.*

***S4 Fig. Forest plots of sensitivity and specificity of relevant specimen types against NP reference, without discrimination by virus.** Comparisons of NP and NPA can be found in the (A) sensitivity and (B) specificity plots of NP against an NPA reference, as well as the (C) sensitivity and (D) specificity plots of NPA against an NP reference. Pooled estimates against an NP reference across all viruses are calculated for the other common index specimen, including the (E) sensitivity and (F) specificity of MT, (G) sensitivity and (H) specificity of ANS, (I) sensitivity and (J) specificity of OP, and (K) sensitivity and (L) specificity of BAL. Pooled estimates are weighted by inverse-variance.*

***S5 Fig. Forest plots of sensitivity and specificity of grouped nasal, oral, and lower respiratory specimens against NP reference.** Data on the a) sensitivity and b) specificity of nasal specimens (ANS, MT, and RA), c) sensitivity and d) specificity of oral specimens (OP and saliva), and e) sensitivity and f) specificity of lower respiratory specimens (BAL, ETTA, TA, and IS) is subgrouped by respiratory virus. Pooled estimates are weighted by inverse-variance.*

