## Supplementary material for "One size fits all: A systematic review of the sample types used for the diagnostics of respiratory viruses in children": S1 Table

**S1 Table. PICOTS eligibility criteria for studies included in this review.**

| | Inclusion criteria | Exclusion criteria |
|---|---|---|
| Population | Children, defined as persons <18 years | Studies not reporting on patients with acute respiratory disease<br>Studies on adults or combined adult and pediatric populations without disaggregated results |
| Exposure | 1. Diagnostic testing specifications:<br>Index test and reference standard available<br>1.1. Specimen specifications:<br>1.2. Respiratory specimen types: NP specimen, OP specimen, nasal only specimen, saliva, blood, sputum, and other studied specimen types (alone and in combination, where possible)<br>1.3. Specimen collection methodology (swab, wash, BAL, others)<br>1.4. Number of specimens tested, per specimen type<br>2. Lab specifications<br>2.1. Laboratory storage and collection methods<br>2.2. RSV laboratory diagnostic techniques and procedures: viral culture, PCR (including all types), serology, antigen test<br>3. Disease specifications:<br>3.1. Clinical presentation of RSV<br>3.2. Time since symptom onset<br>3.3. Level of care for the RSV disease<br>3.4. RSV disease severity<br>3.5. RSV subtype | Respiratory specimen types: blood, feces (non-respiratory specimens) |
| Outcome | 1. Diagnostic test performance: Sensitivity, specificity, positive predictive value, and negative predictive value for the various levels of exposure<br>2. Quantitative comparison of testing methods:<br>Number or percentage of patients<br>with viral infection identified by one testing method versus number or percentage identified by a different testing method in the same population (examples include RT-PCR of NP swab vs sputum, or use of one swab type versus another) | Studies reporting none of the outcomes of interest |
| Time/Period of publication | 2000 - 2025 | Studies published before 2000 |
| Study design | Observational studies: prospective and retrospective cohort studies, prospective and retrospective cross-sectional studies, case control studies, ecological studies, diagnostic tests, randomized controlled clinical trials | Peer-reviewed publications that do not clearly outline methods and sources for data collection and analysis |
| Publication type | Published articles, conference posters. | In vitro studies<br>News and opinion articles<br>Case reports<br>Narrative reviews, letters<br>SLRs and meta-analysis (except for use to identify additional articles) |
| Language | English | Other languages |
