## Supplementary material for "One size fits all: A systematic review of the sample types used for the diagnostics of respiratory viruses in children": S3 Figure

a)

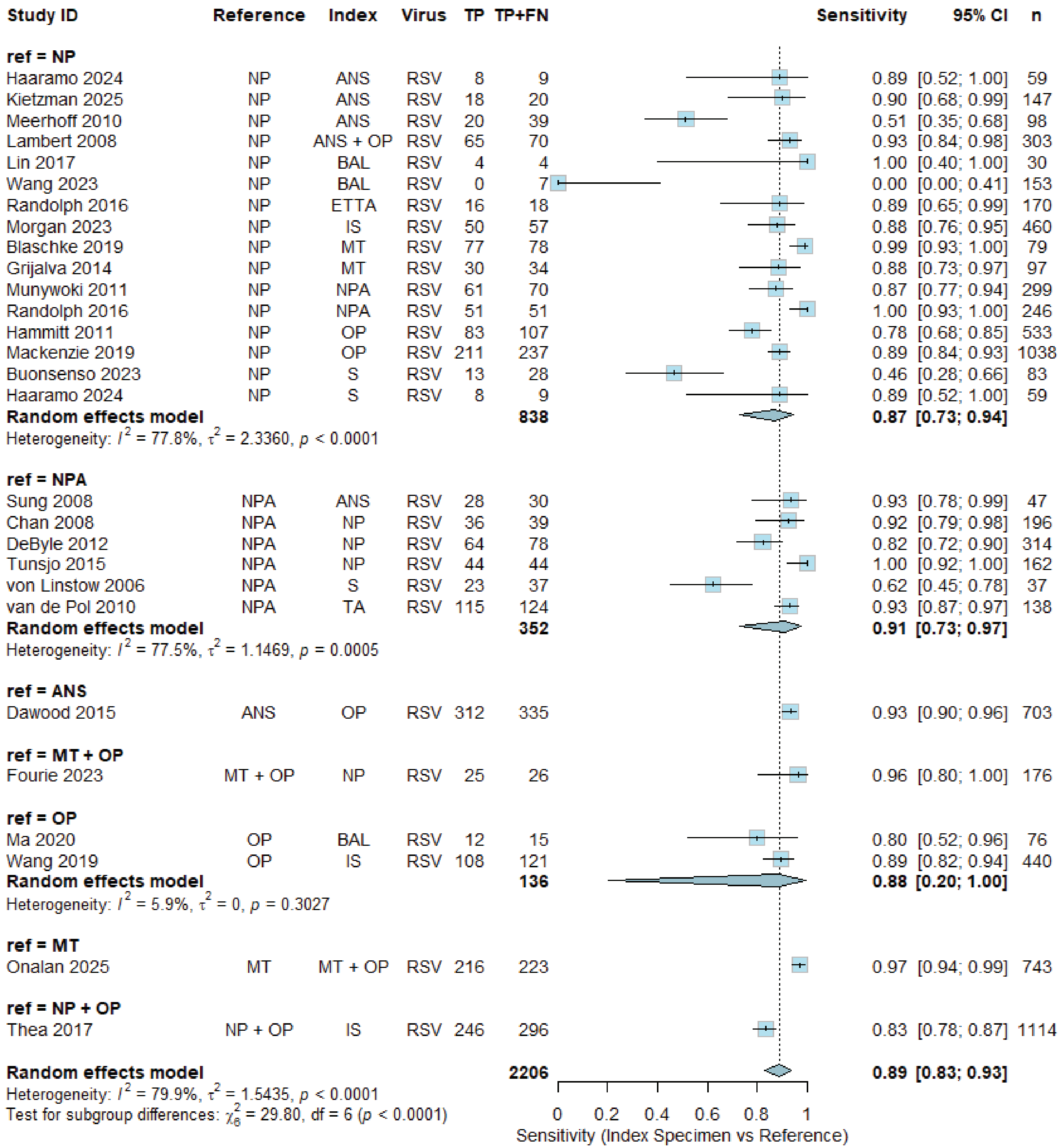

Study ID Reference Index Virus TP TP+FN Sensitivity 95% CI n
ref = NP
Haaramo 2024 NP ANS RSV 8 9 0.89 [0.52; 1.00] 59
Kietzman 2025 NP ANS RSV 18 20 0.90 [0.68; 0.99] 147
Meerhoff 2010 NP ANS RSV 20 39 0.51 [0.35; 0.68] 98
Lambert 2008 NP ANS + OP RSV 65 70 0.93 [0.84; 0.98] 303
Lin 2017 NP BAL RSV 4 4 1.00 [0.40; 1.00] 30
Wang 2023 NP BAL RSV 0 7 0.00 [0.00; 0.41] 153
Randolph 2016 NP ETTA RSV 16 18 0.89 [0.65; 0.99] 170
Morgan 2023 NP IS RSV 50 57 0.88 [0.76; 0.95] 460
Blaschke 2019 NP MT RSV 77 78 0.99 [0.93; 1.00] 79
Grijalva 2014 NP MT RSV 30 34 0.88 [0.73; 0.97] 97
Munywoki 2011 NP NPA RSV 61 70 0.87 [0.77; 0.94] 299
Randolph 2016 NP NPA RSV 51 51 1.00 [0.93; 1.00] 246
Hammitt 2011 NP OP RSV 83 107 0.78 [0.68; 0.85] 533
Mackenzie 2019 NP OP RSV 211 237 0.89 [0.84; 0.93] 1038
Buonsenso 2023 NP S RSV 13 28 0.46 [0.28; 0.66] 83
Haaramo 2024 NP S RSV 8 9 0.89 [0.52; 1.00] 59
Random effects model 838 0.87 [0.73; 0.94]
Heterogeneity: $I^2$ = 77.8%, $\tau^2$ = 2.3360, $p$ < 0.0001
ref = NPA
Sung 2008 NPA ANS RSV 28 30 0.93 [0.78; 0.99] 47
Chan 2008 NPA NP RSV 36 39 0.92 [0.79; 0.98] 196
DeByle 2012 NPA NP RSV 64 78 0.82 [0.72; 0.90] 314
Tunsjo 2015 NPA NP RSV 44 44 1.00 [0.92; 1.00] 162
von Linstow 2006 NPA S RSV 23 37 0.62 [0.45; 0.78] 37
van de Pol 2010 NPA TA RSV 115 124 0.93 [0.87; 0.97] 138
Random effects model 352 0.91 [0.73; 0.97]
Heterogeneity: $I^2$ = 77.5%, $\tau^2$ = 1.1469, $p$ = 0.0005
ref = ANS
Dawood 2015 ANS OP RSV 312 335 0.93 [0.90; 0.96] 703
ref = MT + OP
Fourie 2023 MT + OP NP RSV 25 26 0.96 [0.80; 1.00] 176
ref = OP
Ma 2020 OP BAL RSV 12 15 0.80 [0.52; 0.96] 76
Wang 2019 OP IS RSV 108 121 0.89 [0.82; 0.94] 440
Random effects model 136 0.88 [0.20; 1.00]
Heterogeneity: $I^2$ = 5.9%, $\tau^2$ = 0, $p$ = 0.3027
ref = MT
Onalan 2025 MT MT + OP RSV 216 223 0.97 [0.94; 0.99] 743
ref = NP + OP
Thea 2017 NP + OP IS RSV 246 296 0.83 [0.78; 0.87] 1114
Random effects model 2206 0.89 [0.83; 0.93]
Heterogeneity: $I^2$ = 79.9%, $\tau^2$ = 1.5435, $p$ < 0.0001
Test for subgroup differences: $\chi^2_6$ = 29.80, df = 6 ($p$ < 0.0001)
0 0.2 0.4 0.6 0.8 1
Sensitivity (Index Specimen vs Reference)

b)

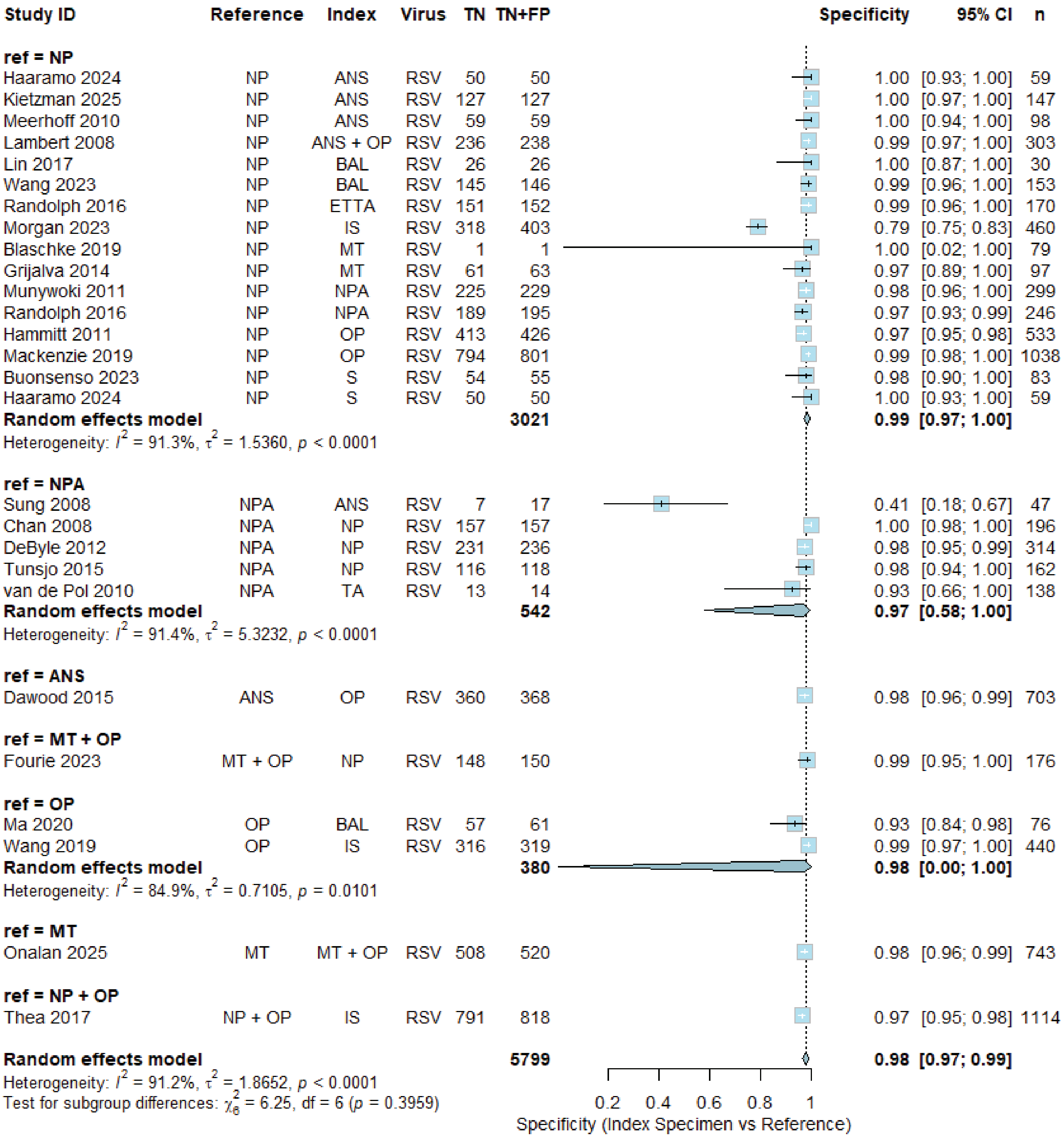
Study ID Reference Index Virus TN TN+FP Specificity 95% CI n
ref = NP
Haaramo 2024 NP ANS RSV 50 50 1.00 [0.93; 1.00] 59
Kietzman 2025 NP ANS RSV 127 127 1.00 [0.97; 1.00] 147
Meerhoff 2010 NP ANS RSV 59 59 1.00 [0.94; 1.00] 98
Lambert 2008 NP ANS + OP RSV 236 238 0.99 [0.97; 1.00] 303
Lin 2017 NP BAL RSV 26 26 1.00 [0.87; 1.00] 30
Wang 2023 NP BAL RSV 145 146 0.99 [0.96; 1.00] 153
Randolph 2016 NP ETTA RSV 151 152 0.99 [0.96; 1.00] 170
Morgan 2023 NP IS RSV 318 403 0.79 [0.75; 0.83] 460
Blaschke 2019 NP MT RSV 1 1 1.00 [0.02; 1.00] 79
Grijalva 2014 NP MT RSV 61 63 0.97 [0.89; 1.00] 97
Munywoki 2011 NP NPA RSV 225 229 0.98 [0.96; 1.00] 299
Randolph 2016 NP NPA RSV 189 195 0.97 [0.93; 0.99] 246
Hammitt 2011 NP OP RSV 413 426 0.97 [0.95; 0.98] 533
Mackenzie 2019 NP OP RSV 794 801 0.99 [0.98; 1.00] 1038
Buonsenso 2023 NP S RSV 54 55 0.98 [0.90; 1.00] 83
Haaramo 2024 NP S RSV 50 50 1.00 [0.93; 1.00] 59
Random effects model 3021 0.99 [0.97; 1.00]
Heterogeneity: $I^2$ = 91.3%, $\tau^2$ = 1.5360, $p$ < 0.0001
ref = NPA
Sung 2008 NPA ANS RSV 7 17 0.41 [0.18; 0.67] 47
Chan 2008 NPA NP RSV 157 157 1.00 [0.98; 1.00] 196
DeByle 2012 NPA NP RSV 231 236 0.98 [0.95; 0.99] 314
Tunsjo 2015 NPA NP RSV 116 118 0.98 [0.94; 1.00] 162
van de Pol 2010 NPA TA RSV 13 14 0.93 [0.66; 1.00] 138
Random effects model 542 0.97 [0.58; 1.00]
Heterogeneity: $I^2$ = 91.4%, $\tau^2$ = 5.3232, $p$ < 0.0001
ref = ANS
Dawood 2015 ANS OP RSV 360 368 0.98 [0.96; 0.99] 703
ref = MT + OP
Fourie 2023 MT + OP NP RSV 148 150 0.99 [0.95; 1.00] 176
ref = OP
Ma 2020 OP BAL RSV 57 61 0.93 [0.84; 0.98] 76
Wang 2019 OP IS RSV 316 319 0.99 [0.97; 1.00] 440
Random effects model 380 0.98 [0.00; 1.00]
Heterogeneity: $I^2$ = 84.9%, $\tau^2$ = 0.7105, $p$ = 0.0101
ref = MT
Onalan 2025 MT MT + OP RSV 508 520 0.98 [0.96; 0.99] 743
ref = NP + OP
Thea 2017 NP + OP IS RSV 791 818 0.97 [0.95; 0.98] 1114
Random effects model 5799 0.98 [0.97; 0.99]
Heterogeneity: $I^2$ = 91.2%, $\tau^2$ = 1.8652, $p$ < 0.0001
Test for subgroup differences: $\chi^2_6$ = 6.25, df = 6 ($p$ = 0.3959)
0.2 0.4 0.6 0.8 1
Specificity (Index Specimen vs Reference)

c)

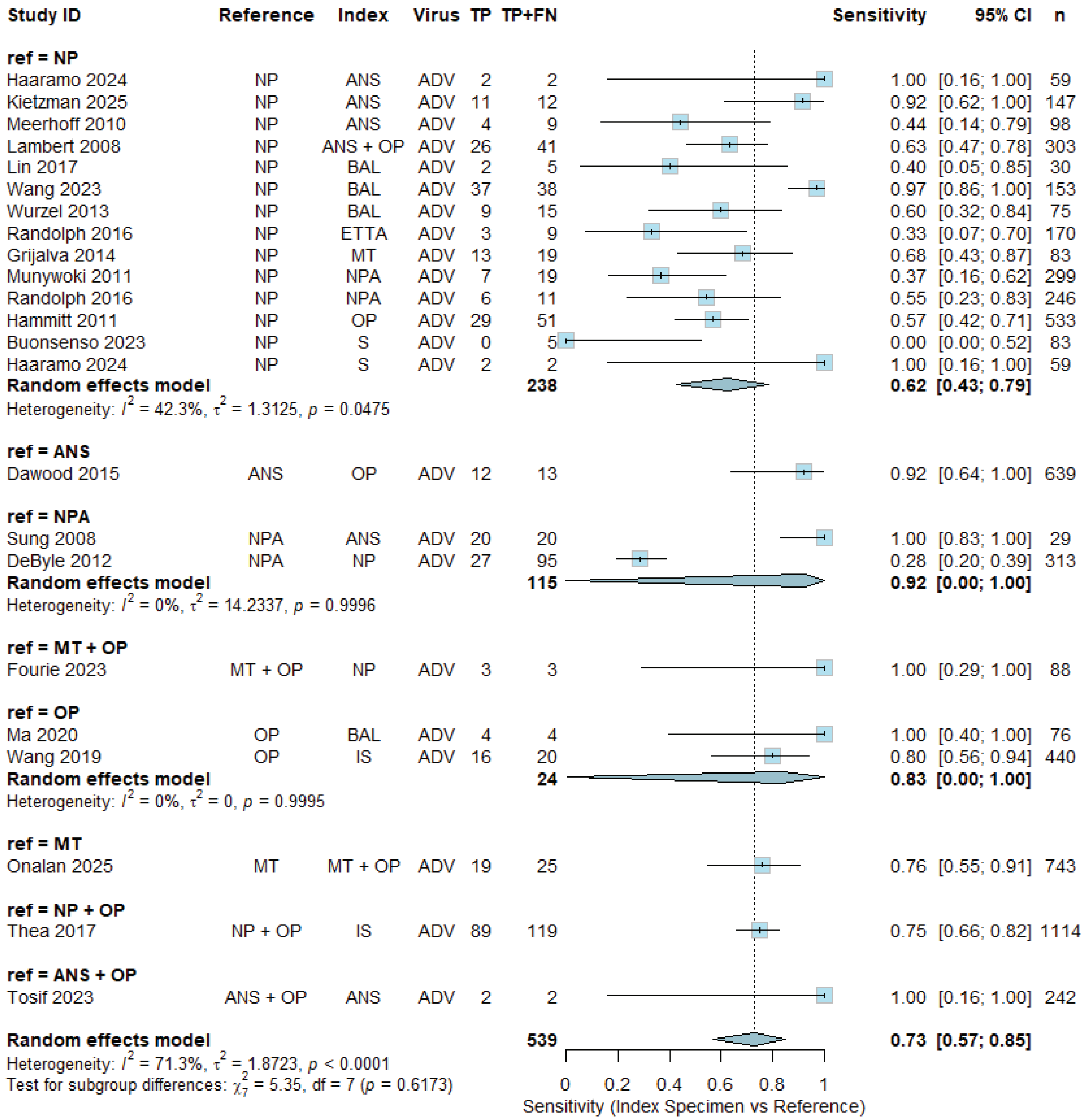
Study ID Reference Index Virus TP TP+FN Sensitivity 95% CI n
ref = NP
Haaramo 2024 NP ANS ADV 2 2 1.00 [0.16; 1.00] 59
Kietzman 2025 NP ANS ADV 11 12 0.92 [0.62; 1.00] 147
Meerhoff 2010 NP ANS ADV 4 9 0.44 [0.14; 0.79] 98
Lambert 2008 NP ANS + OP ADV 26 41 0.63 [0.47; 0.78] 303
Lin 2017 NP BAL ADV 2 5 0.40 [0.05; 0.85] 30
Wang 2023 NP BAL ADV 37 38 0.97 [0.86; 1.00] 153
Wurzel 2013 NP BAL ADV 9 15 0.60 [0.32; 0.84] 75
Randolph 2016 NP ETTA ADV 3 9 0.33 [0.07; 0.70] 170
Grijalva 2014 NP MT ADV 13 19 0.68 [0.43; 0.87] 83
Munywoki 2011 NP NPA ADV 7 19 0.37 [0.16; 0.62] 299
Randolph 2016 NP NPA ADV 6 11 0.55 [0.23; 0.83] 246
Hammitt 2011 NP OP ADV 29 51 0.57 [0.42; 0.71] 533
Buonsenso 2023 NP S ADV 0 5 0.00 [0.00; 0.52] 83
Haaramo 2024 NP S ADV 2 2 1.00 [0.16; 1.00] 59
Random effects model 238 0.62 [0.43; 0.79]
Heterogeneity: $I^2$ = 42.3%, $\tau^2$ = 1.3125, $p$ = 0.0475
ref = ANS
Dawood 2015 ANS OP ADV 12 13 0.92 [0.64; 1.00] 639
ref = NPA
Sung 2008 NPA ANS ADV 20 20 1.00 [0.83; 1.00] 29
DeByle 2012 NPA NP ADV 27 95 0.28 [0.20; 0.39] 313
Random effects model 115 0.92 [0.00; 1.00]
Heterogeneity: $I^2$ = 0%, $\tau^2$ = 14.2337, $p$ = 0.9996
ref = MT + OP
Fourie 2023 MT + OP NP ADV 3 3 1.00 [0.29; 1.00] 88
ref = OP
Ma 2020 OP BAL ADV 4 4 1.00 [0.40; 1.00] 76
Wang 2019 OP IS ADV 16 20 0.80 [0.56; 0.94] 440
Random effects model 24 0.83 [0.00; 1.00]
Heterogeneity: $I^2$ = 0%, $\tau^2$ = 0, $p$ = 0.9995
ref = MT
Onalan 2025 MT MT + OP ADV 19 25 0.76 [0.55; 0.91] 743
ref = NP + OP
Thea 2017 NP + OP IS ADV 89 119 0.75 [0.66; 0.82] 1114
ref = ANS + OP
Tosif 2023 ANS + OP ANS ADV 2 2 1.00 [0.16; 1.00] 242
Random effects model 539 0.73 [0.57; 0.85]
Heterogeneity: $I^2$ = 71.3%, $\tau^2$ = 1.8723, $p$ < 0.0001
Test for subgroup differences: $\chi^2_7$ = 5.35, df = 7 ($p$ = 0.6173)
0 0.2 0.4 0.6 0.8 1
Sensitivity (Index Specimen vs Reference)

d)

| Study ID | Reference | Index | Virus | TN | TN+FP | Specificity | 95% CI | n |
|---|---|---|---|---|---|---|---|---|
| **ref = NP** | | | | | | | | |
| Haaramo 2024 | NP | ANS | ADV | 56 | 57 | 0.98 | [0.91; 1.00] | 59 |
| Kietzman 2025 | NP | ANS | ADV | 131 | 135 | 0.97 | [0.93; 0.99] | 147 |
| Meerhoff 2010 | NP | ANS | ADV | 87 | 89 | 0.98 | [0.92; 1.00] | 98 |
| Lambert 2008 | NP | ANS + OP | ADV | 277 | 280 | 0.99 | [0.97; 1.00] | 303 |
| Lin 2017 | NP | BAL | ADV | 24 | 25 | 0.96 | [0.80; 1.00] | 30 |
| Wang 2023 | NP | BAL | ADV | 63 | 115 | 0.55 | [0.45; 0.64] | 153 |
| Wurzel 2013 | NP | BAL | ADV | 56 | 60 | 0.93 | [0.84; 0.98] | 75 |
| Randolph 2016 | NP | ETTA | ADV | 161 | 161 | 1.00 | [0.98; 1.00] | 170 |
| Grijalva 2014 | NP | MT | ADV | 61 | 64 | 0.95 | [0.87; 0.99] | 83 |
| Munywoki 2011 | NP | NPA | ADV | 277 | 280 | 0.99 | [0.97; 1.00] | 299 |
| Randolph 2016 | NP | NPA | ADV | 223 | 235 | 0.95 | [0.91; 0.97] | 246 |
| Hammitt 2011 | NP | OP | ADV | 466 | 482 | 0.97 | [0.95; 0.98] | 533 |
| Buonsenso 2023 | NP | S | ADV | 77 | 78 | 0.99 | [0.93; 1.00] | 83 |
| Haaramo 2024 | NP | S | ADV | 57 | 57 | 1.00 | [0.94; 1.00] | 59 |
| **Random effects model** | | | | | **2118** | **0.98** | **[0.94; 0.99]** | |
| Heterogeneity: $I^2 = 93.4\%$, $\tau^2 = 1.7955$, $p < 0.0001$ | | | | | | | | |
| **ref = ANS** | | | | | | | | |
| Dawood 2015 | ANS | OP | ADV | 625 | 626 | 1.00 | [0.99; 1.00] | 639 |
| **ref = NPA** | | | | | | | | |
| Sung 2008 | NPA | ANS | ADV | 4 | 9 | 0.44 | [0.14; 0.79] | 29 |
| DeByle 2012 | NPA | NP | ADV | 208 | 218 | 0.95 | [0.92; 0.98] | 313 |
| **Random effects model** | | | | | **227** | **0.82** | **[0.00; 1.00]** | |
| Heterogeneity: $I^2 = 94.8\%$, $\tau^2 = 2.4520$, $p < 0.0001$ | | | | | | | | |
| **ref = MT + OP** | | | | | | | | |
| Fourie 2023 | MT + OP | NP | ADV | 81 | 85 | 0.95 | [0.88; 0.99] | 88 |
| **ref = OP** | | | | | | | | |
| Ma 2020 | OP | BAL | ADV | 69 | 72 | 0.96 | [0.88; 0.99] | 76 |
| Wang 2019 | OP | IS | ADV | 417 | 420 | 0.99 | [0.98; 1.00] | 440 |
| **Random effects model** | | | | | **492** | **0.99** | **[0.02; 1.00]** | |
| Heterogeneity: $I^2 = 78.9\%$, $\tau^2 = 0.4457$, $p = 0.0296$ | | | | | | | | |
| **ref = MT** | | | | | | | | |
| Onalan 2025 | MT | MT + OP | ADV | 702 | 718 | 0.98 | [0.96; 0.99] | 743 |
| **ref = NP + OP** | | | | | | | | |
| Thea 2017 | NP + OP | IS | ADV | 941 | 995 | 0.95 | [0.93; 0.96] | 1114 |
| **ref = ANS + OP** | | | | | | | | |
| Tosif 2023 | ANS + OP | ANS | ADV | 239 | 240 | 1.00 | [0.98; 1.00] | 242 |
| **Random effects model** | | | | | **5501** | **0.98** | **[0.95; 0.99]** | |
| Heterogeneity: $I^2 = 92.5\%$, $\tau^2 = 2.1564$, $p < 0.0001$ | | | | | | | | |
| Test for subgroup differences: $\chi^2_7 = 32.78$, df = 7 ($p < 0.0001$) | | | | | | | | |

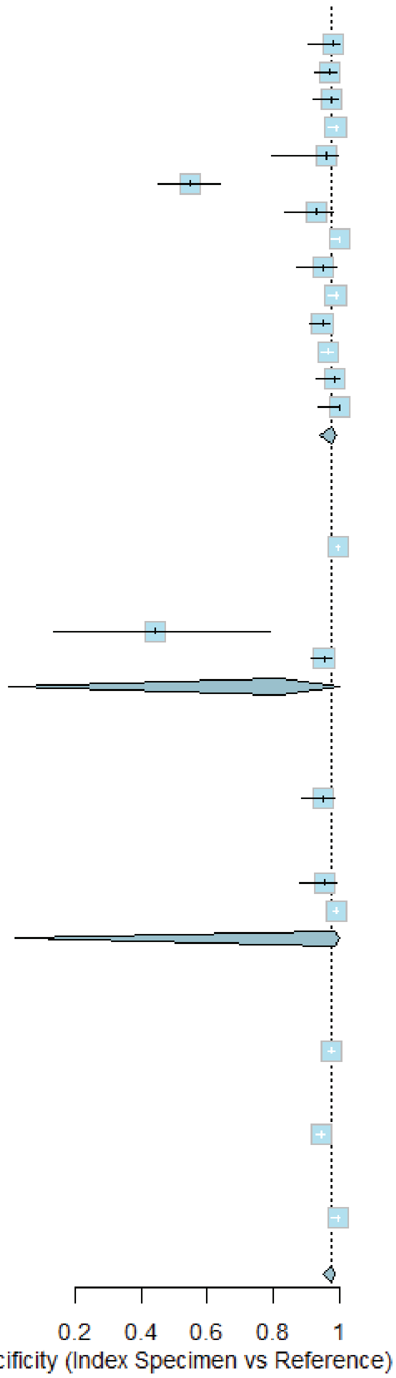

e)

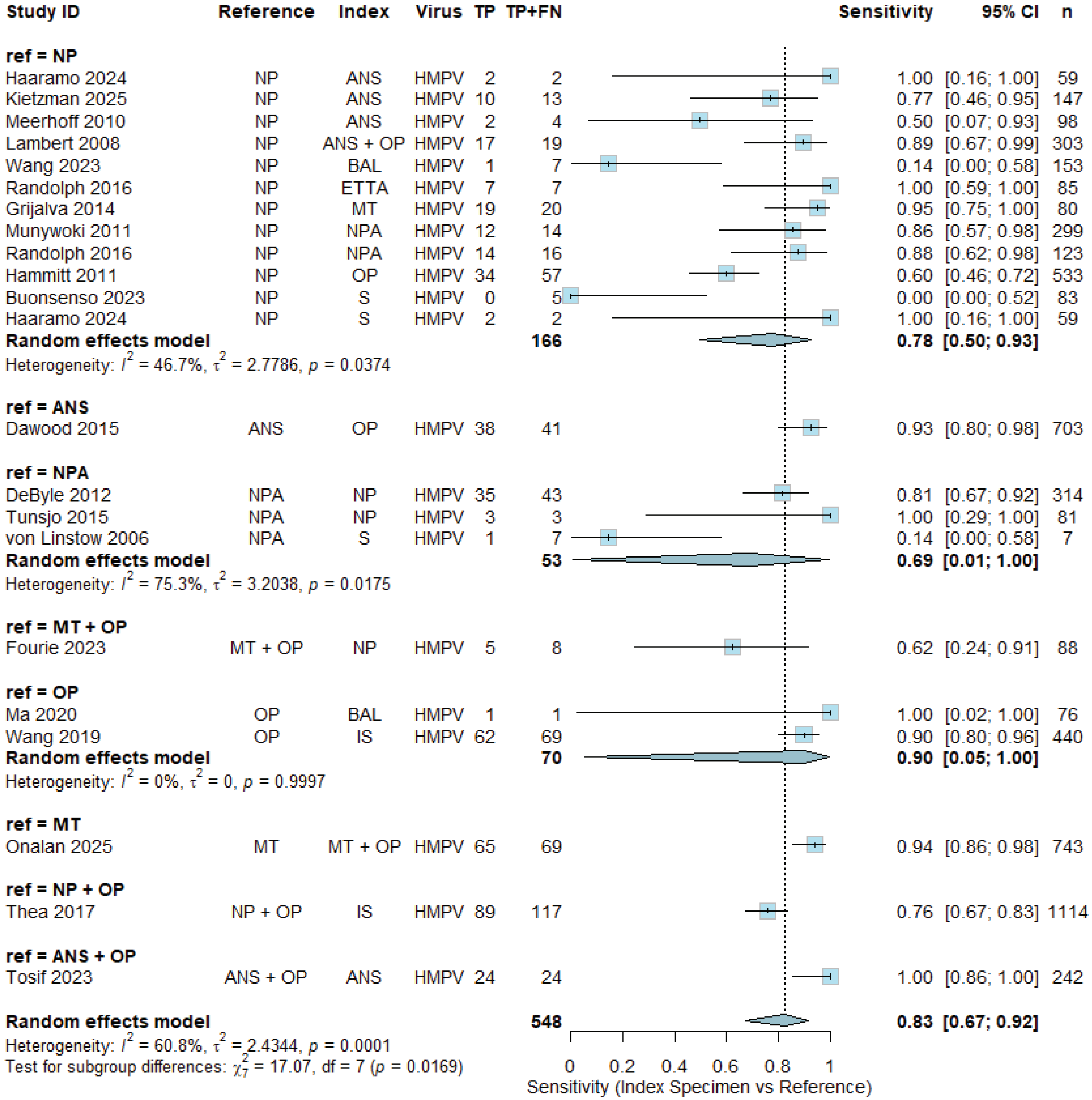

Study ID Reference Index Virus TP TP+FN Sensitivity 95% CI n
ref = NP
Haaramo 2024 NP ANS HMPV 2 2 1.00 [0.16; 1.00] 59
Kietzman 2025 NP ANS HMPV 10 13 0.77 [0.46; 0.95] 147
Meerhoff 2010 NP ANS HMPV 2 4 0.50 [0.07; 0.93] 98
Lambert 2008 NP ANS + OP HMPV 17 19 0.89 [0.67; 0.99] 303
Wang 2023 NP BAL HMPV 1 7 0.14 [0.00; 0.58] 153
Randolph 2016 NP ETTA HMPV 7 7 1.00 [0.59; 1.00] 85
Grijalva 2014 NP MT HMPV 19 20 0.95 [0.75; 1.00] 80
Munywoki 2011 NP NPA HMPV 12 14 0.86 [0.57; 0.98] 299
Randolph 2016 NP NPA HMPV 14 16 0.88 [0.62; 0.98] 123
Hammitt 2011 NP OP HMPV 34 57 0.60 [0.46; 0.72] 533
Buonsenso 2023 NP S HMPV 0 5 0.00 [0.00; 0.52] 83
Haaramo 2024 NP S HMPV 2 2 1.00 [0.16; 1.00] 59
Random effects model 166 0.78 [0.50; 0.93]
Heterogeneity: $I^2$ = 46.7%, $\tau^2$ = 2.7786, $p$ = 0.0374
ref = ANS
Dawood 2015 ANS OP HMPV 38 41 0.93 [0.80; 0.98] 703
ref = NPA
DeByle 2012 NPA NP HMPV 35 43 0.81 [0.67; 0.92] 314
Tunsjo 2015 NPA NP HMPV 3 3 1.00 [0.29; 1.00] 81
von Linstow 2006 NPA S HMPV 1 7 0.14 [0.00; 0.58] 7
Random effects model 53 0.69 [0.01; 1.00]
Heterogeneity: $I^2$ = 75.3%, $\tau^2$ = 3.2038, $p$ = 0.0175
ref = MT + OP
Fourie 2023 MT + OP NP HMPV 5 8 0.62 [0.24; 0.91] 88
ref = OP
Ma 2020 OP BAL HMPV 1 1 1.00 [0.02; 1.00] 76
Wang 2019 OP IS HMPV 62 69 0.90 [0.80; 0.96] 440
Random effects model 70 0.90 [0.05; 1.00]
Heterogeneity: $I^2$ = 0%, $\tau^2$ = 0, $p$ = 0.9997
ref = MT
Onalan 2025 MT MT + OP HMPV 65 69 0.94 [0.86; 0.98] 743
ref = NP + OP
Thea 2017 NP + OP IS HMPV 89 117 0.76 [0.67; 0.83] 1114
ref = ANS + OP
Tosif 2023 ANS + OP ANS HMPV 24 24 1.00 [0.86; 1.00] 242
Random effects model 548 0.83 [0.67; 0.92]
Heterogeneity: $I^2$ = 60.8%, $\tau^2$ = 2.4344, $p$ = 0.0001
Test for subgroup differences: $\chi^2_7$ = 17.07, df = 7 ($p$ = 0.0169)
0 0.2 0.4 0.6 0.8 1
Sensitivity (Index Specimen vs Reference)

f)

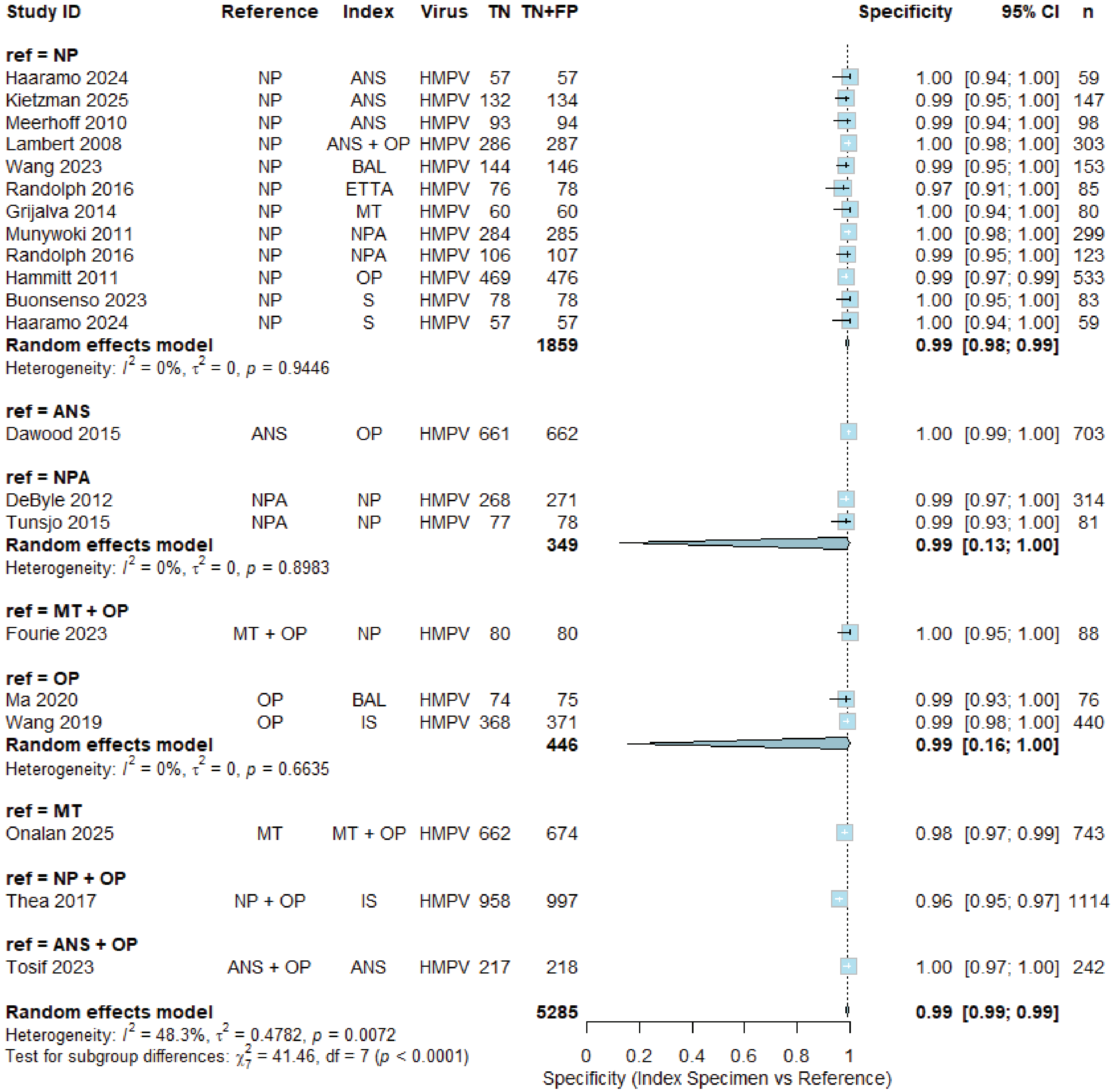

Study ID Reference Index Virus TN TN+FP Specificity 95% CI n
ref = NP
Haaramo 2024 NP ANS HMPV 57 57 1.00 [0.94; 1.00] 59
Kietzman 2025 NP ANS HMPV 132 134 0.99 [0.95; 1.00] 147
Meerhoff 2010 NP ANS HMPV 93 94 0.99 [0.94; 1.00] 98
Lambert 2008 NP ANS + OP HMPV 286 287 1.00 [0.98; 1.00] 303
Wang 2023 NP BAL HMPV 144 146 0.99 [0.95; 1.00] 153
Randolph 2016 NP ETTA HMPV 76 78 0.97 [0.91; 1.00] 85
Grijalva 2014 NP MT HMPV 60 60 1.00 [0.94; 1.00] 80
Munywoki 2011 NP NPA HMPV 284 285 1.00 [0.98; 1.00] 299
Randolph 2016 NP NPA HMPV 106 107 0.99 [0.95; 1.00] 123
Hammitt 2011 NP OP HMPV 469 476 0.99 [0.97; 0.99] 533
Buonsenso 2023 NP S HMPV 78 78 1.00 [0.95; 1.00] 83
Haaramo 2024 NP S HMPV 57 57 1.00 [0.94; 1.00] 59
Random effects model 1859 0.99 [0.98; 0.99]
Heterogeneity: $I^2$ = 0%, $\tau^2$ = 0, $p$ = 0.9446
ref = ANS
Dawood 2015 ANS OP HMPV 661 662 1.00 [0.99; 1.00] 703
ref = NPA
DeByle 2012 NPA NP HMPV 268 271 0.99 [0.97; 1.00] 314
Tunsjo 2015 NPA NP HMPV 77 78 0.99 [0.93; 1.00] 81
Random effects model 349 0.99 [0.13; 1.00]
Heterogeneity: $I^2$ = 0%, $\tau^2$ = 0, $p$ = 0.8983
ref = MT + OP
Fourie 2023 MT + OP NP HMPV 80 80 1.00 [0.95; 1.00] 88
ref = OP
Ma 2020 OP BAL HMPV 74 75 0.99 [0.93; 1.00] 76
Wang 2019 OP IS HMPV 368 371 0.99 [0.98; 1.00] 440
Random effects model 446 0.99 [0.16; 1.00]
Heterogeneity: $I^2$ = 0%, $\tau^2$ = 0, $p$ = 0.6635
ref = MT
Onalan 2025 MT MT + OP HMPV 662 674 0.98 [0.97; 0.99] 743
ref = NP + OP
Thea 2017 NP + OP IS HMPV 958 997 0.96 [0.95; 0.97] 1114
ref = ANS + OP
Tosif 2023 ANS + OP ANS HMPV 217 218 1.00 [0.97; 1.00] 242
Random effects model 5285 0.99 [0.99; 0.99]
Heterogeneity: $I^2$ = 48.3%, $\tau^2$ = 0.4782, $p$ = 0.0072
Test for subgroup differences: $\chi^2_7$ = 41.46, df = 7 ($p$ < 0.0001)
0 0.2 0.4 0.6 0.8 1
Specificity (Index Specimen vs Reference)

g)

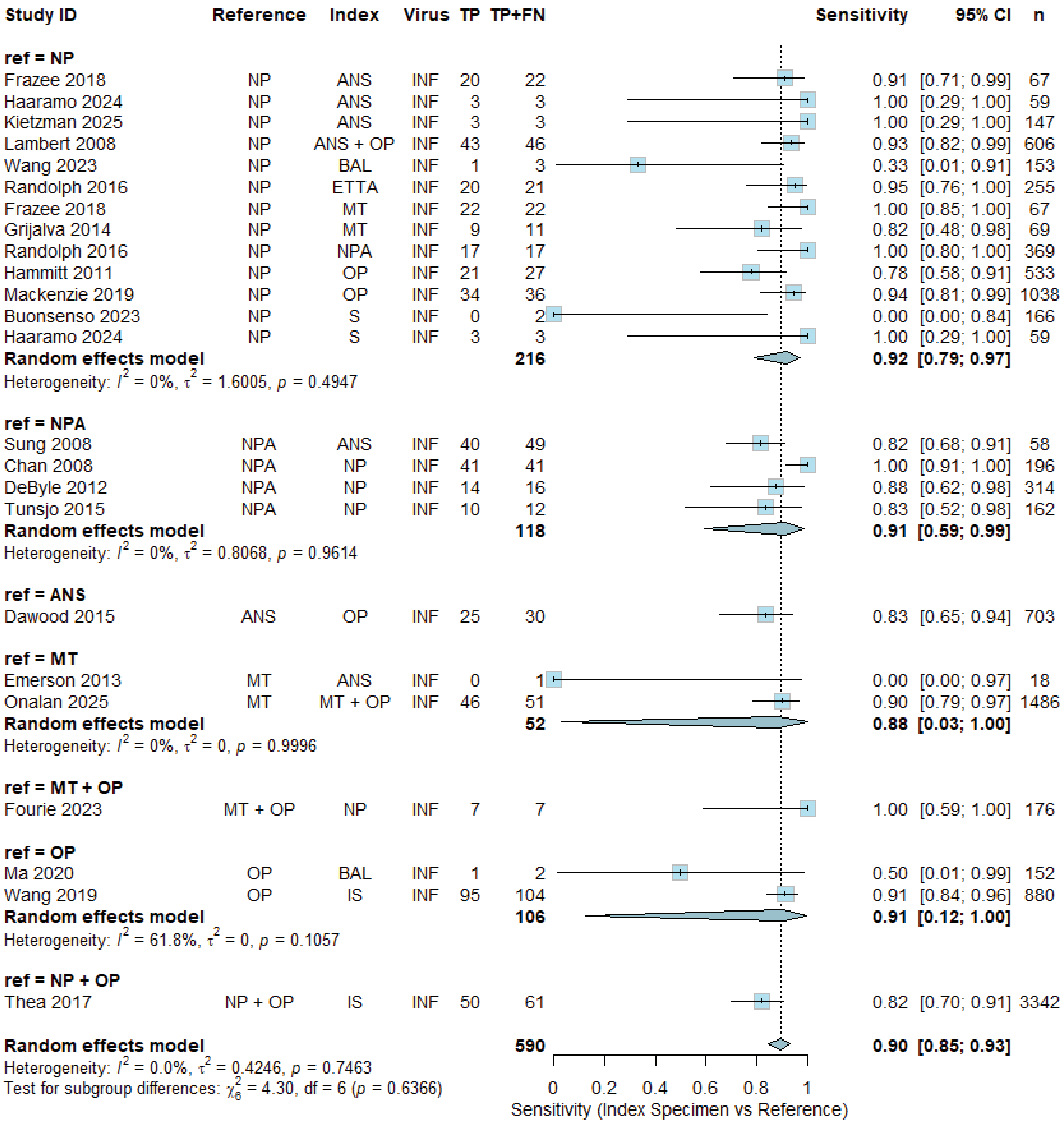

Study ID Reference Index Virus TP TP+FN Sensitivity 95% CI n
ref = NP
Frazee 2018 NP ANS INF 20 22 0.91 [0.71; 0.99] 67
Haaramo 2024 NP ANS INF 3 3 1.00 [0.29; 1.00] 59
Kietzman 2025 NP ANS INF 3 3 1.00 [0.29; 1.00] 147
Lambert 2008 NP ANS + OP INF 43 46 0.93 [0.82; 0.99] 606
Wang 2023 NP BAL INF 1 3 0.33 [0.01; 0.91] 153
Randolph 2016 NP ETTA INF 20 21 0.95 [0.76; 1.00] 255
Frazee 2018 NP MT INF 22 22 1.00 [0.85; 1.00] 67
Grijalva 2014 NP MT INF 9 11 0.82 [0.48; 0.98] 69
Randolph 2016 NP NPA INF 17 17 1.00 [0.80; 1.00] 369
Hammitt 2011 NP OP INF 21 27 0.78 [0.58; 0.91] 533
Mackenzie 2019 NP OP INF 34 36 0.94 [0.81; 0.99] 1038
Buonsenso 2023 NP S INF 0 2 0.00 [0.00; 0.84] 166
Haaramo 2024 NP S INF 3 3 1.00 [0.29; 1.00] 59
Random effects model 216 0.92 [0.79; 0.97]
Heterogeneity: $I^2$ = 0%, $\tau^2$ = 1.6005, $p$ = 0.4947
ref = NPA
Sung 2008 NPA ANS INF 40 49 0.82 [0.68; 0.91] 58
Chan 2008 NPA NP INF 41 41 1.00 [0.91; 1.00] 196
DeByle 2012 NPA NP INF 14 16 0.88 [0.62; 0.98] 314
Tunsjo 2015 NPA NP INF 10 12 0.83 [0.52; 0.98] 162
Random effects model 118 0.91 [0.59; 0.99]
Heterogeneity: $I^2$ = 0%, $\tau^2$ = 0.8068, $p$ = 0.9614
ref = ANS
Dawood 2015 ANS OP INF 25 30 0.83 [0.65; 0.94] 703
ref = MT
Emerson 2013 MT ANS INF 0 1 0.00 [0.00; 0.97] 18
Onalan 2025 MT MT + OP INF 46 51 0.90 [0.79; 0.97] 1486
Random effects model 52 0.88 [0.03; 1.00]
Heterogeneity: $I^2$ = 0%, $\tau^2$ = 0, $p$ = 0.9996
ref = MT + OP
Fourie 2023 MT + OP NP INF 7 7 1.00 [0.59; 1.00] 176
ref = OP
Ma 2020 OP BAL INF 1 2 0.50 [0.01; 0.99] 152
Wang 2019 OP IS INF 95 104 0.91 [0.84; 0.96] 880
Random effects model 106 0.91 [0.12; 1.00]
Heterogeneity: $I^2$ = 61.8%, $\tau^2$ = 0, $p$ = 0.1057
ref = NP + OP
Thea 2017 NP + OP IS INF 50 61 0.82 [0.70; 0.91] 3342
Random effects model 590 0.90 [0.85; 0.93]
Heterogeneity: $I^2$ = 0.0%, $\tau^2$ = 0.4246, $p$ = 0.7463
Test for subgroup differences: $\chi^2_6$ = 4.30, df = 6 ($p$ = 0.6366)
0 0.2 0.4 0.6 0.8 1
Sensitivity (Index Specimen vs Reference)

h)

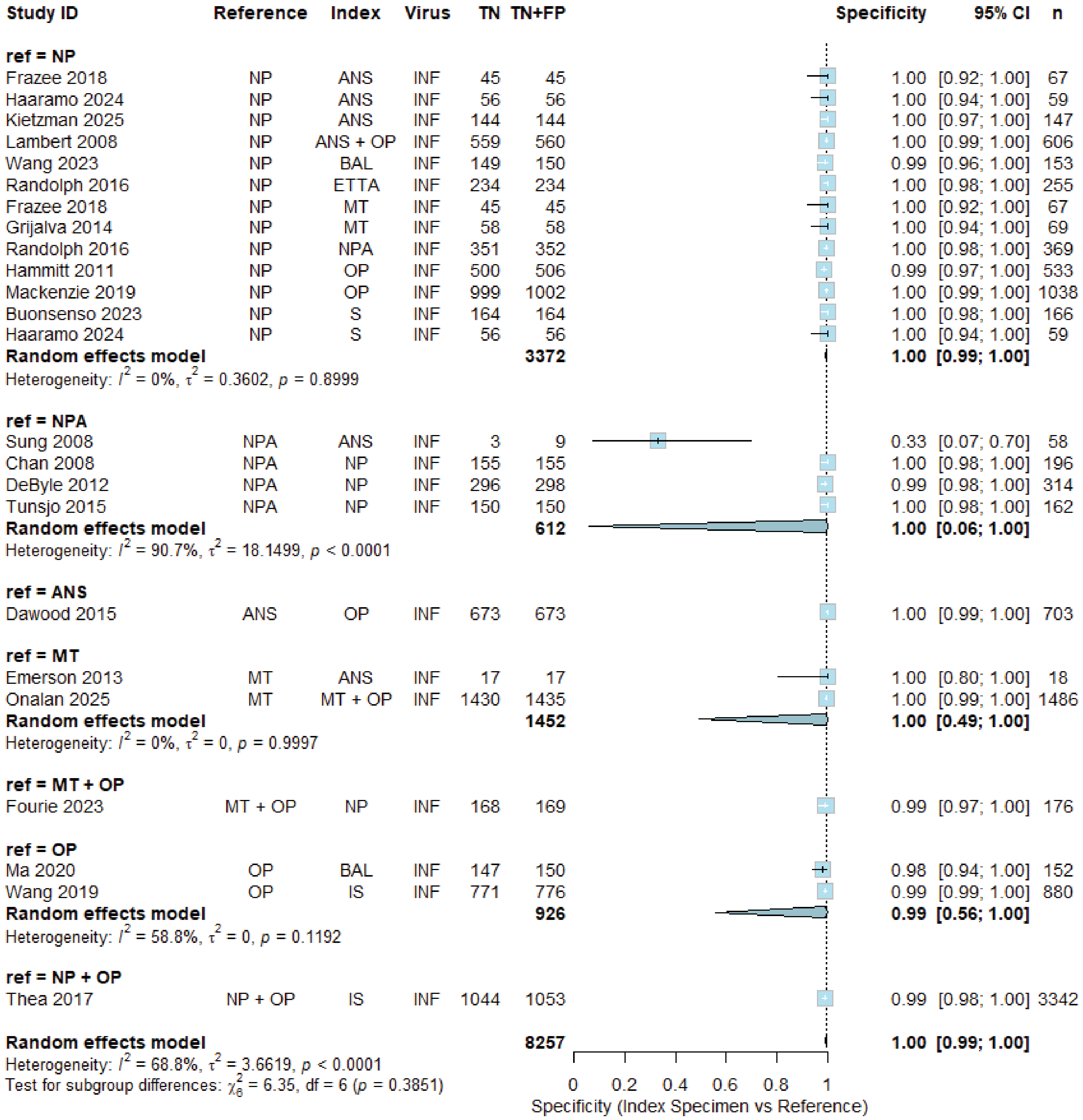

Study ID Reference Index Virus TN TN+FP Specificity 95% CI n
ref = NP
Frazee 2018 NP ANS INF 45 45 1.00 [0.92; 1.00] 67
Haaramo 2024 NP ANS INF 56 56 1.00 [0.94; 1.00] 59
Kietzman 2025 NP ANS INF 144 144 1.00 [0.97; 1.00] 147
Lambert 2008 NP ANS + OP INF 559 560 1.00 [0.99; 1.00] 606
Wang 2023 NP BAL INF 149 150 0.99 [0.96; 1.00] 153
Randolph 2016 NP ETTA INF 234 234 1.00 [0.98; 1.00] 255
Frazee 2018 NP MT INF 45 45 1.00 [0.92; 1.00] 67
Grijalva 2014 NP MT INF 58 58 1.00 [0.94; 1.00] 69
Randolph 2016 NP NPA INF 351 352 1.00 [0.98; 1.00] 369
Hammitt 2011 NP OP INF 500 506 0.99 [0.97; 1.00] 533
Mackenzie 2019 NP OP INF 999 1002 1.00 [0.99; 1.00] 1038
Buonsenso 2023 NP S INF 164 164 1.00 [0.98; 1.00] 166
Haaramo 2024 NP S INF 56 56 1.00 [0.94; 1.00] 59
Random effects model 3372 1.00 [0.99; 1.00]
Heterogeneity: $I^2$ = 0%, $\tau^2$ = 0.3602, $p$ = 0.8999
ref = NPA
Sung 2008 NPA ANS INF 3 9 0.33 [0.07; 0.70] 58
Chan 2008 NPA NP INF 155 155 1.00 [0.98; 1.00] 196
DeByle 2012 NPA NP INF 296 298 0.99 [0.98; 1.00] 314
Tunsjo 2015 NPA NP INF 150 150 1.00 [0.98; 1.00] 162
Random effects model 612 1.00 [0.06; 1.00]
Heterogeneity: $I^2$ = 90.7%, $\tau^2$ = 18.1499, $p$ < 0.0001
ref = ANS
Dawood 2015 ANS OP INF 673 673 1.00 [0.99; 1.00] 703
ref = MT
Emerson 2013 MT ANS INF 17 17 1.00 [0.80; 1.00] 18
Onalan 2025 MT MT + OP INF 1430 1435 1.00 [0.99; 1.00] 1486
Random effects model 1452 1.00 [0.49; 1.00]
Heterogeneity: $I^2$ = 0%, $\tau^2$ = 0, $p$ = 0.9997
ref = MT + OP
Fourie 2023 MT + OP NP INF 168 169 0.99 [0.97; 1.00] 176
ref = OP
Ma 2020 OP BAL INF 147 150 0.98 [0.94; 1.00] 152
Wang 2019 OP IS INF 771 776 0.99 [0.99; 1.00] 880
Random effects model 926 0.99 [0.56; 1.00]
Heterogeneity: $I^2$ = 58.8%, $\tau^2$ = 0, $p$ = 0.1192
ref = NP + OP
Thea 2017 NP + OP IS INF 1044 1053 0.99 [0.98; 1.00] 3342
Random effects model 8257 1.00 [0.99; 1.00]
Heterogeneity: $I^2$ = 68.8%, $\tau^2$ = 3.6619, $p$ < 0.0001
Test for subgroup differences: $\chi^2_6$ = 6.35, df = 6 ($p$ = 0.3851)
0 0.2 0.4 0.6 0.8 1
Specificity (Index Specimen vs Reference)

i)

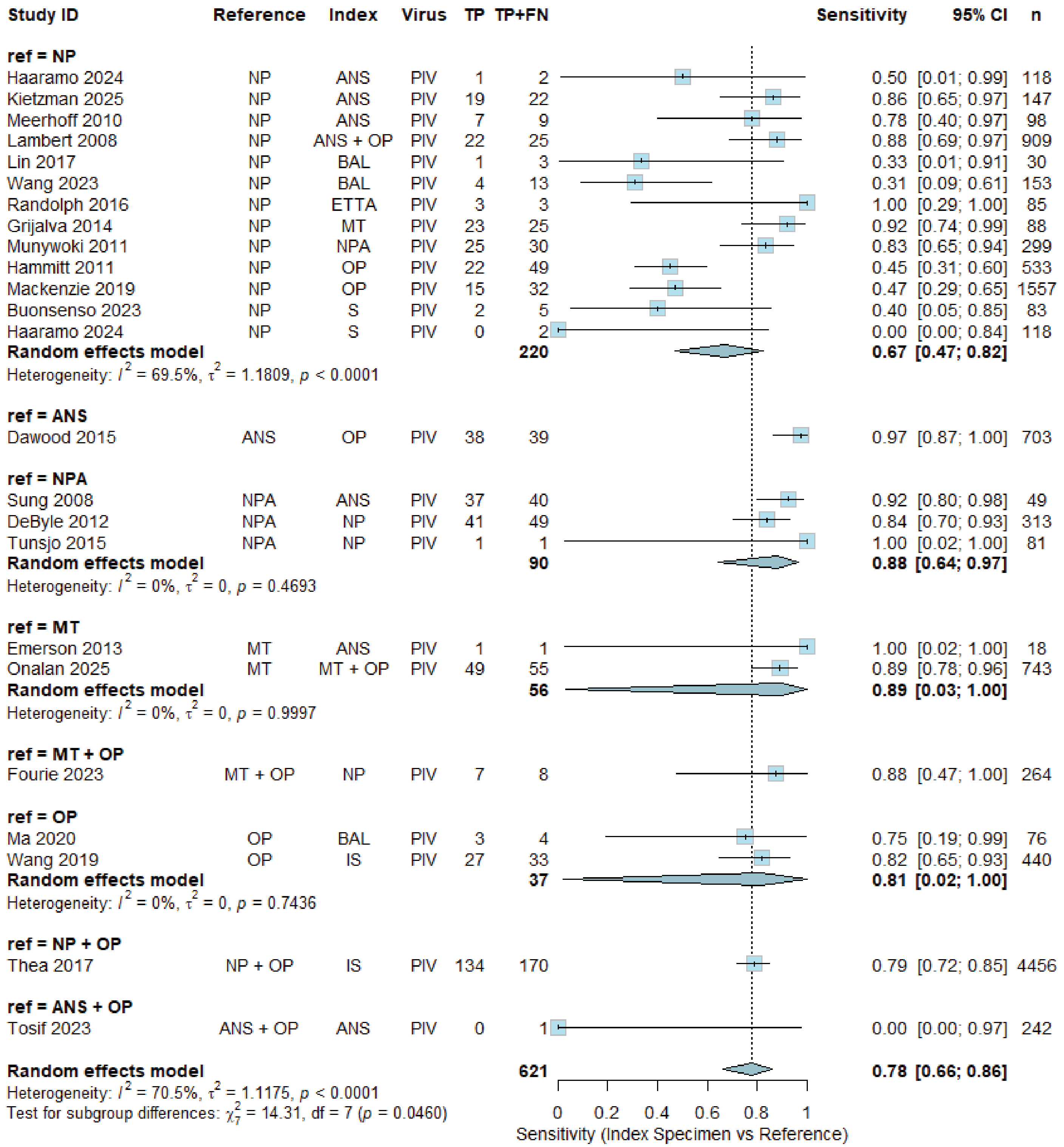

Study ID
Reference
Index
Virus
TP
TP+FN
Sensitivity
95% CI
n
ref = NP
Haaramo 2024 NP ANS PIV 1 2 0.50 [0.01; 0.99] 118
Kietzman 2025 NP ANS PIV 19 22 0.86 [0.65; 0.97] 147
Meerhoff 2010 NP ANS PIV 7 9 0.78 [0.40; 0.97] 98
Lambert 2008 NP ANS + OP PIV 22 25 0.88 [0.69; 0.97] 909
Lin 2017 NP BAL PIV 1 3 0.33 [0.01; 0.91] 30
Wang 2023 NP BAL PIV 4 13 0.31 [0.09; 0.61] 153
Randolph 2016 NP ETTA PIV 3 3 1.00 [0.29; 1.00] 85
Grijalva 2014 NP MT PIV 23 25 0.92 [0.74; 0.99] 88
Munywoki 2011 NP NPA PIV 25 30 0.83 [0.65; 0.94] 299
Hammitt 2011 NP OP PIV 22 49 0.45 [0.31; 0.60] 533
Mackenzie 2019 NP OP PIV 15 32 0.47 [0.29; 0.65] 1557
Buonsenso 2023 NP S PIV 2 5 0.40 [0.05; 0.85] 83
Haaramo 2024 NP S PIV 0 2 0.00 [0.00; 0.84] 118
Random effects model 220 0.67 [0.47; 0.82]
Heterogeneity: $I^2$ = 69.5%, $\tau^2$ = 1.1809, $p$ < 0.0001
ref = ANS
Dawood 2015 ANS OP PIV 38 39 0.97 [0.87; 1.00] 703
ref = NPA
Sung 2008 NPA ANS PIV 37 40 0.92 [0.80; 0.98] 49
DeByle 2012 NPA NP PIV 41 49 0.84 [0.70; 0.93] 313
Tunsjo 2015 NPA NP PIV 1 1 1.00 [0.02; 1.00] 81
Random effects model 90 0.88 [0.64; 0.97]
Heterogeneity: $I^2$ = 0%, $\tau^2$ = 0, $p$ = 0.4693
ref = MT
Emerson 2013 MT ANS PIV 1 1 1.00 [0.02; 1.00] 18
Onalan 2025 MT MT + OP PIV 49 55 0.89 [0.78; 0.96] 743
Random effects model 56 0.89 [0.03; 1.00]
Heterogeneity: $I^2$ = 0%, $\tau^2$ = 0, $p$ = 0.9997
ref = MT + OP
Fourie 2023 MT + OP NP PIV 7 8 0.88 [0.47; 1.00] 264
ref = OP
Ma 2020 OP BAL PIV 3 4 0.75 [0.19; 0.99] 76
Wang 2019 OP IS PIV 27 33 0.82 [0.65; 0.93] 440
Random effects model 37 0.81 [0.02; 1.00]
Heterogeneity: $I^2$ = 0%, $\tau^2$ = 0, $p$ = 0.7436
ref = NP + OP
Thea 2017 NP + OP IS PIV 134 170 0.79 [0.72; 0.85] 4456
ref = ANS + OP
Tosif 2023 ANS + OP ANS PIV 0 1 0.00 [0.00; 0.97] 242
Random effects model 621 0.78 [0.66; 0.86]
Heterogeneity: $I^2$ = 70.5%, $\tau^2$ = 1.1175, $p$ < 0.0001
Test for subgroup differences: $\chi^2_7$ = 14.31, df = 7 ($p$ = 0.0460)
0 0.2 0.4 0.6 0.8 1
Sensitivity (Index Specimen vs Reference)

j)

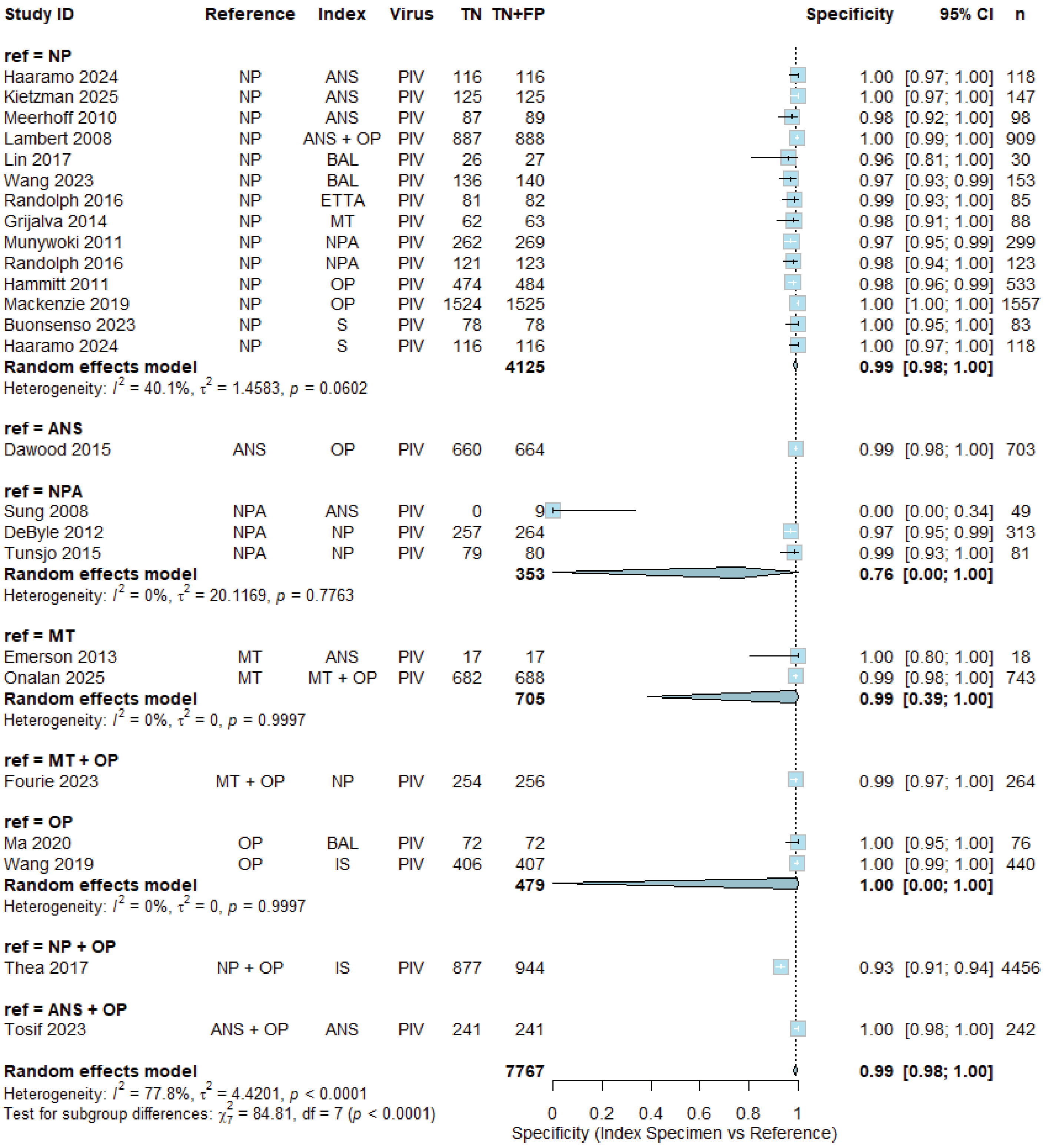
Study ID Reference Index Virus TN TN+FP Specificity 95% CI n
ref = NP
Haaramo 2024 NP ANS PIV 116 116 1.00 [0.97; 1.00] 118
Kietzman 2025 NP ANS PIV 125 125 1.00 [0.97; 1.00] 147
Meerhoff 2010 NP ANS PIV 87 89 0.98 [0.92; 1.00] 98
Lambert 2008 NP ANS + OP PIV 887 888 1.00 [0.99; 1.00] 909
Lin 2017 NP BAL PIV 26 27 0.96 [0.81; 1.00] 30
Wang 2023 NP BAL PIV 136 140 0.97 [0.93; 0.99] 153
Randolph 2016 NP ETTA PIV 81 82 0.99 [0.93; 1.00] 85
Grijalva 2014 NP MT PIV 62 63 0.98 [0.91; 1.00] 88
Munywoki 2011 NP NPA PIV 262 269 0.97 [0.95; 0.99] 299
Randolph 2016 NP NPA PIV 121 123 0.98 [0.94; 1.00] 123
Hammitt 2011 NP OP PIV 474 484 0.98 [0.96; 0.99] 533
Mackenzie 2019 NP OP PIV 1524 1525 1.00 [1.00; 1.00] 1557
Buonsenso 2023 NP S PIV 78 78 1.00 [0.95; 1.00] 83
Haaramo 2024 NP S PIV 116 116 1.00 [0.97; 1.00] 118
Random effects model 4125 0.99 [0.98; 1.00]
Heterogeneity: $I^2$ = 40.1%, $\tau^2$ = 1.4583, $p$ = 0.0602
ref = ANS
Dawood 2015 ANS OP PIV 660 664 0.99 [0.98; 1.00] 703
ref = NPA
Sung 2008 NPA ANS PIV 0 9 0.00 [0.00; 0.34] 49
DeByle 2012 NPA NP PIV 257 264 0.97 [0.95; 0.99] 313
Tunsjo 2015 NPA NP PIV 79 80 0.99 [0.93; 1.00] 81
Random effects model 353 0.76 [0.00; 1.00]
Heterogeneity: $I^2$ = 0%, $\tau^2$ = 20.1169, $p$ = 0.7763
ref = MT
Emerson 2013 MT ANS PIV 17 17 1.00 [0.80; 1.00] 18
Onalan 2025 MT MT + OP PIV 682 688 0.99 [0.98; 1.00] 743
Random effects model 705 0.99 [0.39; 1.00]
Heterogeneity: $I^2$ = 0%, $\tau^2$ = 0, $p$ = 0.9997
ref = MT + OP
Fourie 2023 MT + OP NP PIV 254 256 0.99 [0.97; 1.00] 264
ref = OP
Ma 2020 OP BAL PIV 72 72 1.00 [0.95; 1.00] 76
Wang 2019 OP IS PIV 406 407 1.00 [0.99; 1.00] 440
Random effects model 479 1.00 [0.00; 1.00]
Heterogeneity: $I^2$ = 0%, $\tau^2$ = 0, $p$ = 0.9997
ref = NP + OP
Thea 2017 NP + OP IS PIV 877 944 0.93 [0.91; 0.94] 4456
ref = ANS + OP
Tosif 2023 ANS + OP ANS PIV 241 241 1.00 [0.98; 1.00] 242
Random effects model 7767 0.99 [0.98; 1.00]
Heterogeneity: $I^2$ = 77.8%, $\tau^2$ = 4.4201, $p$ < 0.0001
Test for subgroup differences: $\chi^2_7$ = 84.81, df = 7 ($p$ < 0.0001)
0 0.2 0.4 0.6 0.8 1
Specificity (Index Specimen vs Reference)

k)

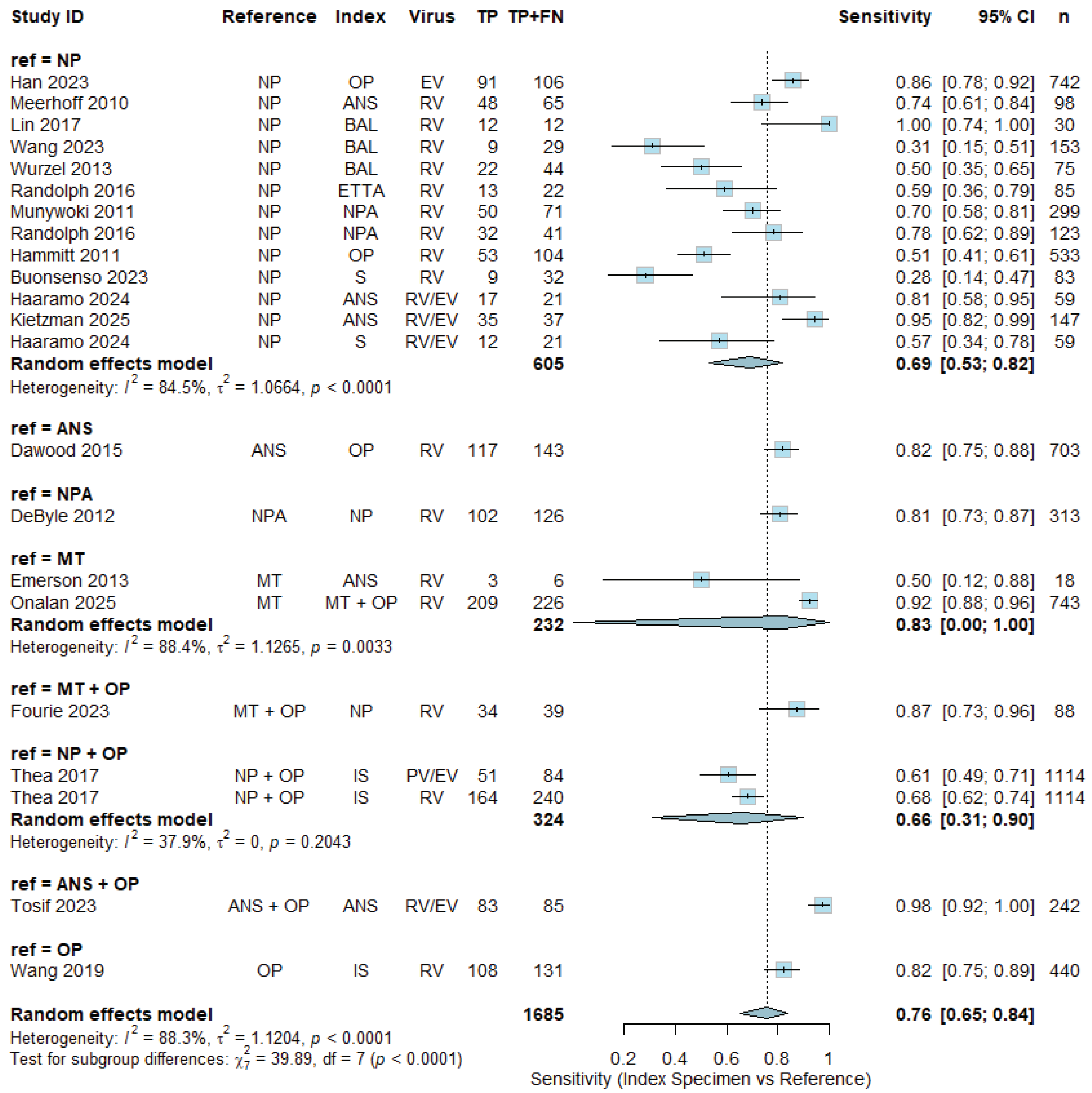

Study ID Reference Index Virus TP TP+FN Sensitivity 95% CI n
ref = NP
Han 2023 NP OP EV 91 106 0.86 [0.78; 0.92] 742
Meerhoff 2010 NP ANS RV 48 65 0.74 [0.61; 0.84] 98
Lin 2017 NP BAL RV 12 12 1.00 [0.74; 1.00] 30
Wang 2023 NP BAL RV 9 29 0.31 [0.15; 0.51] 153
Wurzel 2013 NP BAL RV 22 44 0.50 [0.35; 0.65] 75
Randolph 2016 NP ETTA RV 13 22 0.59 [0.36; 0.79] 85
Munywoki 2011 NP NPA RV 50 71 0.70 [0.58; 0.81] 299
Randolph 2016 NP NPA RV 32 41 0.78 [0.62; 0.89] 123
Hammitt 2011 NP OP RV 53 104 0.51 [0.41; 0.61] 533
Buonsenso 2023 NP S RV 9 32 0.28 [0.14; 0.47] 83
Haaramo 2024 NP ANS RV/EV 17 21 0.81 [0.58; 0.95] 59
Kietzman 2025 NP ANS RV/EV 35 37 0.95 [0.82; 0.99] 147
Haaramo 2024 NP S RV/EV 12 21 0.57 [0.34; 0.78] 59
Random effects model 605 0.69 [0.53; 0.82]
Heterogeneity: $I^2$ = 84.5%, $\tau^2$ = 1.0664, $p$ < 0.0001
ref = ANS
Dawood 2015 ANS OP RV 117 143 0.82 [0.75; 0.88] 703
ref = NPA
DeByle 2012 NPA NP RV 102 126 0.81 [0.73; 0.87] 313
ref = MT
Emerson 2013 MT ANS RV 3 6 0.50 [0.12; 0.88] 18
Onalan 2025 MT MT + OP RV 209 226 0.92 [0.88; 0.96] 743
Random effects model 232 0.83 [0.00; 1.00]
Heterogeneity: $I^2$ = 88.4%, $\tau^2$ = 1.1265, $p$ = 0.0033
ref = MT + OP
Fourie 2023 MT + OP NP RV 34 39 0.87 [0.73; 0.96] 88
ref = NP + OP
Thea 2017 NP + OP IS PV/EV 51 84 0.61 [0.49; 0.71] 1114
Thea 2017 NP + OP IS RV 164 240 0.68 [0.62; 0.74] 1114
Random effects model 324 0.66 [0.31; 0.90]
Heterogeneity: $I^2$ = 37.9%, $\tau^2$ = 0, $p$ = 0.2043
ref = ANS + OP
Tosif 2023 ANS + OP ANS RV/EV 83 85 0.98 [0.92; 1.00] 242
ref = OP
Wang 2019 OP IS RV 108 131 0.82 [0.75; 0.89] 440
Random effects model 1685 0.76 [0.65; 0.84]
Heterogeneity: $I^2$ = 88.3%, $\tau^2$ = 1.1204, $p$ < 0.0001
Test for subgroup differences: $\chi^2_7$ = 39.89, df = 7 ($p$ < 0.0001)
0.2 0.4 0.6 0.8 1
Sensitivity (Index Specimen vs Reference)

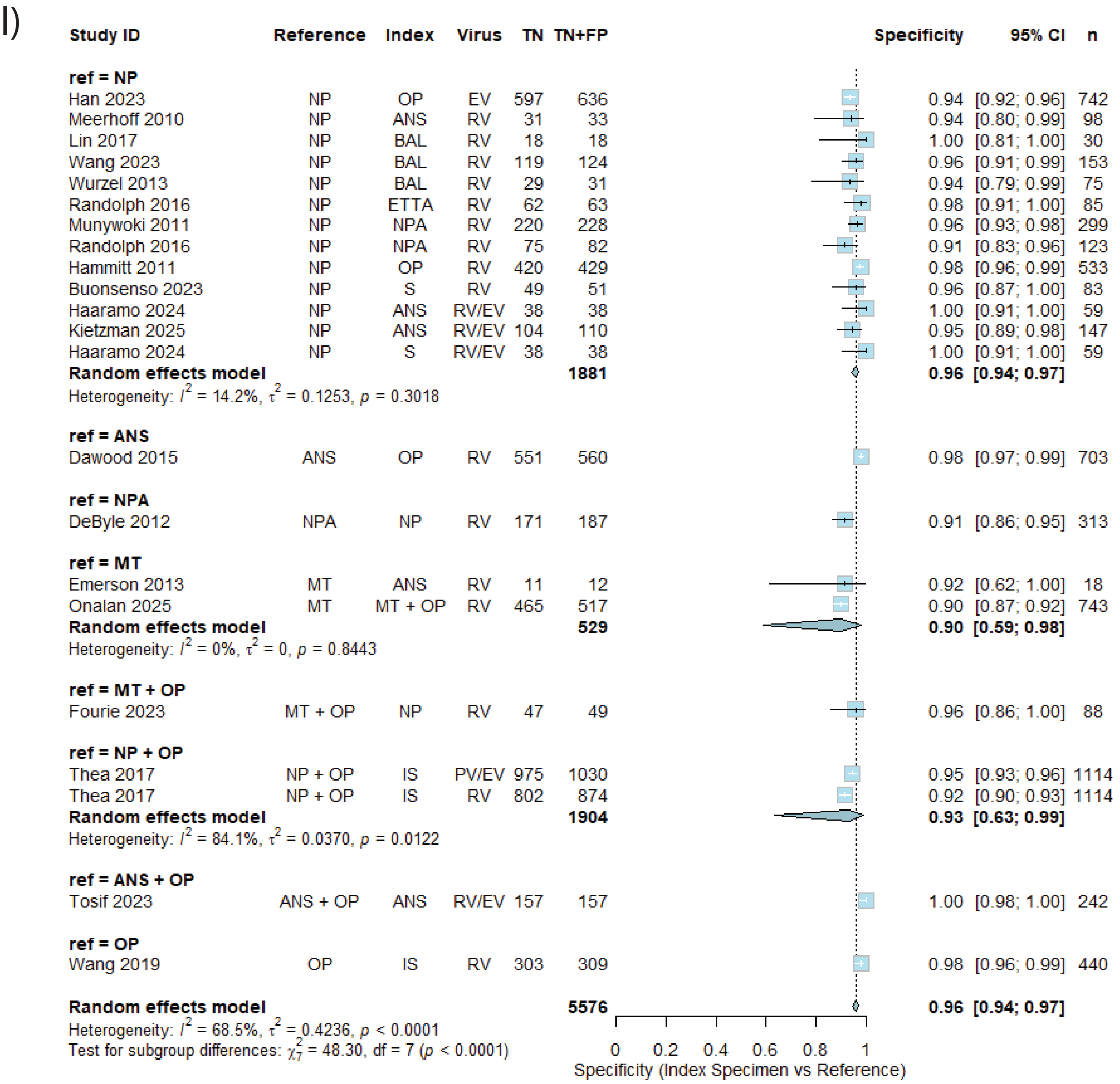
l)
Study ID
Reference
Index
Virus
TN
TN+FP
Specificity
95% CI
n
ref = NP
Han 2023 NP OP EV 597 636 0.94 [0.92; 0.96] 742
Meerhoff 2010 NP ANS RV 31 33 0.94 [0.80; 0.99] 98
Lin 2017 NP BAL RV 18 18 1.00 [0.81; 1.00] 30
Wang 2023 NP BAL RV 119 124 0.96 [0.91; 0.99] 153
Wurzel 2013 NP BAL RV 29 31 0.94 [0.79; 0.99] 75
Randolph 2016 NP ETTA RV 62 63 0.98 [0.91; 1.00] 85
Munywoki 2011 NP NPA RV 220 228 0.96 [0.93; 0.98] 299
Randolph 2016 NP NPA RV 75 82 0.91 [0.83; 0.96] 123
Hammitt 2011 NP OP RV 420 429 0.98 [0.96; 0.99] 533
Buonsenso 2023 NP S RV 49 51 0.96 [0.87; 1.00] 83
Haaramo 2024 NP ANS RV/EV 38 38 1.00 [0.91; 1.00] 59
Kietzman 2025 NP ANS RV/EV 104 110 0.95 [0.89; 0.98] 147
Haaramo 2024 NP S RV/EV 38 38 1.00 [0.91; 1.00] 59
Random effects model 1881 0.96 [0.94; 0.97]
Heterogeneity: $I^2$ = 14.2%, $\tau^2$ = 0.1253, $p$ = 0.3018
ref = ANS
Dawood 2015 ANS OP RV 551 560 0.98 [0.97; 0.99] 703
ref = NPA
DeByle 2012 NPA NP RV 171 187 0.91 [0.86; 0.95] 313
ref = MT
Emerson 2013 MT ANS RV 11 12 0.92 [0.62; 1.00] 18
Onalan 2025 MT MT + OP RV 465 517 0.90 [0.87; 0.92] 743
Random effects model 529 0.90 [0.59; 0.98]
Heterogeneity: $I^2$ = 0%, $\tau^2$ = 0, $p$ = 0.8443
ref = MT + OP
Fourie 2023 MT + OP NP RV 47 49 0.96 [0.86; 1.00] 88
ref = NP + OP
Thea 2017 NP + OP IS PV/EV 975 1030 0.95 [0.93; 0.96] 1114
Thea 2017 NP + OP IS RV 802 874 0.92 [0.90; 0.93] 1114
Random effects model 1904 0.93 [0.63; 0.99]
Heterogeneity: $I^2$ = 84.1%, $\tau^2$ = 0.0370, $p$ = 0.0122
ref = ANS + OP
Tosif 2023 ANS + OP ANS RV/EV 157 157 1.00 [0.98; 1.00] 242
ref = OP
Wang 2019 OP IS RV 303 309 0.98 [0.96; 0.99] 440
Random effects model 5576 0.96 [0.94; 0.97]
Heterogeneity: $I^2$ = 68.5%, $\tau^2$ = 0.4236, $p$ < 0.0001
Test for subgroup differences: $\chi^2_7$ = 48.30, df = 7 ($p$ < 0.0001)
0 0.2 0.4 0.6 0.8 1
Specificity (Index Specimen vs Reference)
