## Supplementary material for "One size fits all: A systematic review of the sample types used for the diagnostics of respiratory viruses in children": S4 Figure

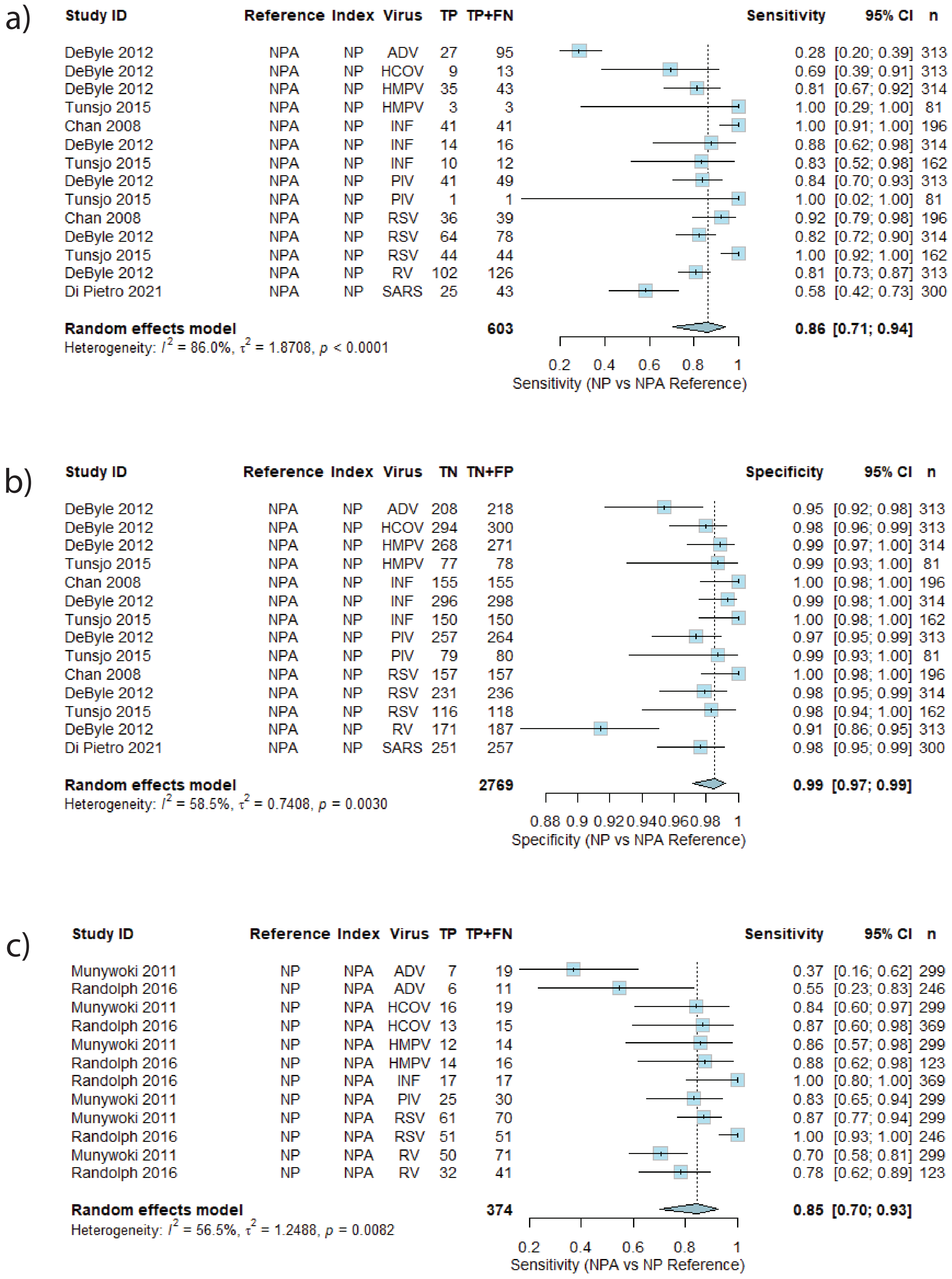
a)
Study ID Reference Index Virus TP TP+FN Sensitivity 95% CI n
DeByle 2012 NPA NP ADV 27 95 0.28 [0.20; 0.39] 313
DeByle 2012 NPA NP HCOV 9 13 0.69 [0.39; 0.91] 313
DeByle 2012 NPA NP HMPV 35 43 0.81 [0.67; 0.92] 314
Tunsjo 2015 NPA NP HMPV 3 3 1.00 [0.29; 1.00] 81
Chan 2008 NPA NP INF 41 41 1.00 [0.91; 1.00] 196
DeByle 2012 NPA NP INF 14 16 0.88 [0.62; 0.98] 314
Tunsjo 2015 NPA NP INF 10 12 0.83 [0.52; 0.98] 162
DeByle 2012 NPA NP PIV 41 49 0.84 [0.70; 0.93] 313
Tunsjo 2015 NPA NP PIV 1 1 1.00 [0.02; 1.00] 81
Chan 2008 NPA NP RSV 36 39 0.92 [0.79; 0.98] 196
DeByle 2012 NPA NP RSV 64 78 0.82 [0.72; 0.90] 314
Tunsjo 2015 NPA NP RSV 44 44 1.00 [0.92; 1.00] 162
DeByle 2012 NPA NP RV 102 126 0.81 [0.73; 0.87] 313
Di Pietro 2021 NPA NP SARS 25 43 0.58 [0.42; 0.73] 300
Random effects model 603 0.86 [0.71; 0.94]
Heterogeneity: $I^2$ = 86.0%, $\tau^2$ = 1.8708, $p$ < 0.0001
0.2 0.4 0.6 0.8 1
Sensitivity (NP vs NPA Reference)
b)
Study ID Reference Index Virus TN TN+FP Specificity 95% CI n
DeByle 2012 NPA NP ADV 208 218 0.95 [0.92; 0.98] 313
DeByle 2012 NPA NP HCOV 294 300 0.98 [0.96; 0.99] 313
DeByle 2012 NPA NP HMPV 268 271 0.99 [0.97; 1.00] 314
Tunsjo 2015 NPA NP HMPV 77 78 0.99 [0.93; 1.00] 81
Chan 2008 NPA NP INF 155 155 1.00 [0.98; 1.00] 196
DeByle 2012 NPA NP INF 296 298 0.99 [0.98; 1.00] 314
Tunsjo 2015 NPA NP INF 150 150 1.00 [0.98; 1.00] 162
DeByle 2012 NPA NP PIV 257 264 0.97 [0.95; 0.99] 313
Tunsjo 2015 NPA NP PIV 79 80 0.99 [0.93; 1.00] 81
Chan 2008 NPA NP RSV 157 157 1.00 [0.98; 1.00] 196
DeByle 2012 NPA NP RSV 231 236 0.98 [0.95; 0.99] 314
Tunsjo 2015 NPA NP RSV 116 118 0.98 [0.94; 1.00] 162
DeByle 2012 NPA NP RV 171 187 0.91 [0.86; 0.95] 313
Di Pietro 2021 NPA NP SARS 251 257 0.98 [0.95; 0.99] 300
Random effects model 2769 0.99 [0.97; 0.99]
Heterogeneity: $I^2$ = 58.5%, $\tau^2$ = 0.7408, $p$ = 0.0030
0.88 0.9 0.92 0.94 0.96 0.98 1
Specificity (NP vs NPA Reference)
c)
Study ID Reference Index Virus TP TP+FN Sensitivity 95% CI n
Munywoki 2011 NP NPA ADV 7 19 0.37 [0.16; 0.62] 299
Randolph 2016 NP NPA ADV 6 11 0.55 [0.23; 0.83] 246
Munywoki 2011 NP NPA HCOV 16 19 0.84 [0.60; 0.97] 299
Randolph 2016 NP NPA HCOV 13 15 0.87 [0.60; 0.98] 369
Munywoki 2011 NP NPA HMPV 12 14 0.86 [0.57; 0.98] 299
Randolph 2016 NP NPA HMPV 14 16 0.88 [0.62; 0.98] 123
Randolph 2016 NP NPA INF 17 17 1.00 [0.80; 1.00] 369
Munywoki 2011 NP NPA PIV 25 30 0.83 [0.65; 0.94] 299
Munywoki 2011 NP NPA RSV 61 70 0.87 [0.77; 0.94] 299
Randolph 2016 NP NPA RSV 51 51 1.00 [0.93; 1.00] 246
Munywoki 2011 NP NPA RV 50 71 0.70 [0.58; 0.81] 299
Randolph 2016 NP NPA RV 32 41 0.78 [0.62; 0.89] 123
Random effects model 374 0.85 [0.70; 0.93]
Heterogeneity: $I^2$ = 56.5%, $\tau^2$ = 1.2488, $p$ = 0.0082
0.2 0.4 0.6 0.8 1
Sensitivity (NPA vs NP Reference)

d)

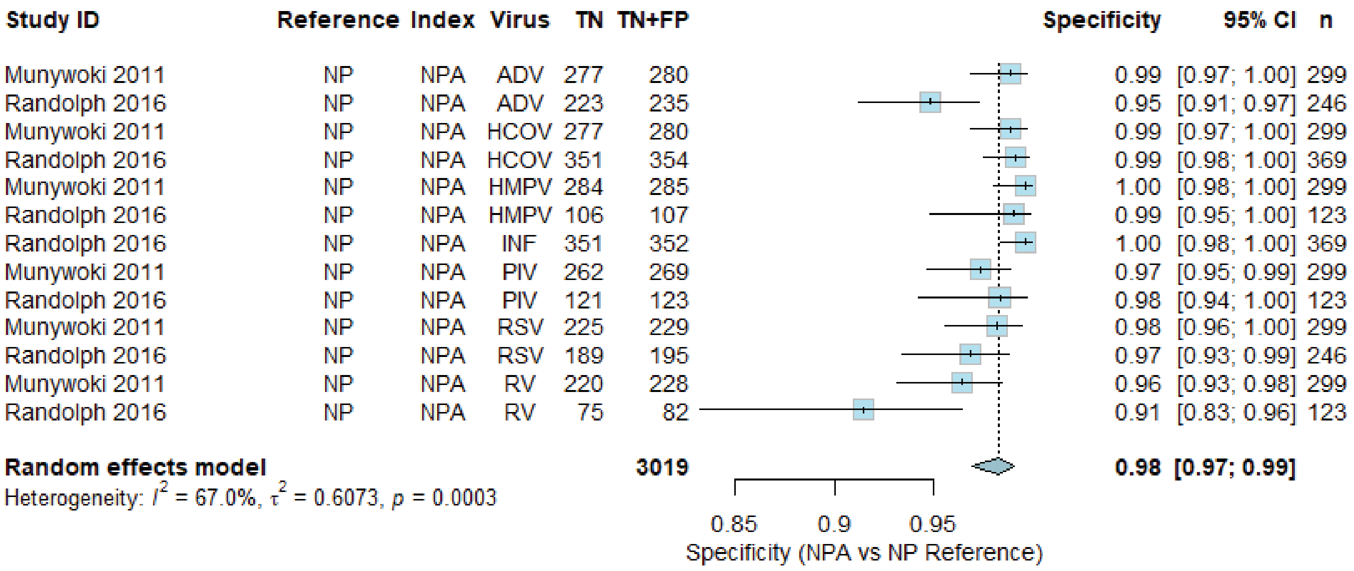
Study ID Reference Index Virus TN TN+FP Specificity 95% CI n
Munywoki 2011 NP NPA ADV 277 280 0.99 [0.97; 1.00] 299
Randolph 2016 NP NPA ADV 223 235 0.95 [0.91; 0.97] 246
Munywoki 2011 NP NPA HCOV 277 280 0.99 [0.97; 1.00] 299
Randolph 2016 NP NPA HCOV 351 354 0.99 [0.98; 1.00] 369
Munywoki 2011 NP NPA HMPV 284 285 1.00 [0.98; 1.00] 299
Randolph 2016 NP NPA HMPV 106 107 0.99 [0.95; 1.00] 123
Randolph 2016 NP NPA INF 351 352 1.00 [0.98; 1.00] 369
Munywoki 2011 NP NPA PIV 262 269 0.97 [0.95; 0.99] 299
Randolph 2016 NP NPA PIV 121 123 0.98 [0.94; 1.00] 123
Munywoki 2011 NP NPA RSV 225 229 0.98 [0.96; 1.00] 299
Randolph 2016 NP NPA RSV 189 195 0.97 [0.93; 0.99] 246
Munywoki 2011 NP NPA RV 220 228 0.96 [0.93; 0.98] 299
Randolph 2016 NP NPA RV 75 82 0.91 [0.83; 0.96] 123
Random effects model 3019 0.98 [0.97; 0.99]
Heterogeneity: I^2 = 67.0%, τ^2 = 0.6073, p = 0.0003
0.85 0.9 0.95
Specificity (NPA vs NP Reference)

e)

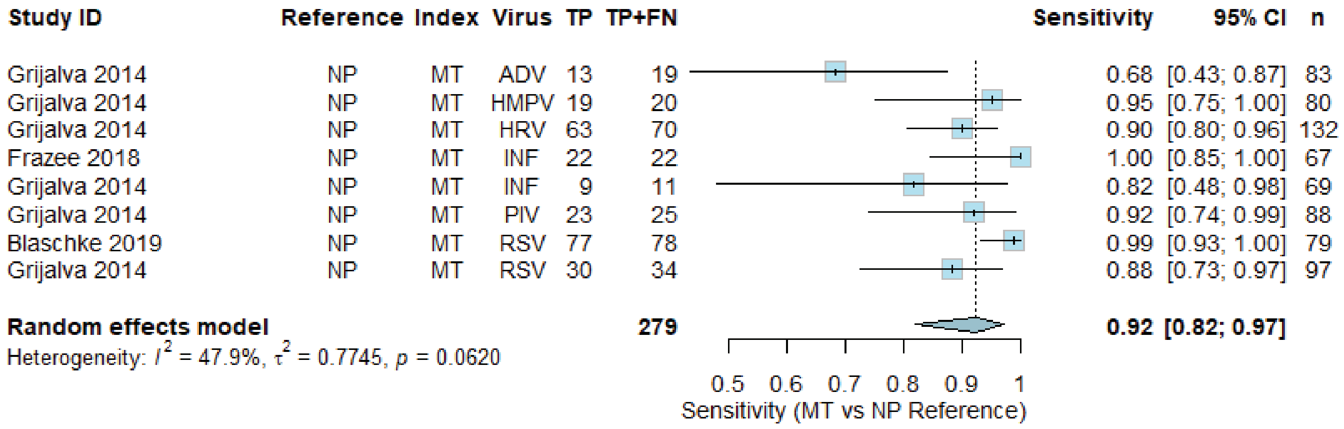
Study ID Reference Index Virus TP TP+FN Sensitivity 95% CI n
Grijalva 2014 NP MT ADV 13 19 0.68 [0.43; 0.87] 83
Grijalva 2014 NP MT HMPV 19 20 0.95 [0.75; 1.00] 80
Grijalva 2014 NP MT HRV 63 70 0.90 [0.80; 0.96] 132
Frazee 2018 NP MT INF 22 22 1.00 [0.85; 1.00] 67
Grijalva 2014 NP MT INF 9 11 0.82 [0.48; 0.98] 69
Grijalva 2014 NP MT PIV 23 25 0.92 [0.74; 0.99] 88
Blaschke 2019 NP MT RSV 77 78 0.99 [0.93; 1.00] 79
Grijalva 2014 NP MT RSV 30 34 0.88 [0.73; 0.97] 97
Random effects model 279 0.92 [0.82; 0.97]
Heterogeneity: I^2 = 47.9%, τ^2 = 0.7745, p = 0.0620
0.5 0.6 0.7 0.8 0.9 1
Sensitivity (MT vs NP Reference)

f)

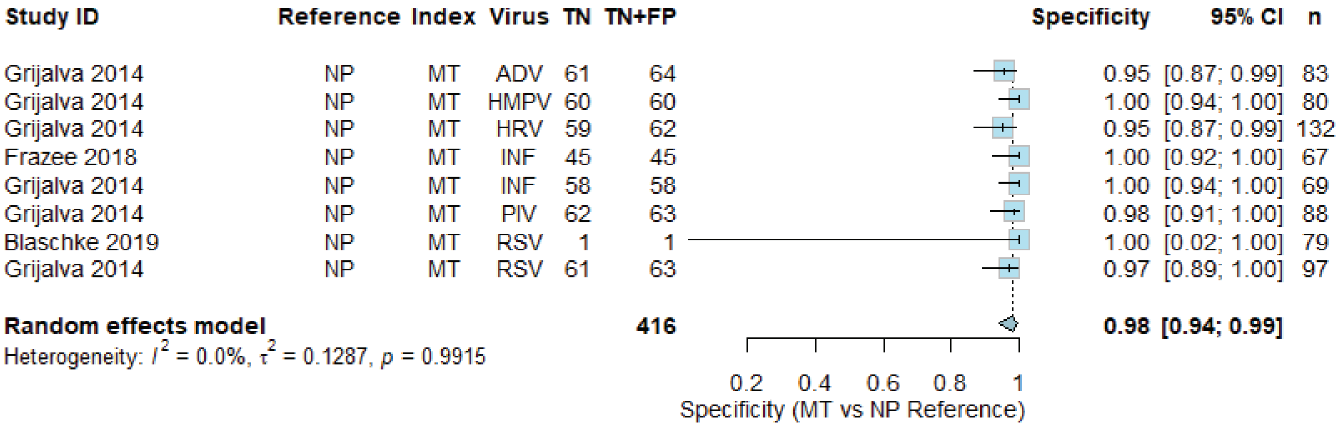
Study ID Reference Index Virus TN TN+FP Specificity 95% CI n
Grijalva 2014 NP MT ADV 61 64 0.95 [0.87; 0.99] 83
Grijalva 2014 NP MT HMPV 60 60 1.00 [0.94; 1.00] 80
Grijalva 2014 NP MT HRV 59 62 0.95 [0.87; 0.99] 132
Frazee 2018 NP MT INF 45 45 1.00 [0.92; 1.00] 67
Grijalva 2014 NP MT INF 58 58 1.00 [0.94; 1.00] 69
Grijalva 2014 NP MT PIV 62 63 0.98 [0.91; 1.00] 88
Blaschke 2019 NP MT RSV 1 1 1.00 [0.02; 1.00] 79
Grijalva 2014 NP MT RSV 61 63 0.97 [0.89; 1.00] 97
Random effects model 416 0.98 [0.94; 0.99]
Heterogeneity: I^2 = 0.0%, τ^2 = 0.1287, p = 0.9915
0.2 0.4 0.6 0.8 1
Specificity (MT vs NP Reference)

g)

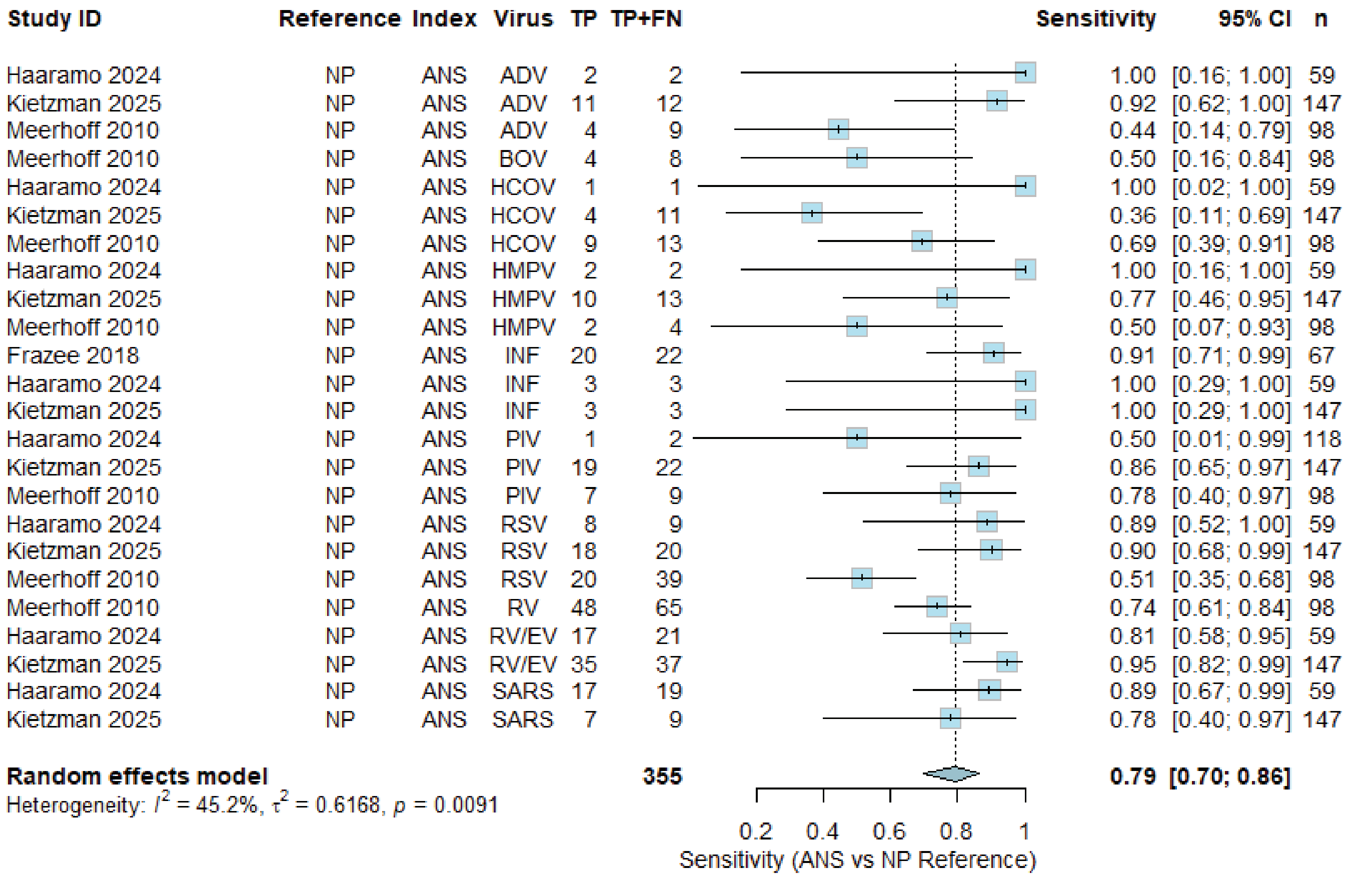
Study ID Reference Index Virus TP TP+FN Sensitivity 95% CI n
Haaramo 2024 NP ANS ADV 2 2 1.00 [0.16; 1.00] 59
Kietzman 2025 NP ANS ADV 11 12 0.92 [0.62; 1.00] 147
Meerhoff 2010 NP ANS ADV 4 9 0.44 [0.14; 0.79] 98
Meerhoff 2010 NP ANS BOV 4 8 0.50 [0.16; 0.84] 98
Haaramo 2024 NP ANS HCOV 1 1 1.00 [0.02; 1.00] 59
Kietzman 2025 NP ANS HCOV 4 11 0.36 [0.11; 0.69] 147
Meerhoff 2010 NP ANS HCOV 9 13 0.69 [0.39; 0.91] 98
Haaramo 2024 NP ANS HMPV 2 2 1.00 [0.16; 1.00] 59
Kietzman 2025 NP ANS HMPV 10 13 0.77 [0.46; 0.95] 147
Meerhoff 2010 NP ANS HMPV 2 4 0.50 [0.07; 0.93] 98
Frazee 2018 NP ANS INF 20 22 0.91 [0.71; 0.99] 67
Haaramo 2024 NP ANS INF 3 3 1.00 [0.29; 1.00] 59
Kietzman 2025 NP ANS INF 3 3 1.00 [0.29; 1.00] 147
Haaramo 2024 NP ANS PIV 1 2 0.50 [0.01; 0.99] 118
Kietzman 2025 NP ANS PIV 19 22 0.86 [0.65; 0.97] 147
Meerhoff 2010 NP ANS PIV 7 9 0.78 [0.40; 0.97] 98
Haaramo 2024 NP ANS RSV 8 9 0.89 [0.52; 1.00] 59
Kietzman 2025 NP ANS RSV 18 20 0.90 [0.68; 0.99] 147
Meerhoff 2010 NP ANS RSV 20 39 0.51 [0.35; 0.68] 98
Meerhoff 2010 NP ANS RV 48 65 0.74 [0.61; 0.84] 98
Haaramo 2024 NP ANS RV/EV 17 21 0.81 [0.58; 0.95] 59
Kietzman 2025 NP ANS RV/EV 35 37 0.95 [0.82; 0.99] 147
Haaramo 2024 NP ANS SARS 17 19 0.89 [0.67; 0.99] 59
Kietzman 2025 NP ANS SARS 7 9 0.78 [0.40; 0.97] 147
Random effects model 355 0.79 [0.70; 0.86]
Heterogeneity: $I^2$ = 45.2%, $\tau^2$ = 0.6168, $p$ = 0.0091
0.2 0.4 0.6 0.8 1
Sensitivity (ANS vs NP Reference)

h)

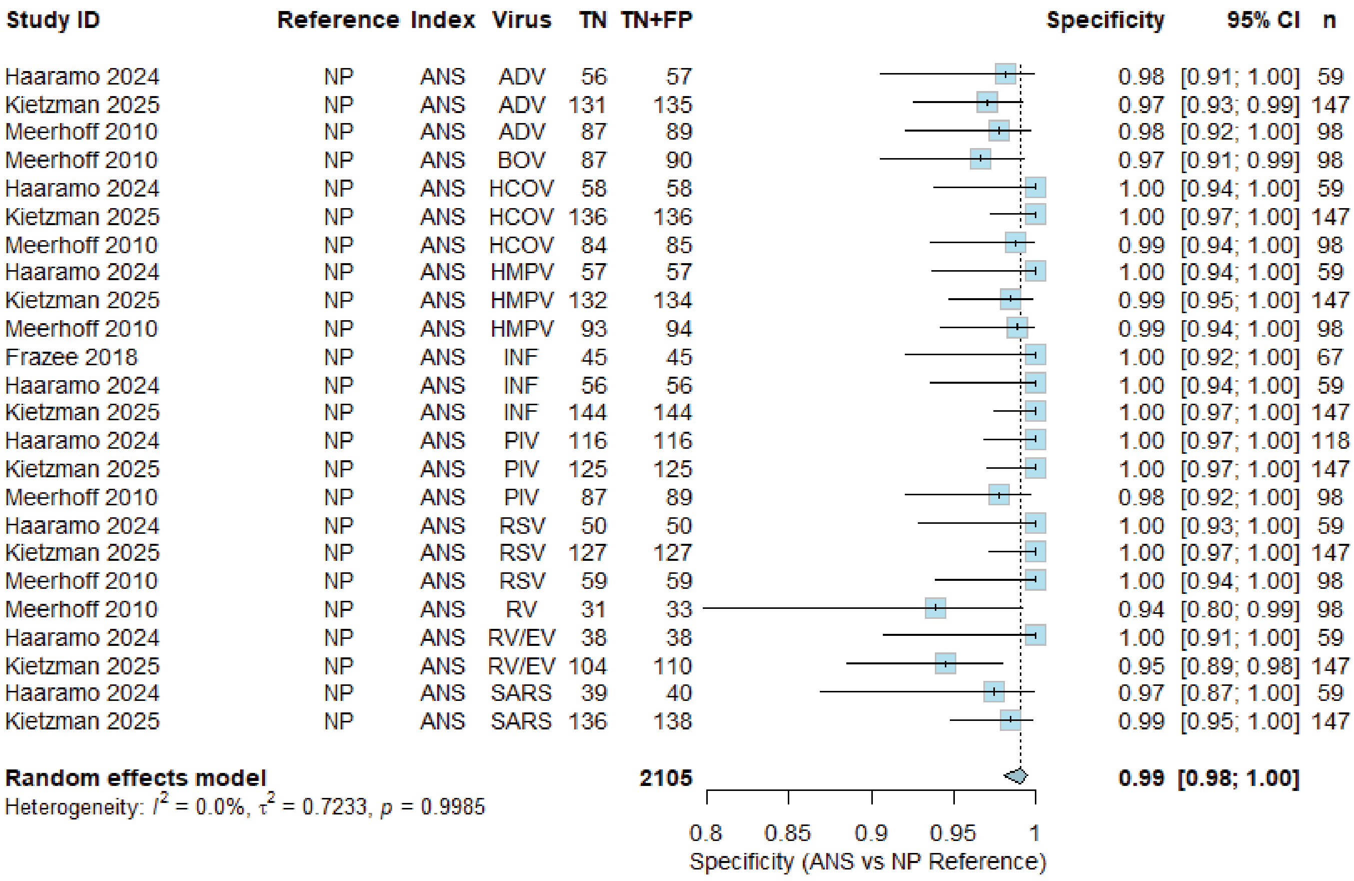
Study ID Reference Index Virus TN TN+FP Specificity 95% CI n
Haaramo 2024 NP ANS ADV 56 57 0.98 [0.91; 1.00] 59
Kietzman 2025 NP ANS ADV 131 135 0.97 [0.93; 0.99] 147
Meerhoff 2010 NP ANS ADV 87 89 0.98 [0.92; 1.00] 98
Meerhoff 2010 NP ANS BOV 87 90 0.97 [0.91; 0.99] 98
Haaramo 2024 NP ANS HCOV 58 58 1.00 [0.94; 1.00] 59
Kietzman 2025 NP ANS HCOV 136 136 1.00 [0.97; 1.00] 147
Meerhoff 2010 NP ANS HCOV 84 85 0.99 [0.94; 1.00] 98
Haaramo 2024 NP ANS HMPV 57 57 1.00 [0.94; 1.00] 59
Kietzman 2025 NP ANS HMPV 132 134 0.99 [0.95; 1.00] 147
Meerhoff 2010 NP ANS HMPV 93 94 0.99 [0.94; 1.00] 98
Frazee 2018 NP ANS INF 45 45 1.00 [0.92; 1.00] 67
Haaramo 2024 NP ANS INF 56 56 1.00 [0.94; 1.00] 59
Kietzman 2025 NP ANS INF 144 144 1.00 [0.97; 1.00] 147
Haaramo 2024 NP ANS PIV 116 116 1.00 [0.97; 1.00] 118
Kietzman 2025 NP ANS PIV 125 125 1.00 [0.97; 1.00] 147
Meerhoff 2010 NP ANS PIV 87 89 0.98 [0.92; 1.00] 98
Haaramo 2024 NP ANS RSV 50 50 1.00 [0.93; 1.00] 59
Kietzman 2025 NP ANS RSV 127 127 1.00 [0.97; 1.00] 147
Meerhoff 2010 NP ANS RSV 59 59 1.00 [0.94; 1.00] 98
Meerhoff 2010 NP ANS RV 31 33 0.94 [0.80; 0.99] 98
Haaramo 2024 NP ANS RV/EV 38 38 1.00 [0.91; 1.00] 59
Kietzman 2025 NP ANS RV/EV 104 110 0.95 [0.89; 0.98] 147
Haaramo 2024 NP ANS SARS 39 40 0.97 [0.87; 1.00] 59
Kietzman 2025 NP ANS SARS 136 138 0.99 [0.95; 1.00] 147
Random effects model 2105 0.99 [0.98; 1.00]
Heterogeneity: $I^2$ = 0.0%, $\tau^2$ = 0.7233, $p$ = 0.9985
0.8 0.85 0.9 0.95 1
Specificity (ANS vs NP Reference)

i)

| Study ID | Reference | Index | Virus | TP | TP+FN | Sensitivity | 95% CI | n |
|---|---|---|---|---|---|---|---|---|
| Hammitt 2011 | NP | OP | ADV | 29 | 51 | 0.57 | [0.42; 0.71] | 533 |
| Han 2023 | NP | OP | EV | 91 | 106 | 0.86 | [0.78; 0.92] | 742 |
| Hammitt 2011 | NP | OP | HCOV | 12 | 22 | 0.55 | [0.32; 0.76] | 533 |
| Hammitt 2011 | NP | OP | HMPV | 34 | 57 | 0.60 | [0.46; 0.72] | 533 |
| Hammitt 2011 | NP | OP | INF | 21 | 27 | 0.78 | [0.58; 0.91] | 533 |
| Mackenzie 2019 | NP | OP | INF | 34 | 36 | 0.94 | [0.81; 0.99] | 1038 |
| Hammitt 2011 | NP | OP | PIV | 22 | 49 | 0.45 | [0.31; 0.60] | 533 |
| Mackenzie 2019 | NP | OP | PIV | 15 | 32 | 0.47 | [0.29; 0.65] | 1557 |
| Hammitt 2011 | NP | OP | RSV | 83 | 107 | 0.78 | [0.68; 0.85] | 533 |
| Mackenzie 2019 | NP | OP | RSV | 211 | 237 | 0.89 | [0.84; 0.93] | 1038 |
| Hammitt 2011 | NP | OP | RV | 53 | 104 | 0.51 | [0.41; 0.61] | 533 |
| **Random effects model** | | | | | **828** | **0.70** | **[0.56; 0.82]** | |

Heterogeneity: $I^2 = 90.6\%$, $\tau^2 = 0.7575$, $p < 0.0001$

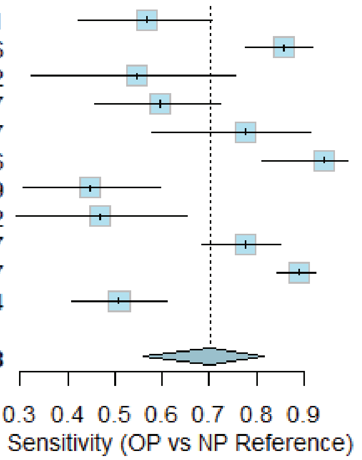


j)

| Study ID | Reference | Index | Virus | TN | TN+FP | Specificity | 95% CI | n |
|---|---|---|---|---|---|---|---|---|
| Hammitt 2011 | NP | OP | ADV | 466 | 482 | 0.97 | [0.95; 0.98] | 533 |
| Han 2023 | NP | OP | EV | 597 | 636 | 0.94 | [0.92; 0.96] | 742 |
| Hammitt 2011 | NP | OP | HCOV | 508 | 511 | 0.99 | [0.98; 1.00] | 533 |
| Hammitt 2011 | NP | OP | HMPV | 469 | 476 | 0.99 | [0.97; 0.99] | 533 |
| Hammitt 2011 | NP | OP | INF | 500 | 506 | 0.99 | [0.97; 1.00] | 533 |
| Mackenzie 2019 | NP | OP | INF | 999 | 1002 | 1.00 | [0.99; 1.00] | 1038 |
| Hammitt 2011 | NP | OP | PIV | 474 | 484 | 0.98 | [0.96; 0.99] | 533 |
| Mackenzie 2019 | NP | OP | PIV | 1524 | 1525 | 1.00 | [1.00; 1.00] | 1557 |
| Hammitt 2011 | NP | OP | RSV | 413 | 426 | 0.97 | [0.95; 0.98] | 533 |
| Mackenzie 2019 | NP | OP | RSV | 794 | 801 | 0.99 | [0.98; 1.00] | 1038 |
| Hammitt 2011 | NP | OP | RV | 420 | 429 | 0.98 | [0.96; 0.99] | 533 |
| **Random effects model** | | | | | **7278** | **0.99** | **[0.97; 0.99]** | |

Heterogeneity: $I^2 = 87.6\%$, $\tau^2 = 1.1190$, $p < 0.0001$

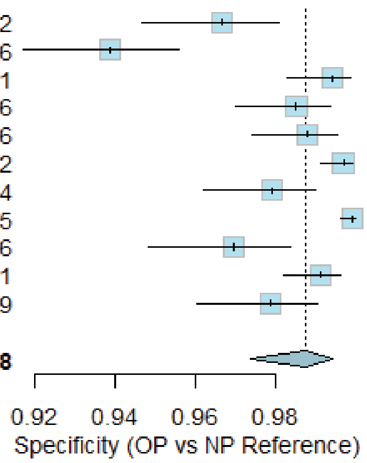


k)

| Study ID | Reference | Index | Virus | TP | TP+FN | Sensitivity | 95% CI | n |
|---|---|---|---|---|---|---|---|---|
| Lin 2017 | NP | BAL | ADV | 2 | 5 | 0.40 | [0.05; 0.85] | 30 |
| Wang 2023 | NP | BAL | ADV | 37 | 38 | 0.97 | [0.86; 1.00] | 153 |
| Wurzel 2013 | NP | BAL | ADV | 9 | 15 | 0.60 | [0.32; 0.84] | 75 |
| Wang 2023 | NP | BAL | BOV | 0 | 1 | 0.00 | [0.00; 0.97] | 153 |
| Wurzel 2013 | NP | BAL | BOV | 2 | 7 | 0.29 | [0.04; 0.71] | 75 |
| Lin 2017 | NP | BAL | HCOV | 1 | 3 | 0.33 | [0.01; 0.91] | 30 |
| Wang 2023 | NP | BAL | HCOV | 0 | 7 | 0.00 | [0.00; 0.41] | 153 |
| Wurzel 2013 | NP | BAL | HCOV | 1 | 4 | 0.25 | [0.01; 0.81] | 75 |
| Wang 2023 | NP | BAL | HMPV | 1 | 7 | 0.14 | [0.00; 0.58] | 153 |
| Wang 2023 | NP | BAL | INF | 1 | 3 | 0.33 | [0.01; 0.91] | 153 |
| Lin 2017 | NP | BAL | PIV | 1 | 3 | 0.33 | [0.01; 0.91] | 30 |
| Wang 2023 | NP | BAL | PIV | 4 | 13 | 0.31 | [0.09; 0.61] | 153 |
| Wurzel 2013 | NP | BAL | PYV | 0 | 4 | 0.00 | [0.00; 0.60] | 75 |
| Lin 2017 | NP | BAL | RSV | 4 | 4 | 1.00 | [0.40; 1.00] | 30 |
| Wang 2023 | NP | BAL | RSV | 0 | 7 | 0.00 | [0.00; 0.41] | 153 |
| Lin 2017 | NP | BAL | RV | 12 | 12 | 1.00 | [0.74; 1.00] | 30 |
| Wang 2023 | NP | BAL | RV | 9 | 29 | 0.31 | [0.15; 0.51] | 153 |
| Wurzel 2013 | NP | BAL | RV | 22 | 44 | 0.50 | [0.35; 0.65] | 75 |
| **Random effects model** | | | | | **206** | **0.37** | **[0.16; 0.65]** | |

Heterogeneity: $I^2 = 26.5\%$, $\tau^2 = 3.8383$, $p = 0.1454$

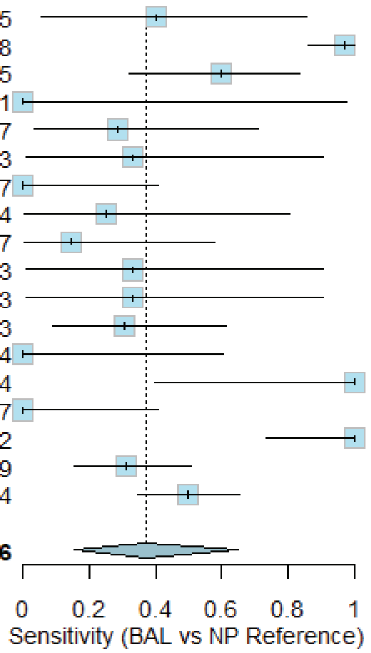

l)

| Study ID | Reference | Index | Virus | TN | TN+FP | Specificity | 95% CI | n |
|---|---|---|---|---|---|---|---|---|
| Lin 2017 | NP | BAL | ADV | 24 | 25 | 0.96 | [0.80; 1.00] | 30 |
| Wang 2023 | NP | BAL | ADV | 63 | 115 | 0.55 | [0.45; 0.64] | 153 |
| Wurzel 2013 | NP | BAL | ADV | 56 | 60 | 0.93 | [0.84; 0.98] | 75 |
| Wang 2023 | NP | BAL | BOV | 151 | 152 | 0.99 | [0.96; 1.00] | 153 |
| Wurzel 2013 | NP | BAL | BOV | 66 | 68 | 0.97 | [0.90; 1.00] | 75 |
| Lin 2017 | NP | BAL | HCOV | 26 | 27 | 0.96 | [0.81; 1.00] | 30 |
| Wang 2023 | NP | BAL | HCOV | 145 | 146 | 0.99 | [0.96; 1.00] | 153 |
| Wurzel 2013 | NP | BAL | HCOV | 69 | 71 | 0.97 | [0.90; 1.00] | 75 |
| Wang 2023 | NP | BAL | HMPV | 144 | 146 | 0.99 | [0.95; 1.00] | 153 |
| Wang 2023 | NP | BAL | INF | 149 | 150 | 0.99 | [0.96; 1.00] | 153 |
| Lin 2017 | NP | BAL | PIV | 26 | 27 | 0.96 | [0.81; 1.00] | 30 |
| Wang 2023 | NP | BAL | PIV | 136 | 140 | 0.97 | [0.93; 0.99] | 153 |
| Wurzel 2013 | NP | BAL | PYV | 70 | 71 | 0.99 | [0.92; 1.00] | 75 |
| Lin 2017 | NP | BAL | RSV | 26 | 26 | 1.00 | [0.87; 1.00] | 30 |
| Wang 2023 | NP | BAL | RSV | 145 | 146 | 0.99 | [0.96; 1.00] | 153 |
| Lin 2017 | NP | BAL | RV | 18 | 18 | 1.00 | [0.81; 1.00] | 30 |
| Wang 2023 | NP | BAL | RV | 119 | 124 | 0.96 | [0.91; 0.99] | 153 |
| Wurzel 2013 | NP | BAL | RV | 29 | 31 | 0.94 | [0.79; 0.99] | 75 |
| **Random effects model** | | | | | **1543** | **0.98** | **[0.95; 0.99]** | |

Heterogeneity: $I^2 = 90.2\%$, $\tau^2 = 1.5436$, $p < 0.0001$

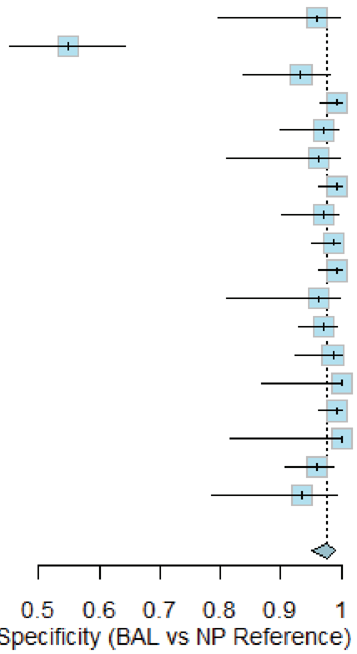
