## Supplementary material for "One size fits all: A systematic review of the sample types used for the diagnostics of respiratory viruses in children": S5 Figure

a)

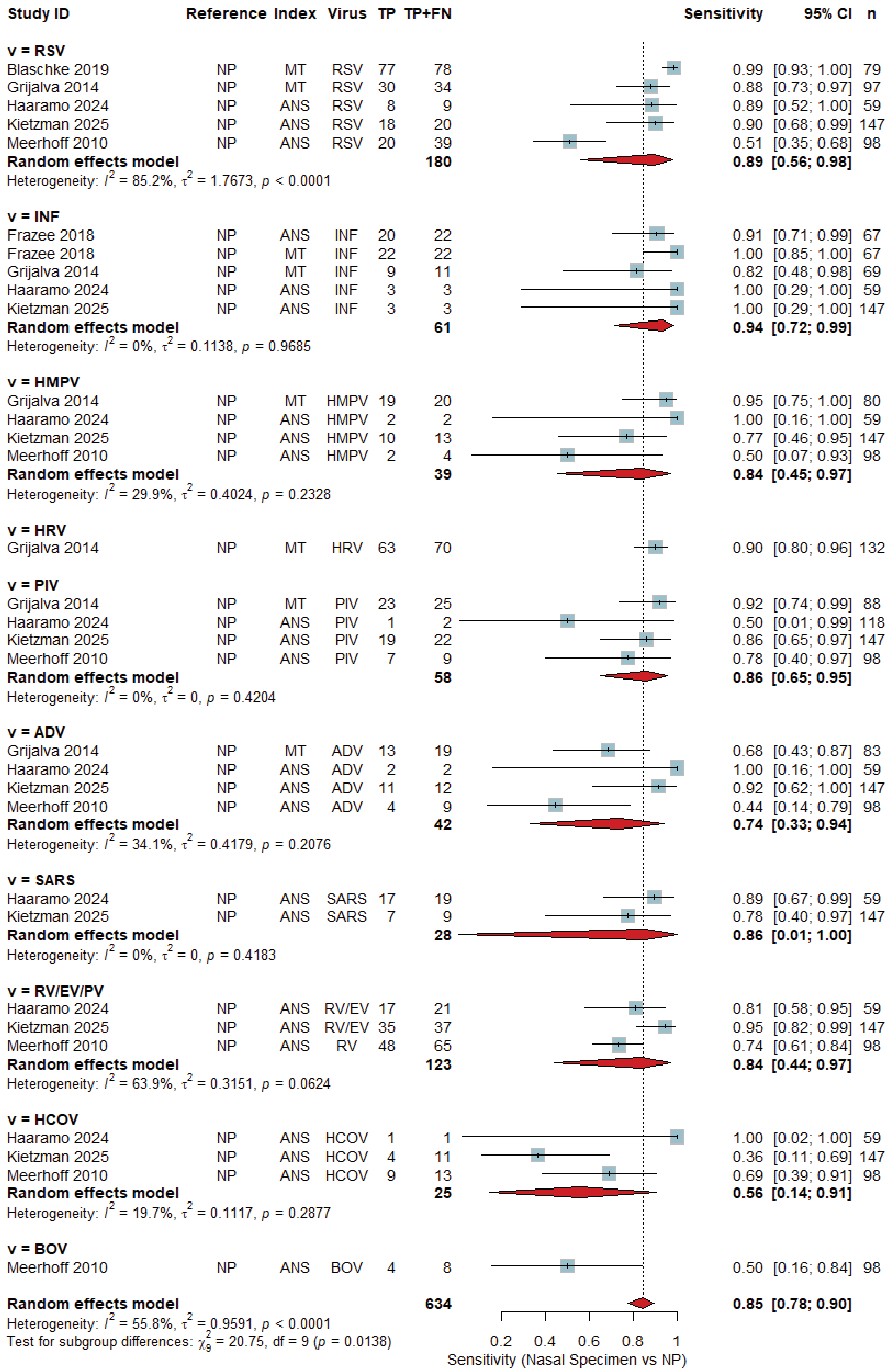
Study ID
Reference
Index
Virus
TP
TP+FN
Sensitivity
95% CI
n
v = RSV
Blaschke 2019 NP MT RSV 77 78 0.99 [0.93; 1.00] 79
Grijalva 2014 NP MT RSV 30 34 0.88 [0.73; 0.97] 97
Haaramo 2024 NP ANS RSV 8 9 0.89 [0.52; 1.00] 59
Kietzman 2025 NP ANS RSV 18 20 0.90 [0.68; 0.99] 147
Meerhoff 2010 NP ANS RSV 20 39 0.51 [0.35; 0.68] 98
Random effects model 180 0.89 [0.56; 0.98]
Heterogeneity: $I^2$ = 85.2%, $\tau^2$ = 1.7673, $p$ < 0.0001
v = INF
Frazee 2018 NP ANS INF 20 22 0.91 [0.71; 0.99] 67
Frazee 2018 NP MT INF 22 22 1.00 [0.85; 1.00] 67
Grijalva 2014 NP MT INF 9 11 0.82 [0.48; 0.98] 69
Haaramo 2024 NP ANS INF 3 3 1.00 [0.29; 1.00] 59
Kietzman 2025 NP ANS INF 3 3 1.00 [0.29; 1.00] 147
Random effects model 61 0.94 [0.72; 0.99]
Heterogeneity: $I^2$ = 0%, $\tau^2$ = 0.1138, $p$ = 0.9685
v = HMPV
Grijalva 2014 NP MT HMPV 19 20 0.95 [0.75; 1.00] 80
Haaramo 2024 NP ANS HMPV 2 2 1.00 [0.16; 1.00] 59
Kietzman 2025 NP ANS HMPV 10 13 0.77 [0.46; 0.95] 147
Meerhoff 2010 NP ANS HMPV 2 4 0.50 [0.07; 0.93] 98
Random effects model 39 0.84 [0.45; 0.97]
Heterogeneity: $I^2$ = 29.9%, $\tau^2$ = 0.4024, $p$ = 0.2328
v = HRV
Grijalva 2014 NP MT HRV 63 70 0.90 [0.80; 0.96] 132
v = PIV
Grijalva 2014 NP MT PIV 23 25 0.92 [0.74; 0.99] 88
Haaramo 2024 NP ANS PIV 1 2 0.50 [0.01; 0.99] 118
Kietzman 2025 NP ANS PIV 19 22 0.86 [0.65; 0.97] 147
Meerhoff 2010 NP ANS PIV 7 9 0.78 [0.40; 0.97] 98
Random effects model 58 0.86 [0.65; 0.95]
Heterogeneity: $I^2$ = 0%, $\tau^2$ = 0, $p$ = 0.4204
v = ADV
Grijalva 2014 NP MT ADV 13 19 0.68 [0.43; 0.87] 83
Haaramo 2024 NP ANS ADV 2 2 1.00 [0.16; 1.00] 59
Kietzman 2025 NP ANS ADV 11 12 0.92 [0.62; 1.00] 147
Meerhoff 2010 NP ANS ADV 4 9 0.44 [0.14; 0.79] 98
Random effects model 42 0.74 [0.33; 0.94]
Heterogeneity: $I^2$ = 34.1%, $\tau^2$ = 0.4179, $p$ = 0.2076
v = SARS
Haaramo 2024 NP ANS SARS 17 19 0.89 [0.67; 0.99] 59
Kietzman 2025 NP ANS SARS 7 9 0.78 [0.40; 0.97] 147
Random effects model 28 0.86 [0.01; 1.00]
Heterogeneity: $I^2$ = 0%, $\tau^2$ = 0, $p$ = 0.4183
v = RV/EV/PV
Haaramo 2024 NP ANS RV/EV 17 21 0.81 [0.58; 0.95] 59
Kietzman 2025 NP ANS RV/EV 35 37 0.95 [0.82; 0.99] 147
Meerhoff 2010 NP ANS RV 48 65 0.74 [0.61; 0.84] 98
Random effects model 123 0.84 [0.44; 0.97]
Heterogeneity: $I^2$ = 63.9%, $\tau^2$ = 0.3151, $p$ = 0.0624
v = HCOV
Haaramo 2024 NP ANS HCOV 1 1 1.00 [0.02; 1.00] 59
Kietzman 2025 NP ANS HCOV 4 11 0.36 [0.11; 0.69] 147
Meerhoff 2010 NP ANS HCOV 9 13 0.69 [0.39; 0.91] 98
Random effects model 25 0.56 [0.14; 0.91]
Heterogeneity: $I^2$ = 19.7%, $\tau^2$ = 0.1117, $p$ = 0.2877
v = BOV
Meerhoff 2010 NP ANS BOV 4 8 0.50 [0.16; 0.84] 98
Random effects model 634 0.85 [0.78; 0.90]
Heterogeneity: $I^2$ = 55.8%, $\tau^2$ = 0.9591, $p$ < 0.0001
Test for subgroup differences: $\chi^2_9$ = 20.75, df = 9 ($p$ = 0.0138)
0.2 0.4 0.6 0.8 1
Sensitivity (Nasal Specimen vs NP)

b)

| Study ID | Reference | Index | Virus | TN | TN+FP | Specificity | 95% CI | n |
|---|---|---|---|---|---|---|---|---|
| **v = RSV** | | | | | | | | |
| Blaschke 2019 | NP | MT | RSV | 1 | 1 | 1.00 | [0.02; 1.00] | 79 |
| Grijalva 2014 | NP | MT | RSV | 61 | 63 | 0.97 | [0.89; 1.00] | 97 |
| Haaramo 2024 | NP | ANS | RSV | 50 | 50 | 1.00 | [0.93; 1.00] | 59 |
| Kietzman 2025 | NP | ANS | RSV | 127 | 127 | 1.00 | [0.97; 1.00] | 147 |
| Meerhoff 2010 | NP | ANS | RSV | 59 | 59 | 1.00 | [0.94; 1.00] | 98 |
| **Random effects model** | | | | | **300** | **1.00** | **[0.51; 1.00]** | |
| Heterogeneity: $I^2 = 0\%$, $\tau^2 = 2.5130$, $p = 1.0000$ | | | | | | | | |
| **v = INF** | | | | | | | | |
| Frazee 2018 | NP | ANS | INF | 45 | 45 | 1.00 | [0.92; 1.00] | 67 |
| Frazee 2018 | NP | MT | INF | 45 | 45 | 1.00 | [0.92; 1.00] | 67 |
| Grijalva 2014 | NP | MT | INF | 58 | 58 | 1.00 | [0.94; 1.00] | 69 |
| Haaramo 2024 | NP | ANS | INF | 56 | 56 | 1.00 | [0.94; 1.00] | 59 |
| Kietzman 2025 | NP | ANS | INF | 144 | 144 | 1.00 | [0.97; 1.00] | 147 |
| **Random effects model** | | | | | **348** | **1.00** | **[0.00; 1.00]** | |
| Heterogeneity: $I^2 = 0\%$, $\tau^2 = 0$, $p = 1.0000$ | | | | | | | | |
| **v = HMPV** | | | | | | | | |
| Grijalva 2014 | NP | MT | HMPV | 60 | 60 | 1.00 | [0.94; 1.00] | 80 |
| Haaramo 2024 | NP | ANS | HMPV | 57 | 57 | 1.00 | [0.94; 1.00] | 59 |
| Kietzman 2025 | NP | ANS | HMPV | 132 | 134 | 0.99 | [0.95; 1.00] | 147 |
| Meerhoff 2010 | NP | ANS | HMPV | 93 | 94 | 0.99 | [0.94; 1.00] | 98 |
| **Random effects model** | | | | | **345** | **0.99** | **[0.95; 1.00]** | |
| Heterogeneity: $I^2 = 0\%$, $\tau^2 = 0$, $p = 0.9944$ | | | | | | | | |
| **v = HRV** | | | | | | | | |
| Grijalva 2014 | NP | MT | HRV | 59 | 62 | 0.95 | [0.87; 0.99] | 132 |
| **v = PIV** | | | | | | | | |
| Grijalva 2014 | NP | MT | PIV | 62 | 63 | 0.98 | [0.91; 1.00] | 88 |
| Haaramo 2024 | NP | ANS | PIV | 116 | 116 | 1.00 | [0.97; 1.00] | 118 |
| Kietzman 2025 | NP | ANS | PIV | 125 | 125 | 1.00 | [0.97; 1.00] | 147 |
| Meerhoff 2010 | NP | ANS | PIV | 87 | 89 | 0.98 | [0.92; 1.00] | 98 |
| **Random effects model** | | | | | **393** | **0.99** | **[0.92; 1.00]** | |
| Heterogeneity: $I^2 = 0\%$, $\tau^2 = 0.3478$, $p = 0.9939$ | | | | | | | | |
| **v = ADV** | | | | | | | | |
| Grijalva 2014 | NP | MT | ADV | 61 | 64 | 0.95 | [0.87; 0.99] | 83 |
| Haaramo 2024 | NP | ANS | ADV | 56 | 57 | 0.98 | [0.91; 1.00] | 59 |
| Kietzman 2025 | NP | ANS | ADV | 131 | 135 | 0.97 | [0.93; 0.99] | 147 |
| Meerhoff 2010 | NP | ANS | ADV | 87 | 89 | 0.98 | [0.92; 1.00] | 98 |
| **Random effects model** | | | | | **345** | **0.97** | **[0.92; 0.99]** | |
| Heterogeneity: $I^2 = 0\%$, $\tau^2 = 0$, $p = 0.7815$ | | | | | | | | |
| **v = SARS** | | | | | | | | |
| Haaramo 2024 | NP | ANS | SARS | 39 | 40 | 0.97 | [0.87; 1.00] | 59 |
| Kietzman 2025 | NP | ANS | SARS | 136 | 138 | 0.99 | [0.95; 1.00] | 147 |
| **Random effects model** | | | | | **178** | **0.98** | **[0.03; 1.00]** | |
| Heterogeneity: $I^2 = 0\%$, $\tau^2 = 0$, $p = 0.6534$ | | | | | | | | |
| **v = RV/EV/PV** | | | | | | | | |
| Haaramo 2024 | NP | ANS | RV/EV | 38 | 38 | 1.00 | [0.91; 1.00] | 59 |
| Kietzman 2025 | NP | ANS | RV/EV | 104 | 110 | 0.95 | [0.89; 0.98] | 147 |
| Meerhoff 2010 | NP | ANS | RV | 31 | 33 | 0.94 | [0.80; 0.99] | 98 |
| **Random effects model** | | | | | **181** | **0.96** | **[0.82; 0.99]** | |
| Heterogeneity: $I^2 = 0\%$, $\tau^2 = 0$, $p = 0.9912$ | | | | | | | | |
| **v = HCOV** | | | | | | | | |
| Haaramo 2024 | NP | ANS | HCOV | 58 | 58 | 1.00 | [0.94; 1.00] | 59 |
| Kietzman 2025 | NP | ANS | HCOV | 136 | 136 | 1.00 | [0.97; 1.00] | 147 |
| Meerhoff 2010 | NP | ANS | HCOV | 84 | 85 | 0.99 | [0.94; 1.00] | 98 |
| **Random effects model** | | | | | **279** | **1.00** | **[0.79; 1.00]** | |
| Heterogeneity: $I^2 = 0\%$, $\tau^2 = 0$, $p = 1.0000$ | | | | | | | | |
| **v = BOV** | | | | | | | | |
| Meerhoff 2010 | NP | ANS | BOV | 87 | 90 | 0.97 | [0.91; 0.99] | 98 |
| **Random effects model** | | | | | **2521** | **0.99** | **[0.98; 0.99]** | |
| Heterogeneity: $I^2 = 0.0\%$, $\tau^2 = 0.6499$, $p = 0.9999$ | | | | | | | | |
| Test for subgroup differences: $\chi^2_9 = 16.11$, df = 9 ($p = 0.0645$) | | | | | | | | |

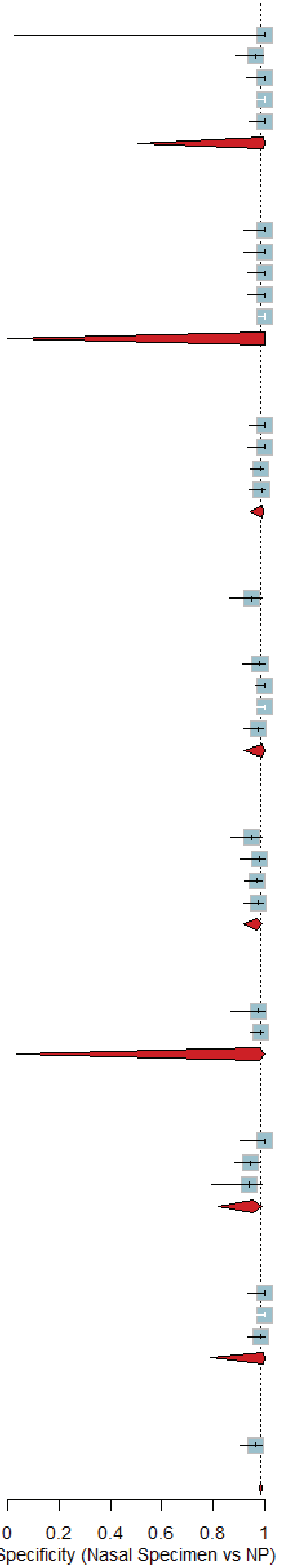

c)

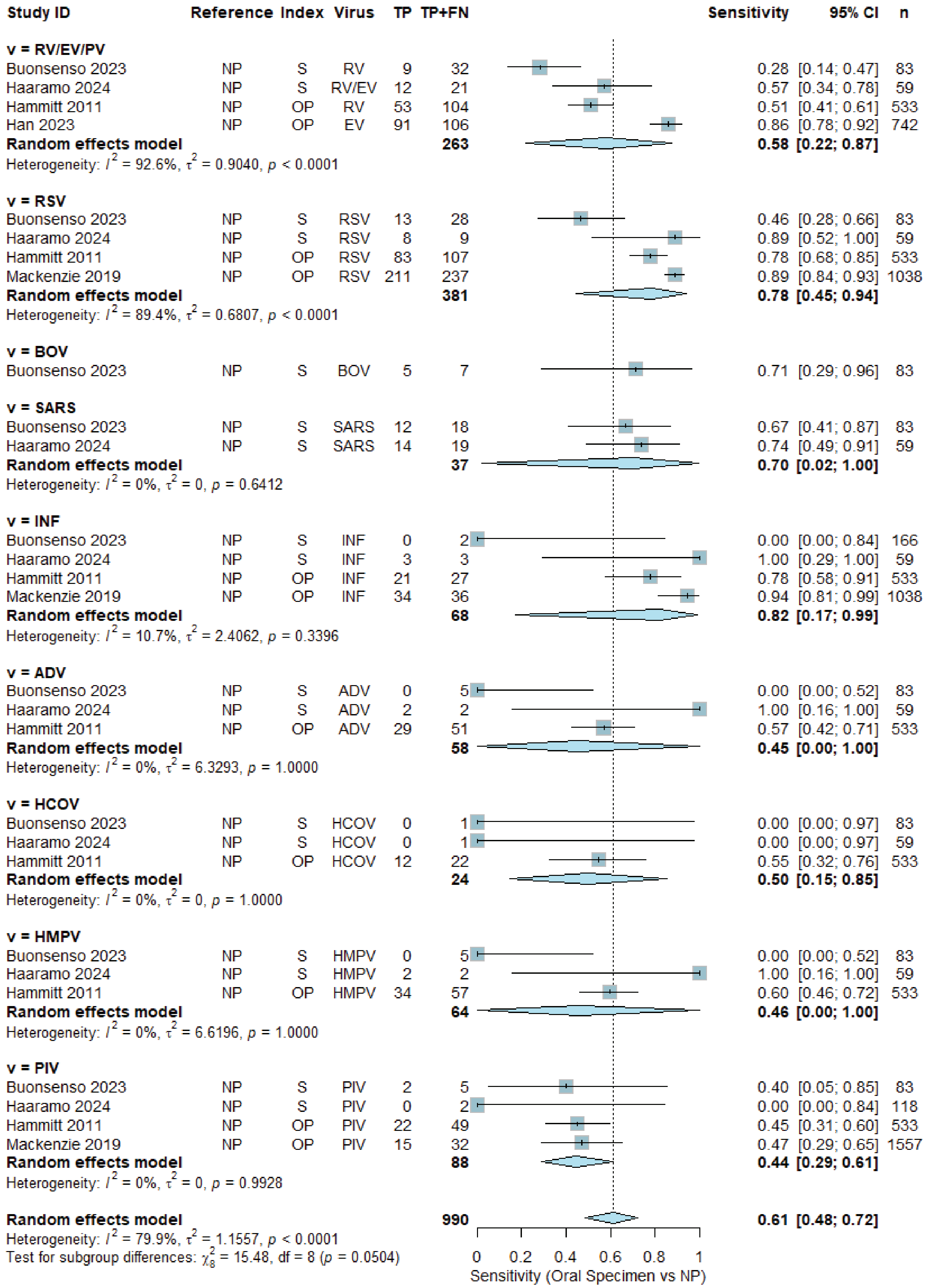

Study ID Reference Index Virus TP TP+FN Sensitivity 95% CI n
v = RV/EV/PV
Buonsenso 2023 NP S RV 9 32 0.28 [0.14; 0.47] 83
Haaramo 2024 NP S RV/EV 12 21 0.57 [0.34; 0.78] 59
Hammitt 2011 NP OP RV 53 104 0.51 [0.41; 0.61] 533
Han 2023 NP OP EV 91 106 0.86 [0.78; 0.92] 742
Random effects model 263 0.58 [0.22; 0.87]
Heterogeneity: $I^2$ = 92.6%, $\tau^2$ = 0.9040, $p$ < 0.0001
v = RSV
Buonsenso 2023 NP S RSV 13 28 0.46 [0.28; 0.66] 83
Haaramo 2024 NP S RSV 8 9 0.89 [0.52; 1.00] 59
Hammitt 2011 NP OP RSV 83 107 0.78 [0.68; 0.85] 533
Mackenzie 2019 NP OP RSV 211 237 0.89 [0.84; 0.93] 1038
Random effects model 381 0.78 [0.45; 0.94]
Heterogeneity: $I^2$ = 89.4%, $\tau^2$ = 0.6807, $p$ < 0.0001
v = BOV
Buonsenso 2023 NP S BOV 5 7 0.71 [0.29; 0.96] 83
v = SARS
Buonsenso 2023 NP S SARS 12 18 0.67 [0.41; 0.87] 83
Haaramo 2024 NP S SARS 14 19 0.74 [0.49; 0.91] 59
Random effects model 37 0.70 [0.02; 1.00]
Heterogeneity: $I^2$ = 0%, $\tau^2$ = 0, $p$ = 0.6412
v = INF
Buonsenso 2023 NP S INF 0 2 0.00 [0.00; 0.84] 166
Haaramo 2024 NP S INF 3 3 1.00 [0.29; 1.00] 59
Hammitt 2011 NP OP INF 21 27 0.78 [0.58; 0.91] 533
Mackenzie 2019 NP OP INF 34 36 0.94 [0.81; 0.99] 1038
Random effects model 68 0.82 [0.17; 0.99]
Heterogeneity: $I^2$ = 10.7%, $\tau^2$ = 2.4062, $p$ = 0.3396
v = ADV
Buonsenso 2023 NP S ADV 0 5 0.00 [0.00; 0.52] 83
Haaramo 2024 NP S ADV 2 2 1.00 [0.16; 1.00] 59
Hammitt 2011 NP OP ADV 29 51 0.57 [0.42; 0.71] 533
Random effects model 58 0.45 [0.00; 1.00]
Heterogeneity: $I^2$ = 0%, $\tau^2$ = 6.3293, $p$ = 1.0000
v = HCOV
Buonsenso 2023 NP S HCOV 0 1 0.00 [0.00; 0.97] 83
Haaramo 2024 NP S HCOV 0 1 0.00 [0.00; 0.97] 59
Hammitt 2011 NP OP HCOV 12 22 0.55 [0.32; 0.76] 533
Random effects model 24 0.50 [0.15; 0.85]
Heterogeneity: $I^2$ = 0%, $\tau^2$ = 0, $p$ = 1.0000
v = HMPV
Buonsenso 2023 NP S HMPV 0 5 0.00 [0.00; 0.52] 83
Haaramo 2024 NP S HMPV 2 2 1.00 [0.16; 1.00] 59
Hammitt 2011 NP OP HMPV 34 57 0.60 [0.46; 0.72] 533
Random effects model 64 0.46 [0.00; 1.00]
Heterogeneity: $I^2$ = 0%, $\tau^2$ = 6.6196, $p$ = 1.0000
v = PIV
Buonsenso 2023 NP S PIV 2 5 0.40 [0.05; 0.85] 83
Haaramo 2024 NP S PIV 0 2 0.00 [0.00; 0.84] 118
Hammitt 2011 NP OP PIV 22 49 0.45 [0.31; 0.60] 533
Mackenzie 2019 NP OP PIV 15 32 0.47 [0.29; 0.65] 1557
Random effects model 88 0.44 [0.29; 0.61]
Heterogeneity: $I^2$ = 0%, $\tau^2$ = 0, $p$ = 0.9928
Random effects model 990 0.61 [0.48; 0.72]
Heterogeneity: $I^2$ = 79.9%, $\tau^2$ = 1.1557, $p$ < 0.0001
Test for subgroup differences: $\chi^2_8$ = 15.48, df = 8 ($p$ = 0.0504)
0 0.2 0.4 0.6 0.8 1
Sensitivity (Oral Specimen vs NP)

d)

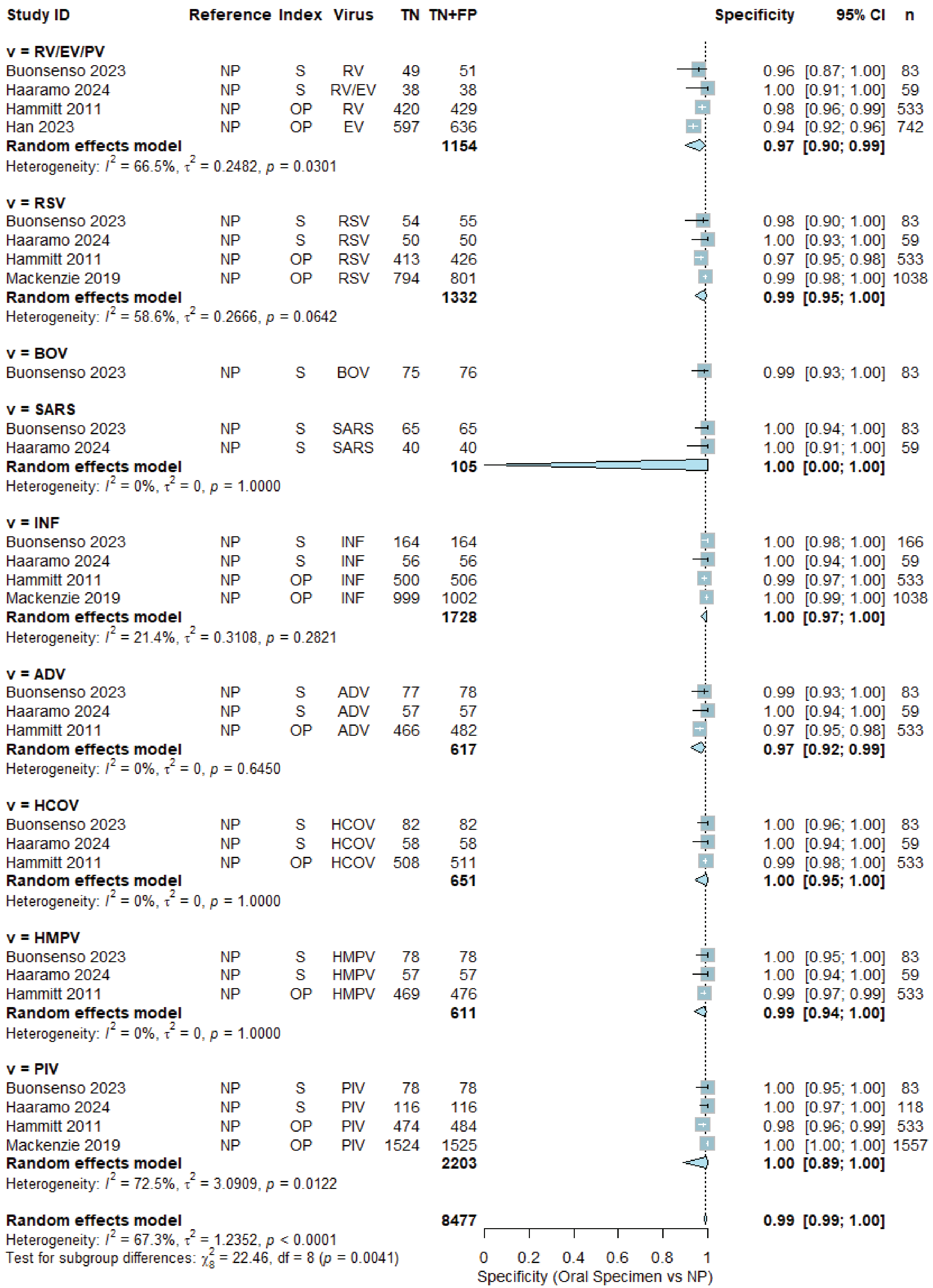
Study ID Reference Index Virus TN TN+FP Specificity 95% CI n
v = RV/EV/PV
Buonsenso 2023 NP S RV 49 51 0.96 [0.87; 1.00] 83
Haaramo 2024 NP S RV/EV 38 38 1.00 [0.91; 1.00] 59
Hammitt 2011 NP OP RV 420 429 0.98 [0.96; 0.99] 533
Han 2023 NP OP EV 597 636 0.94 [0.92; 0.96] 742
Random effects model 1154 0.97 [0.90; 0.99]
Heterogeneity: $I^2$ = 66.5%, $\tau^2$ = 0.2482, $p$ = 0.0301
v = RSV
Buonsenso 2023 NP S RSV 54 55 0.98 [0.90; 1.00] 83
Haaramo 2024 NP S RSV 50 50 1.00 [0.93; 1.00] 59
Hammitt 2011 NP OP RSV 413 426 0.97 [0.95; 0.98] 533
Mackenzie 2019 NP OP RSV 794 801 0.99 [0.98; 1.00] 1038
Random effects model 1332 0.99 [0.95; 1.00]
Heterogeneity: $I^2$ = 58.6%, $\tau^2$ = 0.2666, $p$ = 0.0642
v = BOV
Buonsenso 2023 NP S BOV 75 76 0.99 [0.93; 1.00] 83
v = SARS
Buonsenso 2023 NP S SARS 65 65 1.00 [0.94; 1.00] 83
Haaramo 2024 NP S SARS 40 40 1.00 [0.91; 1.00] 59
Random effects model 105 1.00 [0.00; 1.00]
Heterogeneity: $I^2$ = 0%, $\tau^2$ = 0, $p$ = 1.0000
v = INF
Buonsenso 2023 NP S INF 164 164 1.00 [0.98; 1.00] 166
Haaramo 2024 NP S INF 56 56 1.00 [0.94; 1.00] 59
Hammitt 2011 NP OP INF 500 506 0.99 [0.97; 1.00] 533
Mackenzie 2019 NP OP INF 999 1002 1.00 [0.99; 1.00] 1038
Random effects model 1728 1.00 [0.97; 1.00]
Heterogeneity: $I^2$ = 21.4%, $\tau^2$ = 0.3108, $p$ = 0.2821
v = ADV
Buonsenso 2023 NP S ADV 77 78 0.99 [0.93; 1.00] 83
Haaramo 2024 NP S ADV 57 57 1.00 [0.94; 1.00] 59
Hammitt 2011 NP OP ADV 466 482 0.97 [0.95; 0.98] 533
Random effects model 617 0.97 [0.92; 0.99]
Heterogeneity: $I^2$ = 0%, $\tau^2$ = 0, $p$ = 0.6450
v = HCOV
Buonsenso 2023 NP S HCOV 82 82 1.00 [0.96; 1.00] 83
Haaramo 2024 NP S HCOV 58 58 1.00 [0.94; 1.00] 59
Hammitt 2011 NP OP HCOV 508 511 0.99 [0.98; 1.00] 533
Random effects model 651 1.00 [0.95; 1.00]
Heterogeneity: $I^2$ = 0%, $\tau^2$ = 0, $p$ = 1.0000
v = HMPV
Buonsenso 2023 NP S HMPV 78 78 1.00 [0.95; 1.00] 83
Haaramo 2024 NP S HMPV 57 57 1.00 [0.94; 1.00] 59
Hammitt 2011 NP OP HMPV 469 476 0.99 [0.97; 0.99] 533
Random effects model 611 0.99 [0.94; 1.00]
Heterogeneity: $I^2$ = 0%, $\tau^2$ = 0, $p$ = 1.0000
v = PIV
Buonsenso 2023 NP S PIV 78 78 1.00 [0.95; 1.00] 83
Haaramo 2024 NP S PIV 116 116 1.00 [0.97; 1.00] 118
Hammitt 2011 NP OP PIV 474 484 0.98 [0.96; 0.99] 533
Mackenzie 2019 NP OP PIV 1524 1525 1.00 [1.00; 1.00] 1557
Random effects model 2203 1.00 [0.89; 1.00]
Heterogeneity: $I^2$ = 72.5%, $\tau^2$ = 3.0909, $p$ = 0.0122
Random effects model 8477 0.99 [0.99; 1.00]
Heterogeneity: $I^2$ = 67.3%, $\tau^2$ = 1.2352, $p$ < 0.0001
Test for subgroup differences: $\chi^2_8$ = 22.46, df = 8 ($p$ = 0.0041)
0 0.2 0.4 0.6 0.8 1
Specificity (Oral Specimen vs NP)

e)

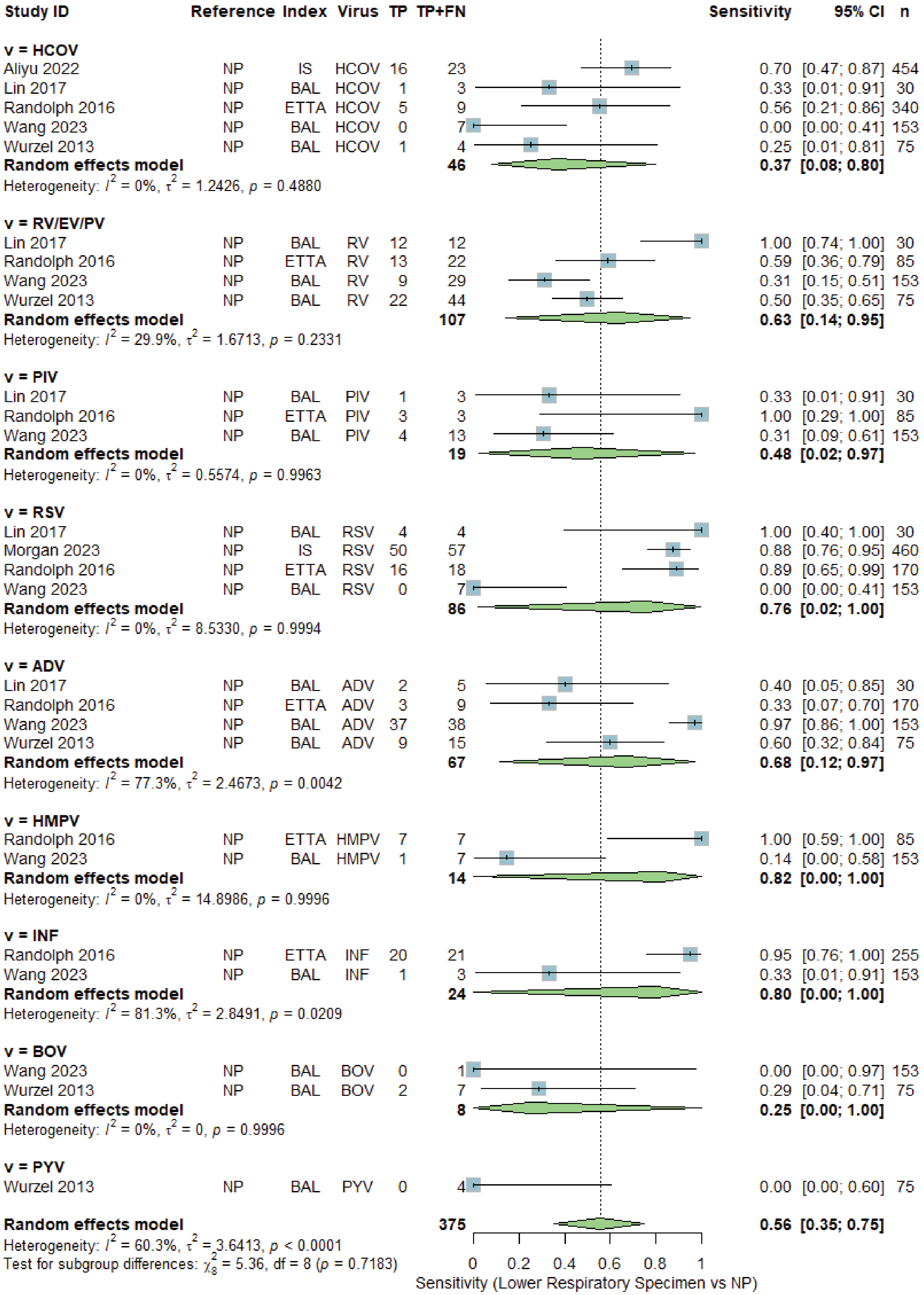
Study ID Reference Index Virus TP TP+FN Sensitivity 95% CI n
v = HCOV
Aliyu 2022 NP IS HCOV 16 23 0.70 [0.47; 0.87] 454
Lin 2017 NP BAL HCOV 1 3 0.33 [0.01; 0.91] 30
Randolph 2016 NP ETTA HCOV 5 9 0.56 [0.21; 0.86] 340
Wang 2023 NP BAL HCOV 0 7 0.00 [0.00; 0.41] 153
Wurzel 2013 NP BAL HCOV 1 4 0.25 [0.01; 0.81] 75
Random effects model 46 0.37 [0.08; 0.80]
Heterogeneity: $I^2$ = 0%, $\tau^2$ = 1.2426, $p$ = 0.4880
v = RV/EV/PV
Lin 2017 NP BAL RV 12 12 1.00 [0.74; 1.00] 30
Randolph 2016 NP ETTA RV 13 22 0.59 [0.36; 0.79] 85
Wang 2023 NP BAL RV 9 29 0.31 [0.15; 0.51] 153
Wurzel 2013 NP BAL RV 22 44 0.50 [0.35; 0.65] 75
Random effects model 107 0.63 [0.14; 0.95]
Heterogeneity: $I^2$ = 29.9%, $\tau^2$ = 1.6713, $p$ = 0.2331
v = PIV
Lin 2017 NP BAL PIV 1 3 0.33 [0.01; 0.91] 30
Randolph 2016 NP ETTA PIV 3 3 1.00 [0.29; 1.00] 85
Wang 2023 NP BAL PIV 4 13 0.31 [0.09; 0.61] 153
Random effects model 19 0.48 [0.02; 0.97]
Heterogeneity: $I^2$ = 0%, $\tau^2$ = 0.5574, $p$ = 0.9963
v = RSV
Lin 2017 NP BAL RSV 4 4 1.00 [0.40; 1.00] 30
Morgan 2023 NP IS RSV 50 57 0.88 [0.76; 0.95] 460
Randolph 2016 NP ETTA RSV 16 18 0.89 [0.65; 0.99] 170
Wang 2023 NP BAL RSV 0 7 0.00 [0.00; 0.41] 153
Random effects model 86 0.76 [0.02; 1.00]
Heterogeneity: $I^2$ = 0%, $\tau^2$ = 8.5330, $p$ = 0.9994
v = ADV
Lin 2017 NP BAL ADV 2 5 0.40 [0.05; 0.85] 30
Randolph 2016 NP ETTA ADV 3 9 0.33 [0.07; 0.70] 170
Wang 2023 NP BAL ADV 37 38 0.97 [0.86; 1.00] 153
Wurzel 2013 NP BAL ADV 9 15 0.60 [0.32; 0.84] 75
Random effects model 67 0.68 [0.12; 0.97]
Heterogeneity: $I^2$ = 77.3%, $\tau^2$ = 2.4673, $p$ = 0.0042
v = HMPV
Randolph 2016 NP ETTA HMPV 7 7 1.00 [0.59; 1.00] 85
Wang 2023 NP BAL HMPV 1 7 0.14 [0.00; 0.58] 153
Random effects model 14 0.82 [0.00; 1.00]
Heterogeneity: $I^2$ = 0%, $\tau^2$ = 14.8986, $p$ = 0.9996
v = INF
Randolph 2016 NP ETTA INF 20 21 0.95 [0.76; 1.00] 255
Wang 2023 NP BAL INF 1 3 0.33 [0.01; 0.91] 153
Random effects model 24 0.80 [0.00; 1.00]
Heterogeneity: $I^2$ = 81.3%, $\tau^2$ = 2.8491, $p$ = 0.0209
v = BOV
Wang 2023 NP BAL BOV 0 1 0.00 [0.00; 0.97] 153
Wurzel 2013 NP BAL BOV 2 7 0.29 [0.04; 0.71] 75
Random effects model 8 0.25 [0.00; 1.00]
Heterogeneity: $I^2$ = 0%, $\tau^2$ = 0, $p$ = 0.9996
v = PYV
Wurzel 2013 NP BAL PYV 0 4 0.00 [0.00; 0.60] 75
Random effects model 375 0.56 [0.35; 0.75]
Heterogeneity: $I^2$ = 60.3%, $\tau^2$ = 3.6413, $p$ < 0.0001
Test for subgroup differences: $\chi^2_8$ = 5.36, df = 8 ($p$ = 0.7183)
0 0.2 0.4 0.6 0.8 1
Sensitivity (Lower Respiratory Specimen vs NP)

f)

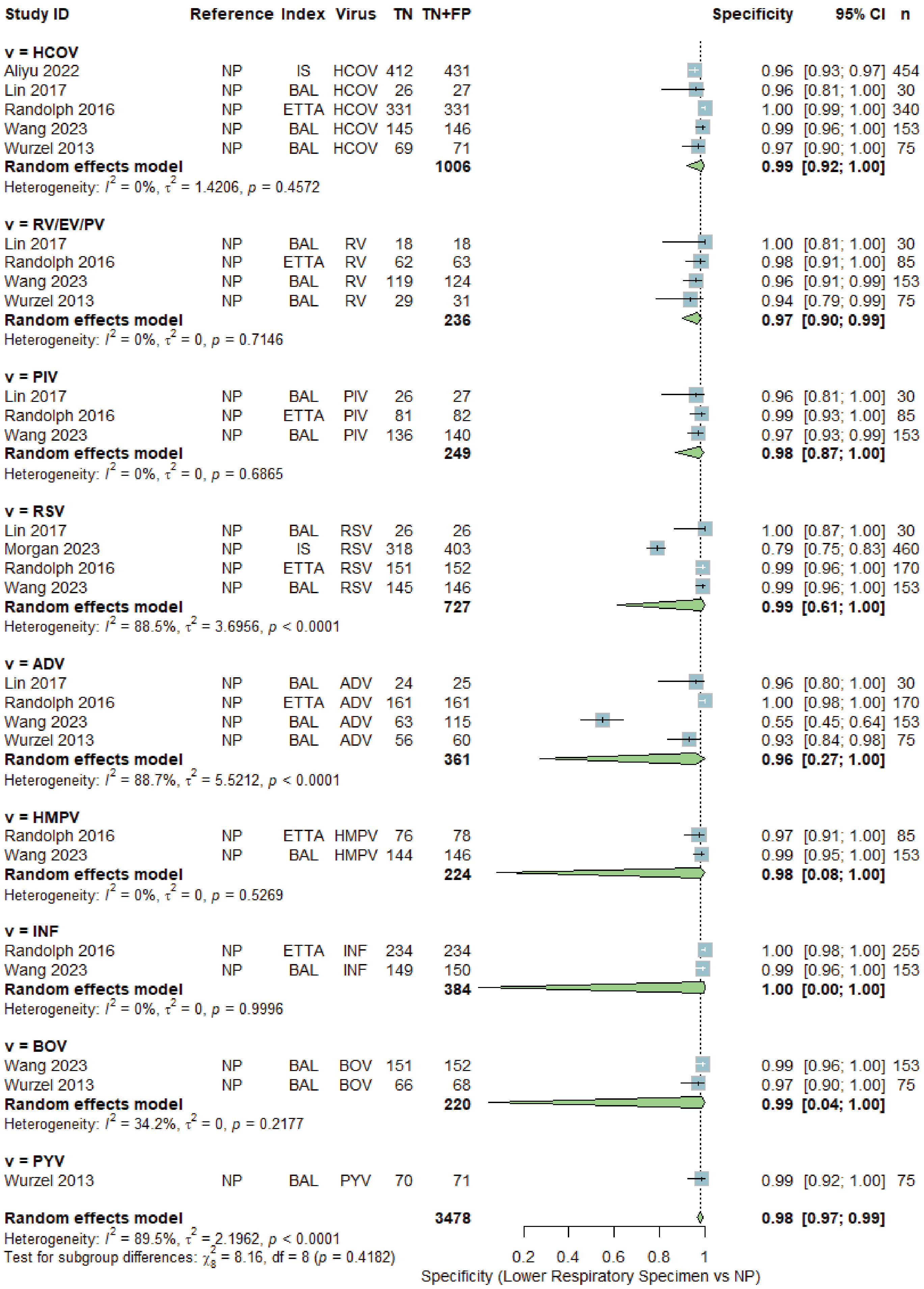

Study ID Reference Index Virus TN TN+FP Specificity 95% CI n
v = HCOV
Aliyu 2022 NP IS HCOV 412 431 0.96 [0.93; 0.97] 454
Lin 2017 NP BAL HCOV 26 27 0.96 [0.81; 1.00] 30
Randolph 2016 NP ETTA HCOV 331 331 1.00 [0.99; 1.00] 340
Wang 2023 NP BAL HCOV 145 146 0.99 [0.96; 1.00] 153
Wurzel 2013 NP BAL HCOV 69 71 0.97 [0.90; 1.00] 75
Random effects model 1006 0.99 [0.92; 1.00]
Heterogeneity: $I^2$ = 0%, $\tau^2$ = 1.4206, $p$ = 0.4572
v = RV/EV/PV
Lin 2017 NP BAL RV 18 18 1.00 [0.81; 1.00] 30
Randolph 2016 NP ETTA RV 62 63 0.98 [0.91; 1.00] 85
Wang 2023 NP BAL RV 119 124 0.96 [0.91; 0.99] 153
Wurzel 2013 NP BAL RV 29 31 0.94 [0.79; 0.99] 75
Random effects model 236 0.97 [0.90; 0.99]
Heterogeneity: $I^2$ = 0%, $\tau^2$ = 0, $p$ = 0.7146
v = PIV
Lin 2017 NP BAL PIV 26 27 0.96 [0.81; 1.00] 30
Randolph 2016 NP ETTA PIV 81 82 0.99 [0.93; 1.00] 85
Wang 2023 NP BAL PIV 136 140 0.97 [0.93; 0.99] 153
Random effects model 249 0.98 [0.87; 1.00]
Heterogeneity: $I^2$ = 0%, $\tau^2$ = 0, $p$ = 0.6865
v = RSV
Lin 2017 NP BAL RSV 26 26 1.00 [0.87; 1.00] 30
Morgan 2023 NP IS RSV 318 403 0.79 [0.75; 0.83] 460
Randolph 2016 NP ETTA RSV 151 152 0.99 [0.96; 1.00] 170
Wang 2023 NP BAL RSV 145 146 0.99 [0.96; 1.00] 153
Random effects model 727 0.99 [0.61; 1.00]
Heterogeneity: $I^2$ = 88.5%, $\tau^2$ = 3.6956, $p$ < 0.0001
v = ADV
Lin 2017 NP BAL ADV 24 25 0.96 [0.80; 1.00] 30
Randolph 2016 NP ETTA ADV 161 161 1.00 [0.98; 1.00] 170
Wang 2023 NP BAL ADV 63 115 0.55 [0.45; 0.64] 153
Wurzel 2013 NP BAL ADV 56 60 0.93 [0.84; 0.98] 75
Random effects model 361 0.96 [0.27; 1.00]
Heterogeneity: $I^2$ = 88.7%, $\tau^2$ = 5.5212, $p$ < 0.0001
v = HMPV
Randolph 2016 NP ETTA HMPV 76 78 0.97 [0.91; 1.00] 85
Wang 2023 NP BAL HMPV 144 146 0.99 [0.95; 1.00] 153
Random effects model 224 0.98 [0.08; 1.00]
Heterogeneity: $I^2$ = 0%, $\tau^2$ = 0, $p$ = 0.5269
v = INF
Randolph 2016 NP ETTA INF 234 234 1.00 [0.98; 1.00] 255
Wang 2023 NP BAL INF 149 150 0.99 [0.96; 1.00] 153
Random effects model 384 1.00 [0.00; 1.00]
Heterogeneity: $I^2$ = 0%, $\tau^2$ = 0, $p$ = 0.9996
v = BOV
Wang 2023 NP BAL BOV 151 152 0.99 [0.96; 1.00] 153
Wurzel 2013 NP BAL BOV 66 68 0.97 [0.90; 1.00] 75
Random effects model 220 0.99 [0.04; 1.00]
Heterogeneity: $I^2$ = 34.2%, $\tau^2$ = 0, $p$ = 0.2177
v = PYV
Wurzel 2013 NP BAL PYV 70 71 0.99 [0.92; 1.00] 75
Random effects model 3478 0.98 [0.97; 0.99]
Heterogeneity: $I^2$ = 89.5%, $\tau^2$ = 2.1962, $p$ < 0.0001
Test for subgroup differences: $\chi^2_8$ = 8.16, df = 8 ($p$ = 0.4182)
0.2 0.4 0.6 0.8 1
Specificity (Lower Respiratory Specimen vs NP)
